# New-onset syncope in diabetic patients treated with sodium-glucose cotransporter-2 inhibitors versus dipeptidyl peptidase-4 inhibitors: A Chinese population-based cohort study

**DOI:** 10.1101/2023.07.04.23292207

**Authors:** Xinyi Gao, Nan Zhang, Lei Lu, Tianyu Gao, Oscar Hou In Chou, Wing Tak Wong, Carlin Chang, Abraham Ka Chung Wai, Gregory Y. H. Lip, Qingpeng Zhang, Gary Tse, Tong Liu, Jiandong Zhou

## Abstract

**Background and Aims:** Syncope and post-syncopal adverse events lead to a heavy burden in the healthcare systems with negative impact on the economy globally. However, no effective treatments have been identified to prevent the risk of new-onset syncope. This study compared the preventive effect of incident syncope between sodium-glucose cotransporter-2 inhibitor (SGLT2i) and dipeptidyl peptidase-4 inhibitor (DPP4i).

**Methods:** This was a retrospective, territory-wide cohort study enrolling type 2 diabetes mellitus (T2DM) patients treated with SGLT2i or DPP4i between January 1st, 2016, and December 31st, 2020, in Hong Kong, China. The outcomes were new-onset syncope, cardiovascular mortality, and all-cause mortality. Multivariable Cox regression and different approaches using the propensity score were used to evaluate the association between SGLT2i vs. DPP4i with incident syncope and mortality.

**Results:** After matching, a total of 37502 patients with T2DM were included (18751 SGLT2i users, 18751 DPP4i users). During a median follow-up of 5.56 years, compared to DPP4i users, SGLT2i therapy was associated with a 51% lower risk of new-onset syncope (HR, 0.49; 95%CI [0.41-0.57], P<0.001), 65% lower risk of cardiovascular mortality (HR, 0.35; 95%CI [0.26-0.46], P<0.001), and a 70% lower risk of all-cause mortality (HR, 0.30; 95%CI [0.26-0.34], P<0.001) in the fully adjusted model. Similar association with syncope was observed for dapagliflozin (HR, 0.70; 95%CI [0.58-0.85], P<0.001), canagliflozin (HR, 0.48; 95%CI [0.36-0.63], P<0.001) and ertuglifolzin (HR, 0.45; 95%CI [0.30-0.68], P<0.001), but was attenuated for empagliflozin (HR, 0.79; 95%CI [0.59-1.05], P=0.100) after adjusting for potential confounders. Subgroup analyses suggested that, compared to DPP4i, SGLT2i showed a significantly protective effect in incident syncope among T2DM patients, regardless of gender, age, comorbidities burden and other medication history, as well as among patients with different levels of fasting glucose, HbA1c, and glycemic variability.

**Conclusions:** Compared to DPP4i, SGLT2i could significantly reduce the risk of new-onset syncope in patients with T2DM, regardless of gender, age, comorbidities, other medication history, and degree of glycemic control. Our findings suggest a promising future of SGLT2i in preventing incident syncope.

Structured graphical abstract CI: confidence interval; DPP4i: dipeptidyl peptidase-4 inhibitor; HR: hazard ratio; SGLT2i: sodium-glucose cotransporter-2 inhibitor.

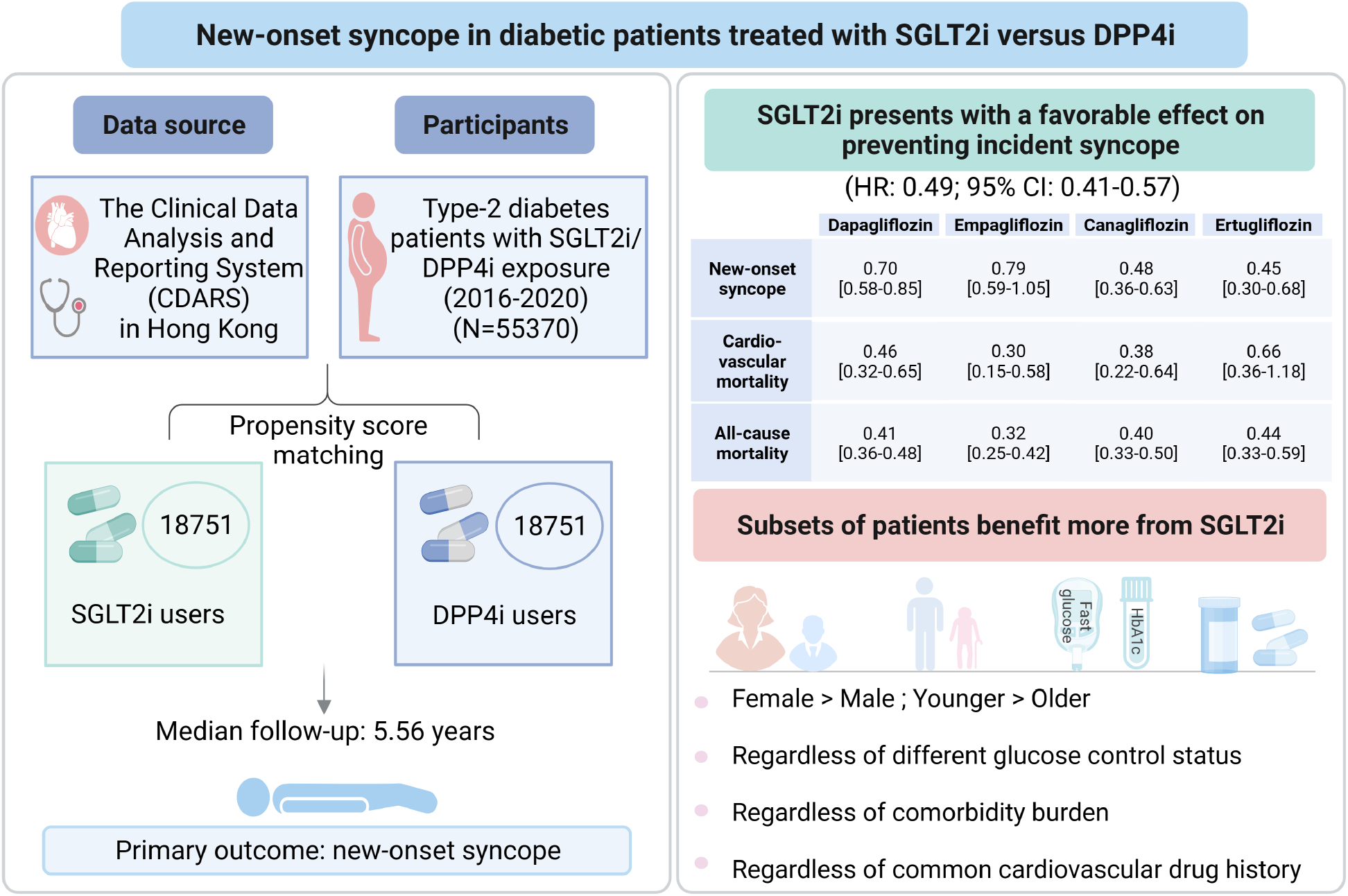

## Introduction

Syncope is defined as a condition of transient loss of consciousness due to global cerebral hypo-perfusion and characterized by rapid, self-limiting onset and complete recovery. It is a common problem that affects all age groups, which accounts for 1-3% of emergency department visits and 6% of hospitalizations, with a lifetime cumulative incidence up to 40% [1, 2]. Syncope serves as a common presentation of a number of clinical conditions ranging from benign to life-threatening diseases, and its causes are difficult to diagnose [3]. Consequently, it often results in unnecessary hospital admissions, multiple consultations, and the performance of many diagnostic tests, which poses a substantial economic burden [4]. Beyond the economic impact, syncope is associated with impaired quality of life, significant morbidity and mortality [5]. It has been reported that the mortality rate is 6.9% and 25.2% at 10 days and 2 years after syncope, respectively [6]. Therefore, it is of the utmost importance to develop some novel agents to prevent incident syncope or to identify the syncope-prevention effect of some old classic drugs, in order to reduce the burden of syncope. However, to our knowledge, management of syncope remains challenging and the field of pharmacologic prevention of incident syncope is still blank until now.

Diabetes represents an important risk factor for syncope, which has been associated with a higher recurrence rate [7, 8]. The latest class of anti-diabetic agent, sodium-glucose cotransporter-2 inhibitor (SGLT2i), have received significant attention owing to their beneficial effects on cardiovascular events, especially in the context of heart failure [9-15]. Interestingly, a recent study which included 324 patients with type 2 diabetes mellitus (T2DM) and vasovagal syncope (VVS), reported that SGLT2i could significantly reduce the risk of VVS recurrence during 1-year follow-up, compared to non-SGLT2i anti-diabetic agents [16]. Meanwhile, another novel class of hypoglycemic agent, the dipeptidyl peptidase-4 inhibitor (DPP4i), has been associated with favorable or neutral cardioprotective effects than non-users [17-20]. SGLT2i and DPP4i represent two promising anti-diabetic agents, both of them are widely used oral preparations in clinical practice. However, no study has investigated the effects of SGLT2i and DPP4i in preventing the new-onset syncope. Therefore, this study used a population-based cohort to compare the prophylactic effects of SGLT2i vs. DPP4i on new-onset syncope, and to explore the subsets of patients who may benefit more from these treatments.

## Method

### Study Design and Population

This study was granted approval by the Institutional Review Board of the University of Hong Kong/Hospital Authority Hong Kong West Cluster and The Joint Chinese University of Hong Kong–New Territories East Cluster Clinical Research Ethics Committee. This was a retrospective, territory-wide cohort study of T2DM patients on either SGLT2i or DPP4i with the first index prescription date between January 1^st^, 2016, and December 31^st^, 2020, in Hong Kong.

Patients of this study have been identified from the Clinical Data Analysis and Reporting System (CDARS), which centralizes information on patients from individual local hospitals to establish comprehensive medical data throughout the Hong Kong. The system has been previously utilized by multiple teams to conduct population-based cohort studies [21-24], including those focused on diabetes [25]. All pertinent data including clinical characteristics, disease diagnoses, laboratory results and drug treatment details based on the population in Hong Kong can be extracted from this system. In the current study, patients aged 18 years and older with T2DM who were prescribed and regularly taken SGLT2i (dapagliflozin, canagliflozin, empagliflozin, or ertugliflozin) or DPP4i (vildagliptin, sitagliptin, saxagliptin, alogliptin, or linagliptin) during the indicated period were enrolled and followed up until death or December 31, 2020. Patients were excluded if one of the following criteria was met: 1) received both SGLT2i and DPP4i; 2) had exposed to SGLT2i or DPP4i less than 1 month; 3) with prior diagnosis of ventricular tachycardia, ventricular fibrillation, sudden cardiac death, congenital long QT syndrome or syncope before the initial drug exposure.

### Data collection

All extracted covariates of interest were displayed in **Table 1** for confounder adjustment, including patient demographic characteristics i.e., gender and age at initiation of SGLT2i/DPP4i, prior comorbidities, Charlson’s standard comorbidity index, frequency and duration of SGLT2i/DPP4i exposure, and other medication histories. Baseline laboratory data, including complete blood count, indicators of liver and kidney function, lipid and glucose profiles were also extracted. Prior comorbidities were documented in CDARS under the International Classification of Diseases Ninth Revision (ICD-9) codes (**Table S1**), as CDARS has not implemented the International Classification of Diseases Tenth Revision (ICD-10) codes for disease diagnoses to date [26, 27].

### Outcomes

The primary outcome of this study was hospitalization for new-onset syncope, which was identified from the CDARS using the ICD-9 code 780.2 “syncope and collapse”. Secondary outcome were cardiovascular mortality and all-cause mortality. Cardiovascular mortality was identified by ICD-10 codes: I00-I09, I11, I13, and I20-I51. The mortality data was documented in the Hong Kong Death Registry, a population-based official government registry with the registered death records of all Hong Kong citizens linked to CDARS under ICD-10 codes [28].

### Statistical Analysis

Baseline characteristics of patients treated with SGLT2i and DPP4i was summarized using descriptive statistics. Continuous variables were presented as median [95% confidence interval (CI)/ interquartile range] or mean [standard deviation (SD)] and categorical variables was presented as total number (percentage). Continuous variables were compared using the two-tailed Mann‒Whitney U test, whilst the two-tailed χ2 test with Yates’ correction was used to test 2 × 2 contingency data.

Propensity score matching (PSM) was used to generate SGLT2i and DPP4i cohorts in a 1:1 ratio using the nearest neighbour matching strategy. All of the variables extracted that may influence treatment selection and outcomes of interest including demographic characteristics, comorbidities, non-SGLT2i/DPP4i drugs, and biochemical indicators were incorporated. Variable selection was based on clinical reasoning, not statistical significance [29]. Baseline and clinical characteristics of patients with/without new-onset syncope were compared before and after PSM using standardised mean differences (SMD). Univariable Cox models were used to identify significant risk factors for the outcomes. Further, multivariate Cox regression analyses were sequentially fitted with 5 models to fully estimate the adjusted hazard ratio (HR) and 95% CI for the outcomes.

Subgroup analyses were conducted according to age (<65 and >65 years), gender, Charlson’s comorbidity index, individual comorbidities, medication history, and a spectrum of glucose measurements, including baseline and mean levels of HbA1c and fasting glucose, as well as glycemic variability assessed by derivation of variance and coefficient of variation (CV). Mean levels of HbA1c and fasting glucose were calculated based on the collected tests before initial drug exposure. The glycemic variability are calculated based on at least three measurements of HbA1c and fasting glucose [30].

To test the robustness of our results, different sensitivity analyses were performed. First, alternative propensity score approaches, including propensity score stratification, inverse probability of treatment weighting (IPTW) and stable inverse probability of treatment weighting (SIPTW), as well as competing risk analyses using cause-specific and sub-distribution hazard models were conducted [31-33]. Second, patients with baseline immune-mediated inflammation and cancer which might affect the outcomes were excluded. Third, patients with chronic kidney disease (CKD) stage 4/5 (estimated glomerular filtration rate <30 ml/min/1.73 m ^2^), peritoneal dialysis or hemodialysis at baseline were excluded to further validate our results. Fourth, a 1-year lag time approach was applied, which could improve drug-outcome association estimates in presence of protopathic bias and minimize time-lag effect of treatment [34]. Besides, marginal effects analyses were performed to explore whether the relationship between SGLT2i/DPP4i exposure and syncope was modulated by other variables [35].

Negligible post-weighting inter-group standardized mean difference was defined as an SMD < 0.2 [21]. The HR, 95% CI and P value will be reported. P value <0.05 is considered statistically significant. The statistical analysis was performed with RStudio software (Version 1.1.456) and Python (Version 3.6).

## Results

### Baseline characteristics

A total of 76147 T2DM patients treated with SGLT2i or DPP4i were identified between January 1, 2015 and December 31, 2020. After exclusion, 55370 patients (50.48% males; mean age, 63.2 years) were enrolled, including 18751 (33.86%) SGLT2i users and 36619 (66.14%) DPP4i users. Of the 18751 SGLT2i users, 10549 were treated with dapagliflozin (56.25%), 3913 with empagliflozin (20.86%), 4998 with canagliflozin (26.65%) and 2509 with ertugliflozin (13.38%). Over a median follow-up of 5.56 (5.24, 5.80) years, 1374 patients were hospitalization for new-onset syncope [incidence rate (IR), 2.48%], and 6000 patients died from any cause (IR: 10.83%), among which 1762 deaths (IR: 3.18%) were associated with cardiovascular causes (**Figure 1**).

**Figure 1.**
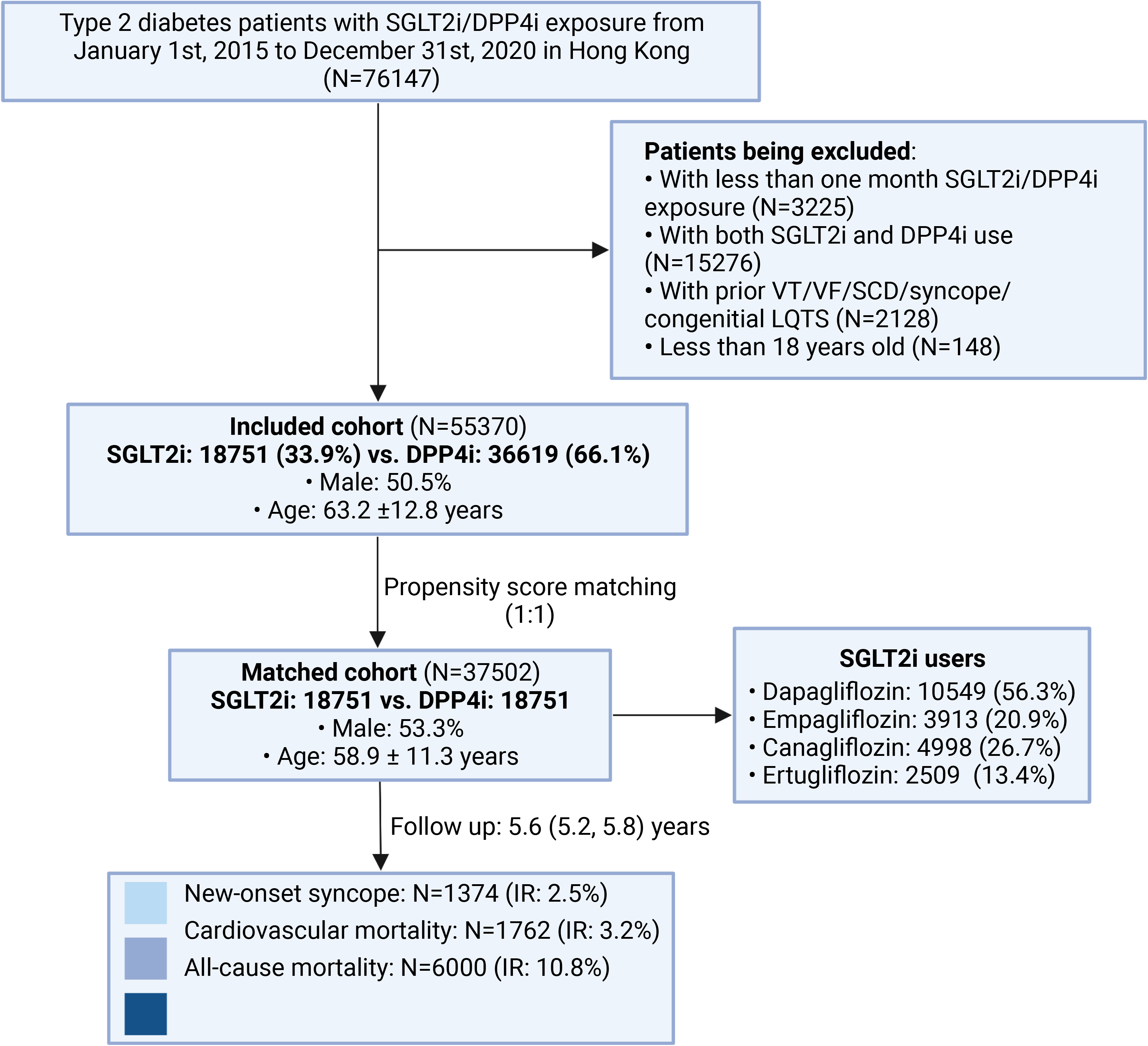
Flowchart of data processing. DPP4i: dipeptidyl peptidase-4 inhibitor; IR: incidence rate; LQTS: long QT interval syndrome; SCD: sudden cardiac death; SGLT2i: sodium-glucose cotransporter-2 inhibitor; VF: ventricular fibrillation; VT: ventricular tachycardia.

After PSM, majority of baseline characteristics between SGLT2i users and DPP4i users were balanced, with the exception of overall age and SD of high-density lipoprotein (SMD > 0.2), and 37502 individuals were analyzed (53.31% males; median age, 58.9 years) (**Table 1** and **Figure S1**). Baseline characteristics of the included patients according to the occurrence of syncope before and after 1:1 PSM are presented in **Table S2**.

### The association between SGLT2i vs. DPP4i and incident syncope

During a median follow-up of 5.56 years, a significantly lower cumulative event rate of new-onset syncope was observed in patients treated with SGLT2i, in relation to DPP4i users (**Figure 2**). The annual person-year incidence ratios of incident syncope in the matched cohort are summarized in **Figure S2** and **Table S3,** patients treated with SGLT2i had smaller person-year incidence ratio in each year of follow-up duration than those with DPP4i exposure. In univariable Cox regression, SGLT2i therapy was associated with a significantly lower risk of new-onset syncope (HR, 0.43; 95%CI [0.37-0.49], P<0.001) (**Table S4**). The results remained stable after adjusting for potential confounders and were consistent across five models (**Table 2**). In the fully adjusted model (Model 5), SGLT2i users presented with an over 50% lower risk of new-onset syncope (HR, 0.49; 95%CI [0.41-0.57], P<0.001) than the DPP4i users.

**Figure 2.**
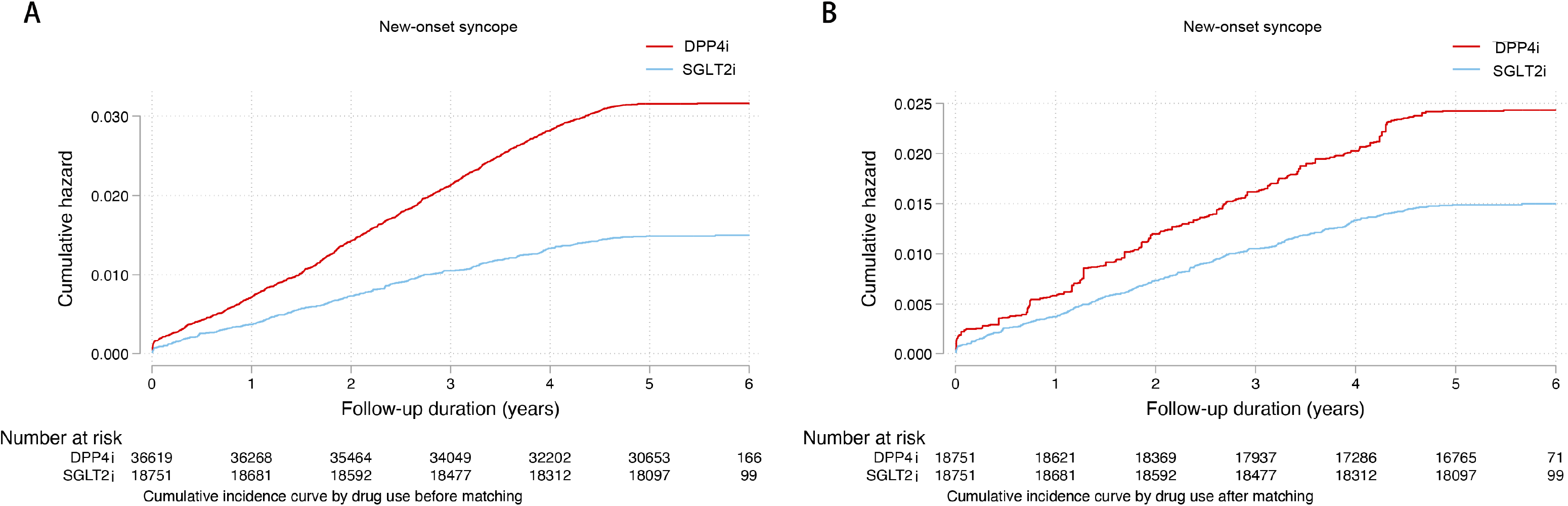
Cumulative incidence curves for new-onset syncope stratified by drug exposure effects of SGLT2i and DPP4i before (A) and after (B) matching (1:1). DPP4i: dipeptidyl peptidase-4 inhibitor; SGLT2i: sodium-glucose cotransporter-2 inhibitor.

Amongst different SGLT2i, dapagliflozin (HR, 0.70; 95%CI [0.58-0.85], P<0.001), canagliflozin (HR, 0.48; 95%CI [0.36-0.63], P<0.001) and ertugliflozin (HR, 0.45; 95%CI [0.30-0.68], P<0.001) were associated with significantly lower risks of new-onset syncope. Whereas the preventive effect of empagliflozin on incident syncope was attenuated after adjusting for potential confounders (HR, 0.79; 95%CI [0.59-1.05], P=0.100) (**Table 2**).

## Results of subgroup analyses

### Subgroup analysis according to gender and age

In subgroup analysis according to gender, compared to DPP4i users, both the female (HR, 0.54; 95%CI [0.42-0.70], P<0.001) and male (HR, 0.66; 95%CI [0.55-0.80], P<0.001) SGLT2i users experienced a significantly lower risk of new-onset syncope. In addition, compare to male SGLT2i users, female patients treated with SGLT2i showed a more prominently decreased risk of syncope, which suggested that both female and male patients could benefit from SGLT2i therapy in preventing incident syncope, whereas female patients might benefit more than male (**Figure S3**). In subgroup analysis according to age, both the younger (<65 years, HR, 0.59; 95%CI [0.47-0.73], P<0.001) and older (>65 years, HR, 0.71; 95%CI [0.58-0.87], P=0.001) SGLT2i users showed a significantly reduced risk of syncope than DPP4i users (**Figure S3**). Amongst the SGLT2i users, younger patients showed a more prominently lower risk of new-onset syncope than the older patients (**Figure 3**).

**Figure 3.**
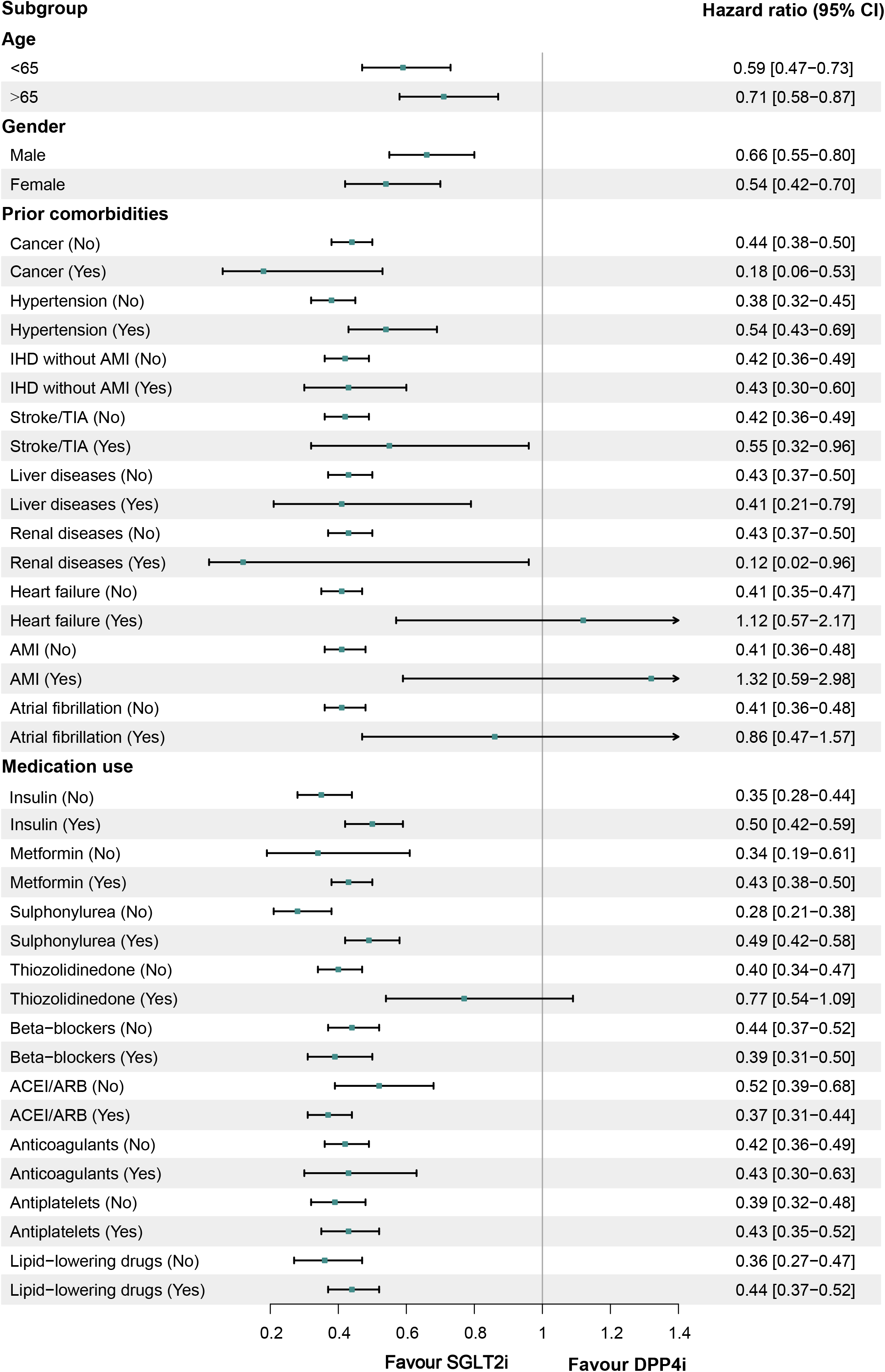
Subgroup analysis for new-onset syncope stratified by drug exposure effects of SGLT2i versus DPP4i in the matched cohort (1:1). ACEI: angiotensin-converting enzyme inhibitor; AMI: acute myocardial infarction; ARB: angiotensin receptor blocker; CI: confidence interval; IHD: ischemic heart disease; TIA: transient ischemic attack.

### Subgroup analyses according to glucose measurements

To investigate the effects of different glucose control status on the association between SGLT2i vs. DPP4i and syncope, subgroup analyses according to a spectrum of glucose measurements, including baseline and mean level, as well as variability of HbA1c and fasting glucose, were conducted (**Table 3**). Compared with DPP4i, SGLT2i treatment could significantly reduce the incidence of new-onset syncope in patients with different levels of baseline HbA1c (less than 7.5%, between 7.5% and 9%, and greater than 9%), as well as in different quartile subgroups of mean HbA1c, and variance/CV of HbA1c. We also observed such protection effects of individual SGLT2i treatments including dapagliflozin, empagliflozin, canagliflozin, and ertugliflozin, in most of the subgroups. Similar findings of the protection effects of SGLT2i vs. DPP4i were obtained in subgroups of fasting glucose.

### Subgroup analyses according to comorbidities and medication history

In addition, we observed a consistently protective effect of SGLT2i in preventing future syncope occurrence among T2DM patients with different levels of Charlson’s comorbidity index (from 0 to 5+), compared to DPP4i (**Table S5**). However, there was also a signal that patients with lower Charlson’s comorbidity index might benefit more from individual SGLT2i treatment than those with higher comorbidity burden. Further analyses according to individual comorbidities were performed (**Table S6**). Compared to DPP4i, SGLT2i could significantly reduce the risk of incident syncope in T2DM patients complicated with cancer (HR, 0.18; 95%CI [0.06-0.53], P=0.002), hypertension (HR, 0.54; 95%CI [0.43-0.69], P<0.001), non-acute myocardial infarction ischemic heart disease (IHD) (HR, 0.43; 95%CI [0.30-0.60], P<0.001), stroke/transient ischemic attack (TIA) (HR, 0.55; 95%CI [0.32-0.96], P=0.036), liver diseases (HR, 0.41; 95%CI [0.21-0.79], P=0.008), and renal diseases (HR, 0.12; 95%CI [0.02-0.96], P=0.046), but not significant among those complicated with other comorbidities listed in this study (**Figure 3**).

Furthermore, compare to DPP4i, SGLT2i presented a favorable effect on preventing incident syncope, regardless of baseline medication history of antihypertensive drugs (such as angiotensin-converting enzyme inhibitor [ACEI]/angiotensin receptor blocker [ARB], beta-blockers, calcium channel blockers, and diuretics), antiplatelet drugs, lipid-lowering drugs (statins and fibrates), and most antidiabetic drugs (such as metformin, sulphonylurea, and insulin) (**Figure 3** and **Table S7**).

### Results of sensitivity analyses and marginal effects analysis

Results from several sensitivity analyses are in agreement with the primary analysis. Compared to DPP4i, SGLT2i therapies were associated with significantly decreased risk of new-onset syncope when using alternative propensity score approaches, including propensity score stratification (HR, 0.32; 95%CI [0.25-0.53], P<0.001), IPTW (HR, 0.39; 95%CI [0.34-0.45], P<0.001), and SIPTW (HR, 0.49; 95%CI [0.42-0.63], P<0.001) or applying cause-specific (HR, 0.29; 95%CI [0.21-0.35], P<0.001) and sub-distribution hazard model (HR, 0.35; 95%CI [0.28-0.52], P<0.001) (**Table S8**). The association between SGLT2i and reduced syncope risk was stable when excluding patients with baseline immune-mediated inflammatory diseases and cancer (HR, 0.42; 95%CI [0.37-0.62], P <0.001) (**Table S9**), or applying 1-year lag time approach (HR, 0.43; 95%CI [0.37-0.49], P<0.001) (**Table S10**). In addition, after excluding patients with stage 4/5 CKD, peritoneal dialysis or haemodialysis, both the overall SGLT2is (HR, 0.47; 95%CI [0.41-0.55], P<0.001) and four individual SGLT2i, including empagliflozin (HR, 0.70; 95%CI [0.53-0.91], P=0.009) showed a significantly superior protective effect on incident syncope than DPP4i (**Table S11**).

To further clarify whether the relationship between SGLT2i/DPP4i exposure and syncope was modulated by other variables, marginal effects analyses were performed. The results demonstrated a superiorly protective effect of SGLT2i than DPP4i on incident syncope, regardless of the age at initial drug exposure, baseline Charlson’s index, number of previous hospitalizations, duration of T2DM, and number of prior anti-diabetic drugs (**Figure S4**).

### The association between SGLT2i vs DPP4i and death outcomes

Apart from syncope, SGLT2i also showed a significantly protective effect on cardiovascular mortality and all-cause mortality, compared to DPP4i (**Figure S5**). In the fully adjusted model, patients treated with SGLT2i showed a 65% and 70% reduced risk of cardiovascular mortality (HR, 0.35; 95%CI [0.26-0.46], P<0.001) and all-cause mortality (HR, 0.30; 95%CI [0.26-0.34], P<0.001), compared to DPP4i (**Table 2**). The results of the subgroup analyses and sensitivity analyses regarding the association between SGLT2i vs. DPP4i with mortality were in line with the primary analysis (**Figure S6-7** and **Table S8** and **Table S10-14**).

## Discussion

In this population-based cohort study, several key findings were noted. First, SGLT2i treatment was associated with significantly lower risk of new-onset syncope, cardiovascular mortality and all-cause mortality compared to DPP4i therapy, especially for dapagliflozin, canagliflozin and ertuglifolzin, which were consistent across different models and through comprehensive sensitivity analyses. Second, compared to DPP4i, SGLT2i showed a significantly protective effect on incident syncope, regardless of gender and age, whereas female and younger patients might benefit more than male and older patients, respectively. Third, compared with DPP4i, SGLT2i treatment could significantly reduce the risk of new-onset syncope among T2DM patients with different levels of baseline and mean HbA1c and fasting glucose, and various degree of glycemic variability. Fourth, SGLT2i showed a significantly favorable effect on preventing incident syncope, regardless of comorbidities burden and other medication use.

To our knowledge, there was only one previous study focusing on anti-diabetic agents and syncope [16]. The SCAN study has included 324 T2DM patients with VVS (161 SGLT2i users and 163 non-SGLT2i users), and observed that SGLT2i significantly reduced the risk of VVS recurrence during 1-year follow-up compared to non-SGLT2i treatments [16]. However, the SCAN study only included patients with VVS and focused on the recurrent but not incident risk of syncope. Compared to the previous study, our study has several strengths. First, our study is a large-scale, population-based cohort including 55370 T2DM patients, and with a relatively long-term follow-up duration (5.56 years), which could provide reliable estimation of the association between SGLT2i and syncope. Second, instead of investigating the recurrence risk, our study has focused on the risk of new-onset syncope, which has never been addressed before and could provide some novel insights into the prevention strategy of syncope. Third, DPP4i has been widely used in clinical practice and associated with favorable or neutral cardioprotective effects [17-20], which has been used as comparator in numerous large-scale studies focusing on the effects of SGLT2i in various clinical outcomes [17, 36]. Some studies have compared the effects between SGLT2i and glucagon-like peptide-1 receptor agonist (GLP-1RA), another promising class of antidiabetic agent, in clinical outcomes, which yield controversial results. We have also conducted an additional three-arm (SGLT2i vs. DPP4i vs. GLP-1RA) as-treat analysis presented in Table S15, which observed a favorable signal for SGLT2i in preventing syncope than GLP-1RA, whereas the difference did not reach the traditional significance (HR, 0.89; 95%CI [0.66-1.19], P=0.382). However, the sample size of the GLP-1RA arm in our database is relatively small, which may introduce selection bias. Therefore, using DPP4i as comparator in our study could guarantee the reliability of our results and also facilitate choosing appropriate preventive therapeutic options. In addition, different from the SCAN study [16], our primary outcome was anchored on overall syncope rather than on a specific type of syncope classification. The accurate etiology of syncope is hard to identify in real-world settings, with up to 42.1% patients having the possibility to be diagnosed with unknown-cause of syncope and a considerable portion of patient with misclassified causes of syncope [37]. Moreover, the accuracy rate of head-up tilt test, which could facilitate the diagnosis of reflex syncope, is only about 60% [38, 39]. Therefore, it is appropriate to focus on the overall syncope population, especially when there lacks a golden standard to accurately identify the etiology of syncope.

Another finding of this study is that empagliflozin is the only SGLT2i of the 4 studied that failed to reach statistical significance on incident syncope in the fully adjusted model, whereas the association between other three SGLT2i and syncope existed across five models, which might suggests a more prominently protective effect of dapagliflozin, canagliflozin and ertuglifolzin on incident syncope. Interestingly, it should be noticed that after applying 1-year lag time approach or excluding patients with stage 4/5 CKD, peritoneal dialysis or haemodialysis, all of these four SGLT2is (including empagliflozin) showed significantly favorable effects on preventing syncope. Some previous studies have also observed a difference in pharmacologic effects on various outcomes between individual SGLT2i [9, 40, 41]. The mechanism underlying the observed heterogeneity between different SGLT2i and syncope remains unclear, which might be due to the difference in SGLT2 selectivity [42] and other non-SGLT2 mediated mechanisms [43]. For instance, prior study has demonstrated off-target effects of SGLT2i, including the direct effect on the sodium-hydrogen exchanger 1 (NHE1) in the heart, NHE3 in the kidney, and NHE9 in inflammatory cells that could impact cardiac and kidney outcomes [44]. Hence, further investigation into the potential mechanisms of such observations and to what degree these effects differ between members of the class is of utmost importance.

Moreover, our findings suggest that extensive groups of individuals could benefit from SGLT2i therapy in reducing incident syncope, which possess important clinical implications and has the potential to improve syncope preventive strategy. First, SGLT2i is superior to DPP4i in reducing the risk of new-onset syncope among patients with T2DM, regardless of gender and age. Whereas compared to male, female SGLT2i users have a lower incidence rate of syncope, which might suggest a more beneficial role of SGLT2i in female population than male in the syncope settings. Our study also observed that younger SGLT2i users have a lower risk of incident syncope than older users, which may be, at least partially, attributed to the multimorbidity, aging and frailty of the older populations [45]. However, this observation may also imply that younger patients could benefit more from SGLT2i treatment in preventing syncope than the older patients, and that remains to be further investigated. Second, SGLT2i could lower the risk of incident syncope among T2DM patients with varying degrees of glycemic control, namely among patients with different levels of fasting glucose, HbA1c, and glycemic variability. Third, SGLT2i therapy prevents incident syncope in patients with T2DM, regardless of comorbidities burden (Charlson’s comorbidity index from 0 to 5+). More specifically, SGLT2i could reduce the risk of new-onset syncope among T2DM patients complicated with hypertension, IHD, renal diseases, or stroke/TIA, which have been demonstrated as risk factors for syncope [8]. Fourth, it has been reported that individuals using cardiac medications were at increased risk for syncope [8]. Our finding suggests that SGLT2i significantly reduces the risk of syncope among T2DM patients, regardless of medication history, such as antihypertensive drugs (ACEI/ARB, beta-blockers, calcium channel blockers, and diuretics), antiplatelet drugs, lipid-lowering drugs, and other antidiabetic drugs. Therefore, SGLT2i holds a promising future in preventing incident syncope.

The underlying mechanisms of the favorable effects of SGLT2i on syncope remains unclear. Based on the current researches, several hypotheses may help explain the observed associations. First, the favorable glycemic lowing effect of SGLT2i can slow down the progression of cardiac autonomic dysfunction (CAN) and therefore reduce the occurrence of VVS [16, 46, 47]. Second, independently of the hypoglycemic effect, SGLT2i may have the potential to directly improve or reverse CAN, thereby reducing the incidence of syncope [16]. Third, the protective role of SGLT2i in preventing syncope may be partially mediated by the favorable effects of SGLT2i in cardiovascular outcomes, such as heart failure, atherosclerotic cardiovascular diseases, atrial fibrillation and related thromboembolic events [36, 48-50]. Fourth, a poor kidney function has been associate with a higher risk of syncope [51]. Therefore, the established renoprotective effects of SGLT2i may also contribute to the favorable effect on syncope. In addition, future investigations are needed to explore the direct effects of SGLT2i on syncope.

### Study limitations

Several limitations should be noted for the present study. First, given the inherited nature of retrospective study, information bias from under-coding and coding errors are possible. However, the outcomes from the CDARS receive validation from review of medical records by physicians who have been responsible for the care of the patients, which could reduce the possibility of miscoding. And given the large sample size of this study, missing of a few syncope cases without hospitalization seems unlikely to significantly affect the primary results. Second, the lack of randomization cannot be fully replaced by propensity score matching. Indeed, it should be acknowledged that it is not always feasible to conduct an randomized clinical trial, such as in population with syncope, among which the underlying cause is often difficult to identify. Additionally, to testify our results, we have applied four PSM approaches and conducted comprehensive sensitivity analyses, which have yielded similar results. Third, out-of-hospital syncope was not included as an outcome event because the outcome was extracted exclusively from the medical record database. Fourth, our study only included four types of SGLT2i approved in China, including dapagliflozin, canagliflozin, empagliflozin, and ertugliflozin, which also represent the most common used SGLT2i in most of other countries. Other novel types of SGLT2i, such as ipragliflozin, tofogliflozin, luseogliflozin and sotaliflozin, are not approved in China yet and were not included in this study. Fifth, given the ICD-codes derived outcomes, the complexity of syncope, and the fact that the etiology of syncope is hard to identify, our study was unable to detailed classify the etiology of syncope and investigate the nature of syncope, which is also a common challenge existed in other large-scale syncope researches [52, 53]. Therefore, the mechanisms of the observed benefits of SGLT2i remain speculative which requires further investigations. Sixth, due to the nature of real-word study, the choice of performing echocardiography was at the discretion of the physicians. Therefore, echocardiographic data, such as left ventricular ejection fraction, was not available in the current study.

## Conclusions

SGLT2i therapy demonstrate robust clinical effectiveness in preventing new-onset syncope among patients with T2DM, regardless of gender, age, comorbidities, other medication history, and degree of glycemic control. Our findings suggest a promising future of SGLT2i in preventing incident syncope which have the potential to improve the preventive strategy of syncope.

## Data Availability

All data produced in the present study are available upon reasonable request to the authors.

## Authors contributions

X.G.: wrote the research project, and wrote and edited the manuscript; N.Z.: wrote and edited the manuscript; L.L., T.G., J.Z.: prepared the figures and performed statistical analyses; G.T.: edited and revised the manuscript; W.T.W., C.C., A.K.C.W.: contributed to the discussion and reviewed the manuscript; G.Y.H.L., T.L., J.Z.: reviewed the results and revised the manuscript. All authors have read and agreed to the published version of the manuscript.

## Ethical approval statement

This study was approved by the Institutional Review Board of the University of Hong Kong/Hospital Authority Hong Kong West Cluster (HKU/HA HKWC IRB) (UW-20-250) and complied with the Declaration of Helsinki.

## Declaration of competing interest

G.Y.H.L. is a consultant and speaker for BMS/Pfizer, Boehringer Ingelheim, Anthos and Daiichi-Sankyo. No fees are directly received personally. The remaining authors have no disclosures to report.

## Data availability statement

The datasets analysed during the current study are not publicly available as they contain personal data and the data custodians have not given permission. The data are available from the electronic health database in the computerized Clinical Management System of the Hong Kong Hospital Authority, subject to an application and research proposal meeting the ethical and governance requirements.

## Code availability

All analysis codes supporting the findings are available from the corresponding author upon reasonable request.

## Acknowledgments

All the authors, and colleagues the Hospital Authority for providing de-identified clinical data are equally thanked for their contributions to this research. Special thanks to the support of the National Natural Science Foundation of China (81970270, 82170327 to TL) and the Tianjin Key Medical Discipline (Specialty) Construction Project (TJYXZDXK-029A). Structural graphical abstract and Figure 1 are created with BioRender.com.

## Notes

### Competing Interest Statement

The authors have declared no competing interest.

### Funding Statement

The author(s) received no funding for the research, authorship, and/or publication of this article.

