## Supplementary material for "New-onset syncope in diabetic patients treated with sodium-glucose cotransporter-2 inhibitors versus dipeptidyl peptidase-4 inhibitors: A Chinese population-based cohort study": Table 1

**Table 1. Baseline and clinical characteristics** **of patients with SGLT2i vs. DPP4i use before and after propensity score matching (1:1).**

| **Characteristics** | **Before matching** | | | | **After matching** | | | |
| --- | --- | --- | --- | --- | --- | --- | --- | --- |
|  | **All (n=55370)** | **SGLT2i users (n=18751)** | **DPP4i users (n=36619)** | **SMD** | **All (n=37502)** | **SGLT2i users (n=18751)** | **DPP4i users (n=18751)** | **SMD** |
| ***Demographics*** |  |  |  |  |  |  |  |  |
| Male; n (%) | 27953 (50.48) | 10042 (53.55) | 17911 (48.91) | 0.09 | 19996 (53.31) | 10042 (53.55) | 9954 (53.08) | 0.01 |
| Baseline age; mean (SD) | 63.2 (12.8) | 57.7 (11.2) | 66.0 (12.7) | 0.69 | 58.9 (11.3) | 57.7 (11.2) | 60.1 (11.3) | 0.21* |
| 18-50; n (%) | 7696 (13.89) | 4094 (21.83) | 3602 (9.83) | 0.33 | 7257 (19.35) | 4094 (21.83) | 3163 (16.86) | 0.13 |
| 50-60; n (%) | 15192 (27.43) | 6702 (35.74) | 8490 (23.18) | 0.28 | 12907 (34.41) | 6702 (35.74) | 6205 (33.09) | 0.06 |
| 60-70; n (%) | 16253 (29.35) | 5638 (30.06) | 10615 (28.98) | 0.02 | 11687 (31.16) | 5638 (30.06) | 6049 (32.25) | 0.05 |
| 70-80; n (%) | 10093 (18.22) | 1896 (10.11) | 8197 (22.38) | 0.34 | 4434 (11.82) | 1896 (10.11) | 2538 (13.53) | 0.11 |
| >80; n (%) | 6142 (11.09) | 424 (2.26) | 5718 (15.61) | 0.48 | 1220 (3.25) | 424 (2.26) | 796 (4.24) | 0.11 |
| ***Prior comorbidities*** |  |  |  |  |  |  |  |  |
| Charlson’s comorbidity index; mean (SD) | 2.1 (1.5) | 1.5 (1.2) | 2.4 (1.6) | 0.60 | 1.6 (1.2) | 1.5 (1.2) | 1.7 (1.2) | 0.12 |
| Duration from earliest diabetes mellitus diagnosis date to baseline date (day); mean (SD) | 6.4 (4.9) | 6.35 (4.79) | 6.42 ( 4.95) | 0.01 | 6.3 (4.7) | 6.4 (4.8) | 6.2 (4.7) | 0.03 |
| Number of hospitalizations; mean (SD) | 1.2 (0.7) | 1.3 (1.0) | 1.1 (0.4) | 0.21 | 1.2 (0.8) | 1.3 (1.0) | 1.1 (0.5) | 0.19 |
| Diabetic retinopathy; n (%) | 3915 (7.07) | 1294 (6.90) | 2621 (7.15) | 0.01 | 2747 (7.32) | 1294 (6.90) | 1453 (7.74) | 0.03 |
| Diabetic nephropathy; n (%) | 704 (1.27) | 97 (0.51) | 607 (1.65) | 0.11 | 232 (0.61) | 97 (0.51) | 135 (0.71) | 0.03 |
| Diabetic neuropathy; n (%) | 1314 (2.37) | 447 (2.38) | 867 (2.36) | <0.01 | 934 (2.49) | 447 (2.38) | 487 (2.59) | 0.01 |
| Gout; n (%) | 1442 (2.60) | 357 (1.90) | 1085 (2.96) | 0.07 | 711 (1.89) | 357 (1.90) | 354 (1.88) | <0.01 |
| Heart failure; n (%) | 3040 (5.49) | 904 (4.82) | 2136 (5.83) | 0.05 | 1764 (4.70) | 904 (4.82) | 860 (4.58) | 0.01 |
| Hyperlipidemia; n (%) | 21961 (39.66) | 9333 (49.77) | 12628 (34.48) | 0.31 | 17756 (47.34) | 9333 (49.77) | 8423 (44.92) | 0.10 |
| Hypertension; n (%) | 17159 (30.98) | 6848 (36.52) | 10311 (28.15) | 0.18 | 13291 (35.44) | 6848 (36.52) | 6443 (34.36) | 0.05 |
| Hypoglycemia; n (%) | 436 (0.78) | 45 (0.23) | 391 (1.06) | 0.10 | 90 (0.23) | 45 (0.23) | 45 (0.23) | <0.01 |
| IHD with AMI; n (%) | 1312 (2.36) | 540 (2.87) | 772 (2.10) | 0.05 | 1072 (2.85) | 540 (2.87) | 532 (2.83) | <0.01 |
| IHD without AMI; n (%) | 3623 (6.54) | 1609 (8.58) | 2014 (5.49) | 0.12 | 3102 (8.27) | 1609 (8.58) | 1493 (7.96) | 0.02 |
| Liver diseases; n (%) | 2050 (3.70) | 838 (4.46) | 1212 (3.30) | 0.06 | 1536 (4.09) | 838 (4.46) | 698 (3.72) | 0.04 |
| COPD; n (%) | 846 (1.52) | 96 (0.51) | 750 (2.04) | 0.14 | 192 (0.51) | 96 (0.51) | 96 (0.51) | <0.01 |
| Peripheral vascular disease; n (%) | 414 (0.74) | 97 (0.51) | 317 (0.86) | 0.04 | 194 (0.51) | 97 (0.51) | 97 (0.51) | <0.01 |
| Renal diseases; n (%) | 985 (1.77) | 94 (0.50) | 891 (2.43) | 0.16 | 188 (0.50) | 94 (0.50) | 94 (0.50) | <0.01 |
| Stroke/TIA; n (%) | 1758 (3.17) | 471 (2.51) | 1287 (3.51) | 0.06 | 940 (2.50) | 471 (2.51) | 469 (2.50) | <0.01 |
| Atrial fibrillation; n (%) | 1329 (2.40) | 384 (2.04) | 945 (2.58) | 0.04 | 764 (2.03) | 384 (2.04) | 380 (2.02) | <0.01 |
| Anemia; n (%) | 2312 (4.17) | 459 (2.44) | 1853 (5.06) | 0.14 | 914 (2.43) | 459 (2.44) | 455 (2.42) | <0.01 |
| Cancer; n (%) | 1494 (2.69) | 389 (2.07) | 1105 (3.01) | 0.06 | 770 (2.05) | 389 (2.07) | 381 (2.03) | <0.01 |
| ***Drug exposure*** |  |  |  |  |  |  |  |  |
| SGLT2i frequency; mean (SD) | 7.2 (9.8) | 7.2 (9.8) | - | - | 7.2 (9.8) | 7.2 (9.8) | - | - |
| DPP4i frequency; mean (SD) | 5.2 (7.3) | - | 5.2 (7.3) | - | 4.5 (6.6) | - | 4.5 (6.6) | - |
| SGLT2i duration (days); mean (SD) | 527.1 (670.9) | 527.1 (670.9) | - | - | 527.1 (670.9) | 527.1 (670.9) | - | - |
| DPP4i duration (days); mean (SD) | 506.9 (286.1) | - | 506.9 (286.1) | - | 481.8 (283.9) | - | 481.8 (283.9) | - |
| Number of anti-diabetic drugs; mean (SD) | 8.0 (19.9) | 12.1 (29.0) | 5.9 (12.6) | 0.28 | 10.7 (26.2) | 12.1 (29.0) | 9.3 (23.0) | 0.11 |
| Metformin; n (%) | 49128 (88.72) | 17399 (92.78) | 31729 (86.64) | 0.20 | 34836 (92.89) | 17399 (92.78) | 17437 (92.99) | 0.01 |
| Sulphonylurea; n (%) | 42534 (76.81) | 13165 (70.20) | 29369 (80.20) | 0.23 | 26966 (71.90) | 13165 (70.20) | 13801 (73.60) | 0.08 |
| Insulin; n (%) | 27773 (50.15) | 9546 (50.90) | 18227 (49.77) | 0.02 | 19620 (52.31) | 9546 (50.90) | 10074 (53.72) | 0.06 |
| Acarbose; n (%) | 2035 (3.67) | 741 (3.95) | 1294 (3.53) | 0.02 | 1446 (3.85) | 741 (3.95) | 705 (3.75) | 0.01 |
| Thiozolidinedone; n (%) | 10356 (18.70) | 5204 (27.75) | 5152 (14.06) | 0.34 | 9293 (24.78) | 5204 (27.75) | 4089 (21.80) | 0.14 |
| Glucagon-like peptide-1 receptor agonists; n (%) | 2924 (5.28) | 1231 (6.56) | 1693 (4.62) | 0.08 | 2331 (6.21) | 1231 (6.56) | 1100 (5.86) | 0.03 |
| ACEI/ARBs; n (%) | 24315 (43.91) | 9215 (49.14) | 15100 (41.23) | 0.16 | 17451 (46.53) | 9215 (49.14) | 8236 (43.92) | 0.10 |
| Antihepatitis; n (%) | 912 (1.64) | 352 (1.87) | 560 (1.52) | 0.03 | 704 (1.87) | 352 (1.87) | 352 (1.87) | <0.01 |
| Anticoagulants; n (%) | 15885 (28.68) | 5991 (31.95) | 9894 (27.01) | 0.11 | 12510 (33.35) | 5991 (31.95) | 6519 (34.76) | 0.06 |
| Antiplatelets; n (%) | 16054 (28.99) | 5755 (30.69) | 10299 (28.12) | 0.06 | 11209 (29.88) | 5755 (30.69) | 5454 (29.08) | 0.04 |
| Lipid-lowering drugs; n (%) | 25241 (45.58) | 10543 (56.22) | 14698 (40.13) | 0.33 | 21565 (57.50) | 10543 (56.22) | 11022 (58.78) | 0.05 |
| Statins and fibrates; n (%) | 31248 (56.43) | 13811 (73.65) | 17437 (47.61) | 0.55 | 26843 (71.57) | 13811 (73.65) | 13032 (69.50) | 0.09 |
| Nitrates; n (%) | 6520 (11.77) | 2519 (13.43) | 4001 (10.92) | 0.08 | 4785 (12.75) | 2519 (13.43) | 2266 (12.08) | 0.04 |
| Non-steroidal anti-inflammatory drugs; n (%) | 13672 (24.69) | 5495 (29.30) | 8177 (22.32) | 0.16 | 10911 (29.09) | 5495 (29.30) | 5416 (28.88) | 0.01 |
| Diuretics; n (%) | 13673 (24.69) | 5587 (29.79) | 8086 (22.08) | 0.18 | 10570 (28.18) | 5587 (29.79) | 4983 (26.57) | 0.07 |
| Beta-blockers; n (%) | 11273 (20.35) | 4603 (24.54) | 6670 (18.21) | 0.15 | 8863 (23.63) | 4603 (24.54) | 4260 (22.71) | 0.04 |
| Calcium channel blockers; n (%) | 18820 (33.98) | 8172 (43.58) | 10648 (29.07) | 0.31 | 15871 (42.32) | 8172 (43.58) | 7699 (41.05) | 0.05 |
| Antihepatitis; n (%) | 1518 (2.74) | 352 (1.87) | 1166 (3.18) | 0.08 | 704 (1.87) | 352 (1.87) | 352 (1.87) | <0.01 |
| Anti-cancer drugs; n (%) | 4710 (8.50) | 868 (4.62) | 3842 (10.49) | 0.22 | 1780 (4.74) | 868 (4.62) | 912 (4.86) | 0.01 |
| Steroids/Corticosteroids; n (%) | 1607 (2.90) | 238 (1.26) | 1369 (3.73) | 0.16 | 476 (1.26) | 238 (1.26) | 238 (1.26) | <0.01 |
| ***Abbreviated MDRD*** ***(ml/min/1.73m^2^)*** |  |  |  |  |  |  |  |  |
| Abbreviated MDRD; mean (SD) | 79.3 (28.8)  n=45725 | 90.4 (24.0)  n=15756 | 73.5 (29.4)  n=29969 | 0.63 | 90.2 (26.6)  n=31601 | 90.4 (24.0)  n=15756 | 90.0 (28.9)  n=15845 | 0.01 |
| Most severe renal damage (<15); n (%) | 545.0 (0.98) | 16.0 (0.08) | 529.0 (1.44) | 0.16 | 38.0 (0.10) | 16.0 (0.08) | 22.0 (0.11) | 0.01 |
| Severe renal damage [15, 30); n (%) | 1406.0 (2.53) | 45.0 (0.23) | 1361.0 (3.71) | 0.25 | 188.0 (0.50) | 45.0 (0.23) | 143.0 (0.76) | 0.07 |
| Moderate to severe renal damage [30, 45); n (%) | 3843.0 (6.94) | 265.0 (1.41) | 3578.0 (9.77) | 0.37 | 902.0 (2.40) | 265.0 (1.41) | 637.0 (3.39) | 0.13 |
| Mild to moderate renal damage [45, 60); n (%) | 5642.0 (10.18) | 1028.0 (5.48) | 4614.0 (12.60) | 0.25 | 2597.0 (6.92) | 1028.0 (5.48) | 1569.0 (8.36) | 0.11 |
| Mild renal damage [60, 90]; n (%) | 17916.0 (32.35) | 6786.0 (36.19) | 11130.0 (30.39) | 0.12 | 14017.0 (37.37) | 6786.0 (36.19) | 7231.0 (38.56) | 0.05 |
| Chronic kidney disease (>90); n (%) | 16373.0 (29.57) | 7616.0 (40.61) | 8757.0 (23.91) | 0.36 | 13859.0 (36.95) | 7616.0 (40.61) | 6243.0 (33.29) | 0.15 |
| ***Complete blood counts; mean (SD)*** |  |  |  |  |  |  |  |  |
| Mean corpuscular volume (fL) | 87.1 (7.6)  n=27807 | 86.5 (7.2)  n=10574 | 87.5 (7.8)  n=17233 | 0.13 | 86.6 (7.7)  n=20492 | 86.5 (7.2)  n=10574 | 86.7 (8.2)  n=9918 | 0.02 |
| Eosinophil (x10^9^/L) | 0.2 (0.2)  n=22299 | 0.21 (0.19)  n=8239 | 0.21 (0.27)  n=14060 | 0.01 | 0.2 (0.2)  n=16631 | 0.21 (0.19)  n=8239 | 0.21 (0.2)  n=8392 | 0.03 |
| Lymphocyte (x10^9^/L) | 2.0 (0.9)  n=22323 | 2.2 (0.9)  n=8244 | 1.9 (0.9)  n=14079 | 0.31 | 2.1 (0.9)  n=16637 | 2.2 (0.9)  n=8244 | 2.0 (0.9)  n=8393 | 0.14 |
| Neutrophil (x10^9^/L) | 5.4 (2.8)  n=22323 | 5.1 (2.4)  n=8244 | 5.5 (3.0)  n=14079 | 0.13 | 5.3 (2.7)  n=16637 | 5.1 (2.4)  n=8244 | 5.5 (2.9)  n=8393 | 0.12 |
| White cell count (x10^9^/L) | 8.0 (3.0)  n=27817 | 7.97 (2.56)  n=10579 | 8.05 (3.25)  n=17238 | 0.02 | 8.0 (2.7)  n=20517 | 8.0 (2.6)  n=10579 | 8.1 (2.8)  n=9938 | 0.04 |
| Mean cell haemoglobin (pg) | 29.4 (3.0)  n=27807 | 29.1 (2.9)  n=10574 | 29.5 (3.1)  n=17233 | 0.15 | 29.2 (3.1)  n=20492 | 29.1 (2.9)  n=10574 | 29.3 (3.3)  n=9918 | 0.07 |
| Platelet (x10^9^/L) | 242.3 (72.7)  n=27814 | 248.4 (68.2)  n=10577 | 238.5 (75.1)  n=17237 | 0.14 | 247.1 (71.2)  n=20515 | 248.4 (68.2)  n=10577 | 245.8 (74.2)  n=9938 | 0.04 |
| Red cell count (x10^12^/L) | 4.5 (0.7)  n=27807 | 4.7 (0.6)  n=10574 | 4.3 (0.7)  n=17233 | 0.57 | 4.7 (0.6)  n=20492 | 4.7 (0.6)  n=10574 | 4.6 (0.7)  n=9918 | 0.16 |
| ***Liver and renal functions******; mean (SD)*** |  |  |  |  |  |  |  |  |
| Potassium (mmol/L) | 4.4 (0.5)  n=45578 | 4.3 (0.4)  n=15722 | 4.4 (0.5)  n=29856 | 0.14 | 4.3 (0.4)  n=31537 | 4.31 (0.43)  n=15722 | 4.26 (0.45)  n=15815 | 0.11 |
| Albumin (g/L) | 41.6 (4.0)  n=34813 | 42.5 (3.3)  n=13342 | 41.0 (4.3)  n=21471 | 0.38 | 42.3 (3.6)  n=25898 | 42.5 (3.3)  n=13342 | 42.1 (3.9)  n=12556 | 0.11 |
| Sodium (mmol/L) | 139.3 (3.0)  n=45604 | 139.2 (2.7)  n=15726 | 139.3 (3.1)  n=29878 | 0.05 | 139.3 (2.9)  n=31547 | 139.2 (2.7)  n=15726 | 139.5 (3.0)  n=15821 | 0.10 |
| Urea (mmol/L) | 6.6 (3.6)  n=45597 | 5.7 (2.0)  n=15723 | 7.1 (4.1)  n=29874 | 0.45 | 5.8 (2.2)  n=31556 | 5.7 (2.0)  n=15723 | 6.0 (2.3)  n=15833 | 0.12 |
| Protein (g/L) | 73.8 (5.6)  n=32721 | 74.4 (4.9)  n=12581 | 73.5 (5.9)  n=20140 | 0.17 | 74.4 (5.1)  n=24780 | 74.4 (4.92)  n=12581 | 74.44 (5.29)  n=12199 | 0.01 |
| Creatinine (umol/L) | 95.1 (78.2)  n=45725 | 77.1 (28.3)  n=15756 | 104.5 (93.0)  n=29969 | 0.40 | 79.7 (31.4)  n=31601 | 77.1 (28.3)  n=15756 | 82.2 (34.1)  n=15845 | 0.16 |
| Alkaline phosphatase (U/L) | 77.5 (33.5)  n=34925 | 73.9 (26.5)  n=13349 | 79.7 (37.0)  n=21576 | 0.18 | 76.6 (33.6)  n=25919 | 73.9 (26.5)  n=13349 | 79.5 (39.6)  n=12570 | 0.17 |
| Aspartate transaminase (U/L) | 27.8 (50.2)  n=13838 | 28.0 (28.4)  n=5341 | 27.7 (59.9)  n=8497 | 0.01 | 28.6 (29.6)  n=10764 | 28.0 (28.4)  n=5341 | 29.2 (30.7)  n=5423 | 0.04 |
| Alanine transaminase (U/L) | 28.7 (33.8)  n=29616 | 31.9 (29.0)  n=11418 | 26.6 (36.4)  n=18198 | 0.16 | 31.1 (30.3)  n=21765 | 31.9 (29.0)  n=11418 | 30.2 (31.8)  n=10347 | 0.06 |
| Bilirubin (umol/L) | 11.1 (7.1)  n=34745 | 11.3 (6.0)  n=13318 | 11.0 (7.7)  n=21427 | 0.05 | 11.1 (6.7)  n=25853 | 11.3 (6.0)  n=13318 | 10.8 (7.4)  n=12535 | 0.08 |
| ***Lipid profile and variability measures; mean (SD)*** |  |  |  |  |  |  |  |  |
| Triglyceride (mmol/L) | 1.7 (1.5)  n=42868 | 1.8 (1.7)  n=15120 | 1.7 (1.3)  n=27748 | 0.09 | 1.8 (1.5)  n=29991 | 1.8 (1.7)  n=15120 | 1.7 (1.3)  n=14871 | 0.06 |
| SD of triglyceride | 0.5 (1.0)  n=21932 | 0.5 (1.1)  n=9534 | 0.46 (0.84)  n=12398 | 0.04 | 0.5 (1.0)  n=17577 | 0.5 (1.1)  n=9534 | 0.51 (0.87)  n=8043 | 0.01 |
| Low-density lipoprotein (mmol/L) | 2.4 (0.8)  n=42172 | 2.4 (0.81)  n=14873 | 2.39 (0.81)  n=27299 | 0.01 | 2.4 (0.8)  n=29570 | 2.4 (0.81)  n=14873 | 2.42 (0.85)  n=14697 | 0.02 |
| SD of low-density lipoprotein | 0.4 (0.3)  n=21333 | 0.3 (0.3)  n=9285 | 0.4 (0.4)  n=12048 | 0.13 | 0.4 (0.3)  n=17161 | 0.3 (0.3)  n=9285 | 0.4 (0.4)  n=7876 | 0.17 |
| High-density lipoprotein (mmol/L) | 1.2 (0.3)  n=42809 | 1.18 (0.31)  n=15095 | 1.22 (0.34)  n=27714 | 0.14 | 1.2 (0.3)  n=29950 | 1.18 (0.31)  n=15095 | 1.22 (0.35)  n=14855 | 0.15 |
| SD of high-density lipoprotein | 0.1 (0.1)  n=21108 | 0.09 (0.08)  n=9147 | 0.11 (0.09)  n=11961 | 0.18 | 0.1 (0.1)  n=16842 | 0.09 (0.08)  n=9147 | 0.12 (0.1)  n=7695 | 0.27* |
| Total cholesterol (mmol/L) | 4.3 (1.0)  n=42909 | 4.4 (1.0)  n=15138 | 4.3 (1.0)  n=27771 | 0.01 | 4.4 (1.0)  n=30012 | 4.36 (1.01)  n=15138 | 4.39 (1.0)  n=14874 | 0.03 |
| SD of total cholesterol | 0.4 (0.4)  n=21961 | 0.4 (0.4)  n=9541 | 0.5 (0.4)  n=12420 | 0.13 | 0.4 (0.4)  n=17600 | 0.4 (0.4)  n=9541 | 0.5 (0.4)  n=8059 | 0.17 |
| ***Glucose tests and variability measures; mean (SD)*** |  |  |  |  |  |  |  |  |
| Hemoglobin A1C (%) | 8.0 (1.5)  n=44818 | 8.3 (1.6)  n=15508 | 7.9 (1.5)  n=29310 | 0.25 | 8.2 (1.6)  n=30935 | 8.3 (1.6)  n=15508 | 8.1 (1.6)  n=15427 | 0.11 |
| Mean hemoglobin A1C (%) | 8.0 (1.4)  n=43706 | 8.2 (1.4)  n=15088 | 7.8 (1.3)  n=28618 | 0.25 | 8.1 (1.5)  n=30221 | 8.2 (1.4)  n=15088 | 8.0 (1.5)  n=15133 | 0.11 |
| Variance of hemoglobin A1C | 0.8 (8.2)  n=31473 | 0.81 (10.34)  n=12472 | 0.81 (6.44)  n=19001 | <0.01 | 0.9 (9.7)  n=23614 | 0.8 (10.3)  n=12472 | 1.0 (8.9)  n=11142 | 0.02 |
| SD of hemoglobin A1C | 0.6 (0.7)  n=31473 | 0.56 (0.71)  n=12472 | 0.56 (0.7)  n=19001 | 0.01 | 0.6 (0.8)  n=23614 | 0.56 (0.71)  n=12472 | 0.59 (0.83)  n=11142 | 0.04 |
| Fasting glucose (mmol/L) | 8.9 (3.8)  n=40551 | 9.2 (3.6)  n=14293 | 8.7 (4.0)  n=26258 | 0.12 | 9.1 (3.9)  n=27830 | 9.2 (3.6)  n=14293 | 9.1 (4.2)  n=13537 | 0.03 |
| Mean fasting glucose (mmol/L) | 8.7 (2.9)  n=41411 | 9.0 (2.8)  n=14746 | 8.6 (3.0)  n=26665 | 0.13 | 8.9 (3.0)  n=28752 | 9.0 (2.8)  n=14746 | 8.9 (3.2)  n=14006 | 0.04 |
| Variance of fasting glucose | 8.8 (30.4)  n=25134 | 6.6 (22.1)  n=10596 | 10.3 (35.1)  n=14538 | 0.13 | 8.0 (26.2)  n=19382 | 6.6 (22.1)  n=10596 | 9.7 (30.4)  n=8786 | 0.12 |
| SD of fasting glucose | 2.0 (2.2)  n=25160 | 1.8 (1.8)  n=10612 | 2.1 (2.4)  n=14548 | 0.15 | 1.9 (2.0)  n=19408 | 1.8 (1.8)  n=10612 | 2.1 (2.3)  n=8796 | 0.15 |

* for SMD$\geq$0.2; ACEI: angiotensin-converting enzyme inhibitor; AMI: acute myocardial infarction; ARB: angiotensin receptor blocker; COPD: chronic obstructive pulmonary disease; DPP4i: dipeptidyl peptidase-4 inhibitor; IHD: ischemic heart disease; MDRD: Modification of Diet in Renal Disease; SD: standard deviation; SGLT2i: sodium glucose cotransporter-2 inhibitor; SMD: standardized mean difference; TIA: transient ischemic attack.
