## Supplementary material for "New-onset syncope in diabetic patients treated with sodium-glucose cotransporter-2 inhibitors versus dipeptidyl peptidase-4 inhibitors: A Chinese population-based cohort study": Table 2

**Table 2. Results of multivariate Cox analyses for primary and secondary outcomes in the matched cohort.**

| **Characteristics** | **New-onset Syncope** | | | **All-cause mortality** | | | **Cardiovascular mortality** | | | | | |
| --- | --- | --- | --- | --- | --- | --- | --- | --- | --- | --- | --- | --- |
|  | **aHR [95% CI]** | **P value** | | **aHR [95% CI]** | **P value** | | **aHR [95% CI]** | **P value** | | | | |
| **Model 1** | | | | | | | | | | | | |
| SGLT2i vs.DPP4i | 0.47 [0.40-0.54] | | <0.001 | 0.28 [0.26-0.31] | | <0.001 | 0.29 [0.23-0.37] | | | <0.001 | | |
| Dapagliflozin | 0.62 [0.52-0.73] | | <0.001 | 0.38 [0.33-0.43] | | <0.001 | 0.39 [0.29-0.53] | | | <0.001 | | |
| Empagliflozin | 0.62 [0.48-0.81] | | <0.001 | 0.34 [0.27-0.42] | | <0.001 | 0.30 [0.18-0.51] | | | <0.001 | | |
| Canagliflozin | 0.48 [0.37-0.61] | | <0.001 | 0.36 [0.30-0.44] | | <0.001 | 0.31 [0.20-0.49] | | | <0.001 | | |
| Ertugliflozin | 0.42 [0.29-0.61] | | <0.001 | 0.43 [0.34-0.54] | | <0.001 | 0.45 [0.26-0.76] | | | 0.003 | | |
| **Model 2** | | | | | | | | | | | | |
| SGLT2i vs.DPP4i | 0.02 [0--0.03] | | <0.001 | 0.29 [0.21-0.32] | | <0.001 | 0.28 [0.28-0.38] | | | | <0.001 | |
| Dapagliflozin | 0.48 [0.46-0.5] | | <0.001 | 0.33 [0.31-0.51] | | <0.001 | 0.39 [0.33-0.49] | | | | <0.001 | |
| Empagliflozin | 0.62 [0.55-0.74] | | 0.001 | 0.32 [0.34-0.45] | | <0.001 | 0.23 [0.19-0.52] | | | | <0.001 | |
| Canagliflozin | 0.65 [0.51-0.83] | | <0.001 | 0.34 [0.30-0.45] | | <0.001 | 0.3 [0.2-0.48] | | | | <0.001 | |
| Ertugliflozin | 0.45 [0.32-0.65] | | <0.001 | 0.43 [0.26-0.49] | | <0.001 | 0.35 [0.17-0.74] | | | | 0.003 | |
| **Model 3** | | | | | | | | | | | | |
| SGLT2i vs.DPP4i | 0.46 [0.40-0.53] | | <0.001 | 0.28 [0.25-0.31] | | <0.001 | 0.29 [0.23-0.36] | | | | <0.001 | |
| Dapagliflozin | 0.62 [0.52-0.73] | | <0.001 | 0.38 [0.33-0.43] | | <0.001 | 0.40 [0.30-0.53] | | | | <0.001 | |
| Empagliflozin | 0.62 [0.48-0.81] | | <0.001 | 0.34 [0.27-0.42] | | <0.001 | 0.30 [0.17-0.51] | | | | <0.001 | |
| Canagliflozin | 0.47 [0.36-0.60] | | <0.001 | 0.36 [0.30-0.43] | | <0.001 | 0.31 [0.20-0.49] | | | | <0.001 | |
| Ertugliflozin | 0.42 [0.29-0.62] | | <0.001 | 0.42 [0.33-0.54] | | <0.001 | 0.43 [0.25-0.74] | | | | 0.002 | |
| **Model 4** | | | | | | | | | | | | |
| SGLT2i vs.DPP4i | 0.43 [0.37-0.50] | | <0.001 | 0.30 [0.27-0.33] | | <0.001 | 0.34 [0.27-0.43] | | | | | <0.001 |
| Dapagliflozin | 0.67 [0.57-0.80] | | <0.001 | 0.40 [0.36-0.46] | | <0.001 | 0.44 [0.33-0.59] | | | | | <0.001 |
| Empagliflozin | 0.67 [0.51-0.87] | | 0.003 | 0.35 [0.28-0.44] | | <0.001 | 0.32 [0.19-0.54] | | | | | <0.001 |
| Canagliflozin | 0.42 [0.32-0.54] | | <0.001 | 0.38 [0.32-0.46] | | <0.001 | 0.37 [0.24-0.57] | | | | | <0.001 |
| Ertugliflozin | 0.35 [0.24-0.52] | | <0.001 | 0.46 [0.36-0.58] | | <0.001 | 0.53 [0.31-0.91] | | | | | 0.021 |
| **Model 5** | | | | | | | | | | | | |
| SGLT2i vs.DPP4i | 0.49 [0.41-0.57] | | <0.001 | 0.30 [0.26-0.34] | | <0.001 | 0.35 [0.26-0.46] | | <0.001 | | | |
| Dapagliflozin | 0.70 [0.58-0.85] | | <0.001 | 0.41 [0.36-0.48] | | <0.001 | 0.46 [0.32-0.65] | | <0.001 | | | |
| Empagliflozin | 0.79 [0.59-1.05] | | 0.100 | 0.32 [0.25-0.42] | | <0.001 | 0.30 [0.15-0.58] | | <0.001 | | | |
| Canagliflozin | 0.48 [0.36-0.63] | | <0.001 | 0.40 [0.33-0.50] | | <0.001 | 0.38 [0.22-0.64] | | <0.001 | | | |
| Ertugliflozin | 0.45 [0.30-0.68] | | <0.001 | 0.44 [0.33-0.59] | | <0.001 | 0.66 [0.36-1.18] | | 0.157 | | | |

Model 1 adjusted for significant demographics.

Model 2 adjusted for significant demographics, prior immune-mediated inflammatory diseases, and cancer.

Model 3 adjusted for significant demographics, and other past comorbidities.

Model 4 adjusted for significant demographics, past comorbidities, and non-SGLT2i /DPP4i medications.

Model 5 adjusted for significant demographics, past comorbidities, non-SGLT2i /DPP4i medications, abbreviated MDRD, fasting glucose, HbA1c, and duration from earliest diabetes mellitus date to initial drug exposure date.

aHR: adjusted hazard ratio; CI: confidence interval; DPP4i: dipeptidyl peptidase-4 inhibitor; HbA1c: hemoglobin A1c; MDRD: Modification of Diet in Renal Disease; SGLT2i : sodium glucose cotransporter-2 inhibitor.
