## Supplementary material for "New-onset syncope in diabetic patients treated with sodium-glucose cotransporter-2 inhibitors versus dipeptidyl peptidase-4 inhibitors: A Chinese population-based cohort study": Table 3

**Table 3. Subgroup analyses according to glucose measurements for the exposure effects of SGLT2i vs. DPP4i on new-onset syncope**

| **Subgroup** | **No. patients** | **No. SGLT2i (%)** | **New-onset syncope**  **in SGLT2i**  **(%)** | **No. DPP4i (%)** | **New-onset syncope**  **in DPP4i (%)** | **SGLT2i vs. DPP4i** | | **Dapagliflozin vs. DPP4i** | | **Empagliflozin vs. DPP4i** | | **Canagliflozin vs. DPP4i** | | **Ertugliflozin vs. DPP4i** | |
| --- | --- | --- | --- | --- | --- | --- | --- | --- | --- | --- | --- | --- | --- | --- | --- |
|  |  |  |  |  |  | **HR [95% CI]** | **P value** | **HR [95% CI]** | **P value** | **HR [95% CI]** | **P value** | **HR [95% CI]** | **P value** | **HR [95% CI]** | **P value** |
| ***Baseline HbA1c (%)*** | | | | | | | | | | | | | | | |
| <7.5 | 11425 | 5247 (45.93) | 93 (1.77) | 6178 (54.07) | 243 (3.93) | 0.44 [0.34-0.56] | <0.001 | 0.58 [0.43-0.78] | <0.001 | 0.62 [0.39-0.97] | 0.038 | 0.43  [0.28-0.67] | <0.001 | 0.51 [0.29-0.91] | 0.022 |
| 7.5-9 | 11973 | 6041 (50.46) | 93 (1.54) | 5932 (49.54) | 148 (2.49) | 0.61 [0.47-0.79] | <0.001 | 0.7 [0.52-0.95] | 0.022 | 0.9 [0.57-1.4] | 0.631 | 0.77  [0.51-1.16] | 0.211 | 0.59 [0.3-1.14] | 0.115 |
| >9 | 7537 | 4220 (55.99) | 68 (1.61) | 3317 (44.01) | 101 (3.04) | 0.51 [0.38-0.7] | <0.001 | 0.68 [0.48-0.95] | 0.026 | 0.83 [0.49-1.42] | 0.500 | 0.48  [0.26-0.85] | 0.013 | 0.24 [0.08-0.77] | 0.016 |
| ***Mean HbA1c (%)*** | | | | | | | | | | | | | | | |
| Q1 | 7533 | 3424 (45.45) | 58 (1.69) | 4109 (54.55) | 176 (4.28) | 0.38 [0.28-0.51] | <0.001 | 0.52 [0.36-0.76] | <0.001 | 0.61 [0.35-1.04] | 0.069 | 0.33  [0.18-0.59] | <0.001 | 0.42 [0.2-0.88] | 0.022 |
| Q2 | 7545 | 3465 (45.92) | 60 (1.73) | 4080 (54.08) | 129 (3.16) | 0.54 [0.4-0.73] | <0.001 | 0.65 [0.45-0.94] | 0.021 | 0.54 [0.28-1.05] | 0.070 | 0.75  [0.46-1.22] | 0.246 | 0.68 [0.34-1.39] | 0.291 |
| Q3 | 7585 | 4037 (53.22) | 58 (1.44) | 3548 (46.78) | 75 (2.11) | 0.67 [0.48-0.94] | 0.022 | 0.69 [0.46-1.03] | 0.071 | 1.21 [0.72-2.04] | 0.469 | 0.72  [0.41-1.25] | 0.245 | 0.53 [0.22-1.29] | 0.160 |
| Q4 | 7558 | 4162 (55.07) | 70 (1.68) | 3396 (44.93) | 105 (3.09) | 0.53 [0.39-0.71] | <0.001 | 0.71 [0.51-1.0] | 0.048 | 0.91 [0.55-1.51] | 0.727 | 0.5  [0.29-0.89] | 0.017 | 0.34 [0.13-0.91] | 0.032 |
| ***Variance of HbA1c*** | | | | | | | | | | | | | | | |
| Q1 | 5751 | 2862 (49.77) | 45 (1.57) | 2889 (50.23) | 114 (3.95) | 0.39 [0.28-0.55] | <0.001 | 0.48 [0.31-0.74] | <0.001 | 0.92 [0.54-1.57] | 0.761 | 0.43  [0.23-0.82] | 0.0104 | 0.19 [0.05-0.77] | 0.020 |
| Q2 | 5974 | 3258 (54.54) | 36 (1.1) | 2716 (45.46) | 82 (3.02) | 0.36 [0.24-0.53] | <0.001 | 0.51 [0.32-0.81] | 0.004 | 0.4 [0.16-0.98] | 0.046 | 0.48  [0.24-0.95] | 0.035 | 0.45 [0.17-1.23] | 0.119 |
| Q3 | 5927 | 3194 (53.89) | 50 (1.57) | 2733 (46.11) | 33 (1.21) | 1.28 [0.83-1.99] | 0.268 | 1.25 [0.8-1.95] | 0.333 | 1.12 [0.56-2.24] | 0.742 | 0.86  [0.44-1.66] | 0.644 | 1.07 [0.47-2.45] | 0.874 |
| Q4 | 5962 | 3158 (52.97) | 69 (2.18) | 2804 (47.03) | 131 (4.67) | 0.45 [0.34-0.6] | <0.001 | 0.55 [0.39-0.78] | <0.001 | 0.77 [0.46-1.29] | 0.325 | 0.52  [0.31-0.86] | 0.0114 | 0.55 [0.26-1.17] | 0.118 |
| ***CV of HbA1c*** | | | | | | | | | | | | | | | |
| Q1 | 5759 | 2951 (51.24) | 46 (1.56) | 2808 (48.76) | 91 (3.24) | 0.47 [0.33-0.67] | <0.001 | 0.55 [0.36-0.85] | 0.007 | 1.11 [0.66-1.88] | 0.687 | 0.56  [0.3-1.03] | 0.062 | 0.11 [0.02-0.78] | 0.027 |
| Q2 | 5759 | 3139 (54.51) | 32 (1.02) | 2620 (45.49) | 64 (2.44) | 0.41 [0.27-0.63] | <0.001 | 0.54 [0.33-0.9] | 0.017 | 0.42 [0.15-1.13] | 0.085 | 0.45  [0.21-0.98] | 0.044 | 0.86 [0.38-1.96] | 0.716 |
| Q3 | 5751 | 3133 (54.48) | 55 (1.76) | 2618 (45.52) | 45 (1.72) | 1.0 [0.68-1.49] | 0.987 | 1.04 [0.68-1.58] | 0.862 | 1.08 [0.58-2.03] | 0.799 | 0.83  [0.46-1.52] | 0.551 | 0.69 [0.28-1.7] | 0.418 |
| Q4 | 5769 | 3005 (52.09) | 62 (2.06) | 2764 (47.91) | 121 (4.38) | 0.46 [0.34-0.62] | <0.001 | 0.57 [0.4-0.81] | 0.002 | 0.7 [0.4-1.23] | 0.218 | 0.55  [0.32-0.93] | 0.025 | 0.64 [0.3-1.36] | 0.245 |
| ***Baseline fasting glucose (mmol/L)*** | | | | | | | | | | | | | | | |
| <5.6 | 6669 | 2807 (42.09) | 51 (1.82) | 3862 (57.91) | 119 (3.08) | 0.57 [0.41-0.8] | <0.001 | 0.74 [0.5-1.09] | 0.128 | 0.62 [0.32-1.22] | 0.166 | 0.49  [0.26-0.92] | 0.027 | 0.5 [0.2-1.21] | 0.122 |
| 5.6-6.9 | 5092 | 2382 (46.78) | 41 (1.72) | 2710 (53.22) | 126 (4.65) | 0.36 [0.25-0.51] | <0.001 | 0.53 [0.35-0.8] | 0.003 | 0.42 [0.2-0.9] | 0.026 | 0.29 [0.14-0.61] | 0.001 | 0.27 [0.08-0.83] | 0.023 |
| >6.9 | 19994 | 10679 (53.41) | 168 (1.57) | 9315 (46.59) | 259 (2.78) | 0.55 [0.46-0.67] | <0.001 | 0.65 [0.52-0.82] | <0.001 | 0.9 [0.65-1.25] | 0.534 | 0.69 [0.51-0.94] | 0.020 | 0.56 [0.35-0.91] | 0.020 |
| ***Mean fasting glucose (mmol/L)*** | | | | | | | | | | | | | | | |
| Q1 | 7111 | 3285 (46.2) | 54 (1.64) | 3826 (53.8) | 175 (4.57) | 0.35 [0.26-0.47] | <0.001 | 0.54 [0.38-0.77] | <0.001 | 0.31 [0.15-0.65] | 0.002 | 0.28 [0.14-0.54] | <0.001 | 0.33 [0.14-0.8] | 0.015 |
| Q2 | 7254 | 3645 (50.25) | 54 (1.48) | 3609 (49.75) | 70 (1.94) | 0.75 [0.53-1.08] | 0.121 | 0.67 [0.43-1.03] | 0.069 | 1.67 [1.02-2.76] | 0.043 | 0.84 [0.49-1.44] | 0.526 | 0.6 [0.25-1.47] | 0.264 |
| Q3 | 7195 | 3979 (55.3) | 65 (1.63) | 3216 (44.7) | 97 (3.02) | 0.53 [0.39-0.72] | <0.001 | 0.63 [0.43-0.91] | 0.013 | 0.66 [0.37-1.2] | 0.174 | 0.8  [0.5-1.29] | 0.363 | 0.68 [0.34-1.39] | 0.294 |
| Q4 | 7192 | 3837 (53.35) | 74 (1.93) | 3355 (46.65) | 120 (3.58) | 0.52 [0.39-0.7] | <0.001 | 0.67 [0.48-0.93] | 0.018 | 0.75 [0.45-1.26] | 0.280 | 0.57 [0.34-0.95] | 0.032 | 0.53 [0.25-1.14] | 0.104 |
| ***Variance of fasting glucose*** | | | | | | | | | | | | | | | |
| Q1 | 4844 | 2616 (54.0) | 28 (1.07) | 2228 (46.0) | 84 (3.77) | 0.28 [0.18-0.43] | <0.001 | 0.6 [0.38-0.95] | 0.028 | 0.32 [0.12-0.87] | 0.026 | 0.11 [0.03-0.43] | 0.002 | 0.12 [0.02-0.84] | 0.033 |
| Q2 | 4847 | 2888 (59.58) | 32 (1.11) | 1959 (40.42) | 67 (3.42) | 0.31 [0.21-0.48] | <0.001 | 0.48 [0.3-0.79] | 0.004 | 0.32 [0.12-0.88] | 0.028 | 0.38 [0.18-0.83] | 0.015 | 0.67 [0.27-1.65] | 0.388 |
| Q3 | 4844 | 2771 (57.2) | 55 (1.98) | 2073 (42.8) | 78 (3.76) | 0.52 [0.37-0.73] | <0.001 | 0.46 [0.3-0.7] | <0.001 | 1.35 [0.82-2.22] | 0.234 | 0.76 [0.45-1.31] | 0.326 | 0.48 [0.18-1.3] | 0.147 |
| Q4 | 4847 | 2321 (47.89) | 63 (2.71) | 2526 (52.11) | 110 (4.35) | 0.6 [0.44-0.82] | 0.001 | 0.72 [0.51-1.04] | 0.079 | 0.62 [0.33-1.17] | 0.140 | 0.6 [0.35-1.03] | 0.066 | 0.93 [0.48-1.83] | 0.843 |
| ***CV of fasting glucose*** | | | | | | | | | | | | | | | |
| Q1 | 4723 | 2573 (54.48) | 29 (1.13) | 2150 (45.52) | 78 (3.63) | 0.3 [0.2-0.47] | <0.001 | 0.63 [0.4-0.99] | 0.047 | 0.42 [0.17-1.02] | 0.056 | 0.17 [0.05-0.52] | 0.002 | 0.12 [0.02-0.89] | 0.038 |
| Q2 | 4722 | 2852 (60.4) | 32 (1.12) | 1870 (39.6) | 54 (2.89) | 0.38 [0.25-0.59] | <0.001 | 0.54 [0.32-0.9] | 0.018 | 0.55 [0.24-1.26] | 0.157 | 0.52 [0.25-1.07] | 0.075 | 0.78 [0.32-1.92] | 0.588 |
| Q3 | 4716 | 2680 (56.83) | 49 (1.83) | 2036 (43.17) | 106 (5.21) | 0.34 [0.24-0.48] | <0.001 | 0.4 [0.26-0.61] | <0.001 | 0.54 [0.27-1.05] | 0.07 | 0.42 [0.22-0.79] | 0.007 | 0.6 [0.27-1.36] | 0.219 |
| Q4 | 4730 | 2252 (47.61) | 60 (2.66) | 2478 (52.39) | 67 (2.7) | 0.96 [0.68-1.36] | 0.808 | 0.91 [0.61-1.35] | 0.622 | 1.43 [0.84-2.46] | 0.191 | 0.86 [0.49-1.49] | 0.582 | 1.03 [0.48-2.21] | 0.935 |

CI: confidence interval; CV: coefficient of variation; DPP4i: dipeptidyl peptidase-4 inhibitor; HbA1c: hemoglobin A1c; HR: adjusted hazard ratio; SGLT2i: sodium-glucose cotransporter-2 inhibitor.
