## Supplementary Appendix for "New-onset syncope in diabetic patients treated with sodium-glucose cotransporter-2 inhibitors versus dipeptidyl peptidase-4 inhibitors: A Chinese population-based cohort study"

[Table S1. The](#_Toc10643) *[International Classification of Diseases](#_Toc10643)* [codes](#_Toc10643) *[(Ninth Revision, ICD-9](#_Toc10643)*[) for disease diagnosis.	9](#_Toc10643)

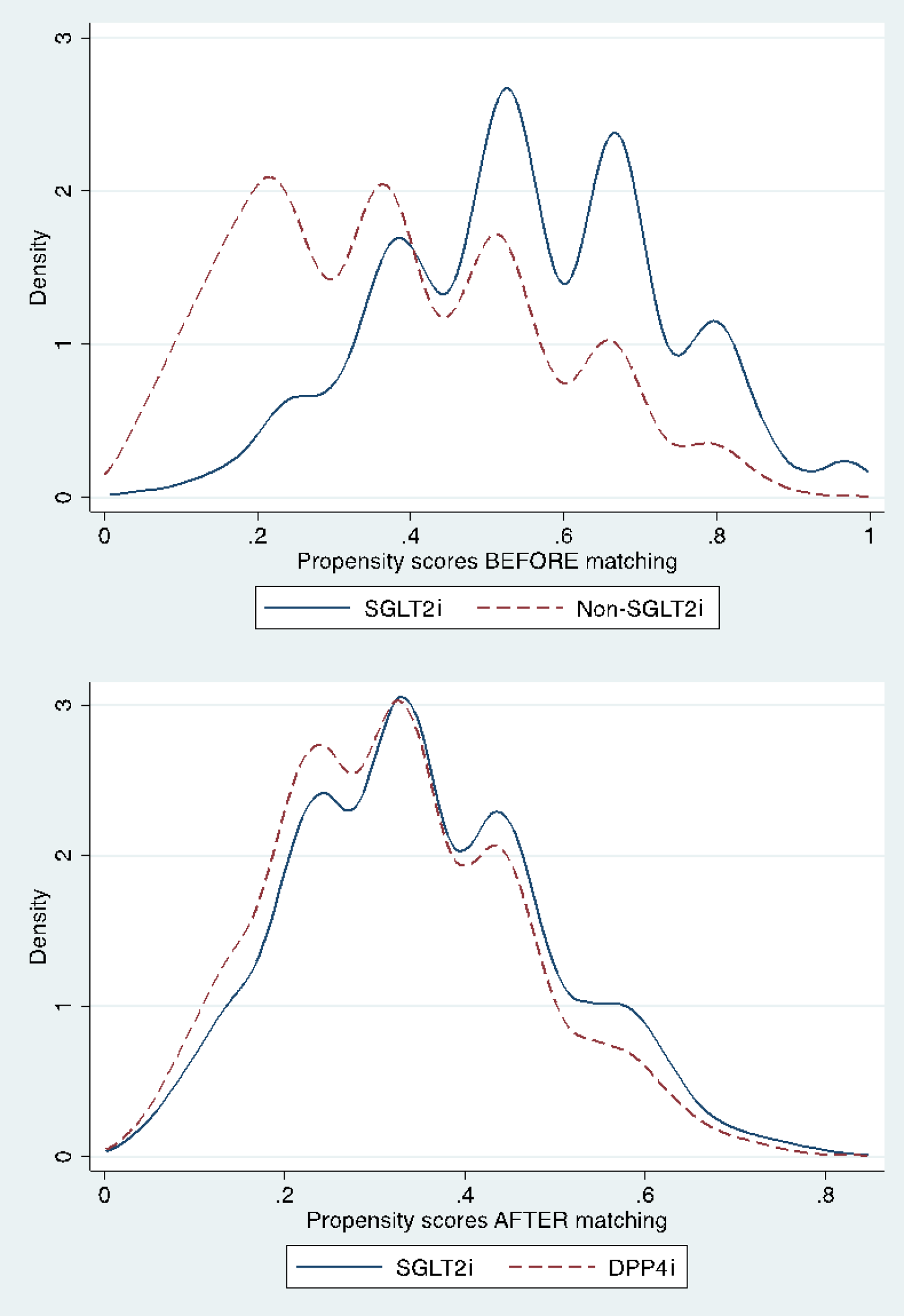

Figure S1. Propensity score matching for SGLT2i use versus DPP4i use before and after 1:1 matching with nearest neighbour search strategy (caliper=0.5).

DPP4i: dipeptidyl peptidase-4 inhibitor; SGLT2i: sodium glucose cotransporter-2 inhibitor.

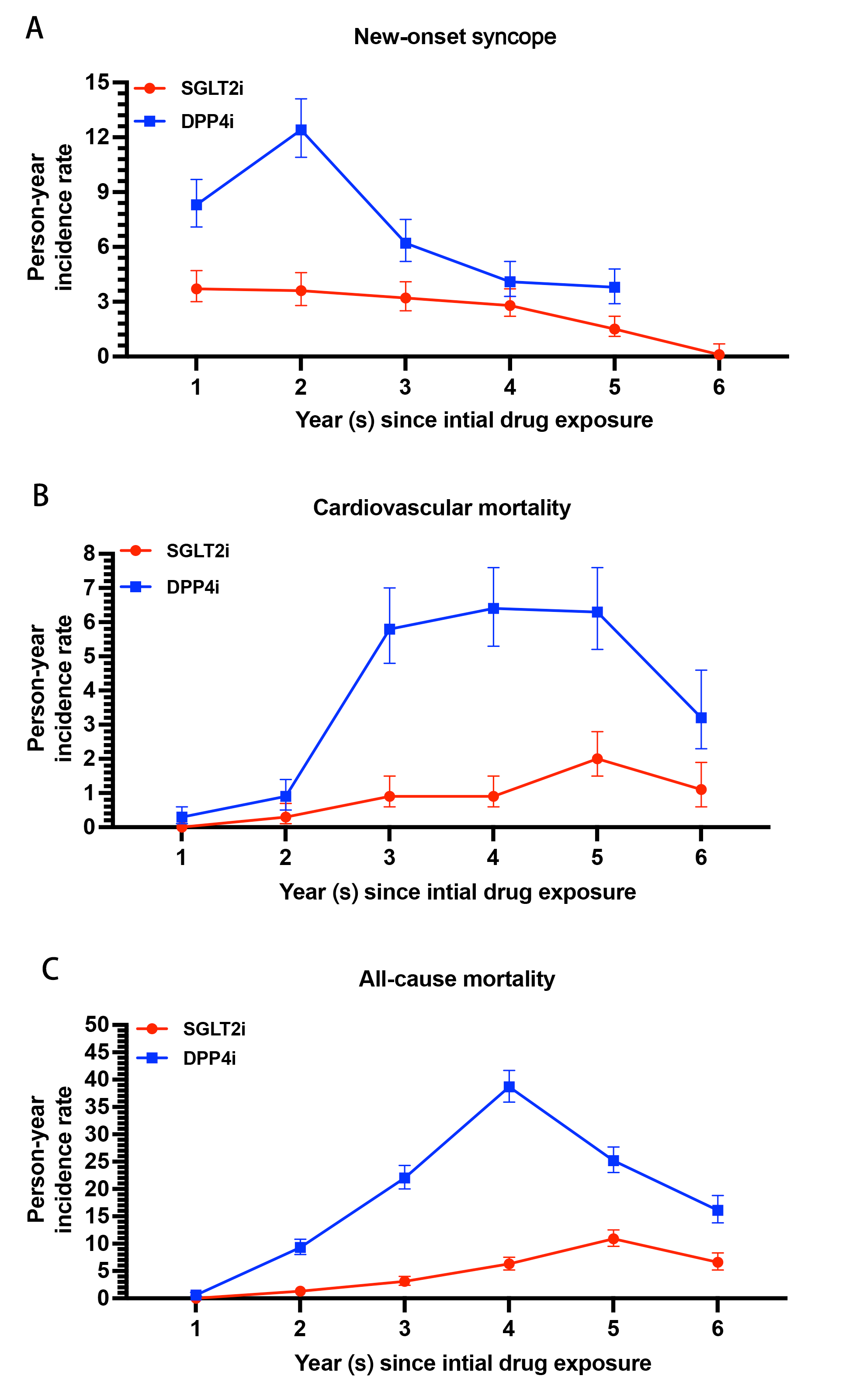

Figure S2. The annual person-year incidence ratios of (A) new-onset syncope, (B) cardiovascular mortality, and (C) all-cause mortality in the matched cohort.

DPP4i: dipeptidyl peptidase-4 inhibitor; SGLT2i: sodium glucose cotransporter-2 inhibitor.

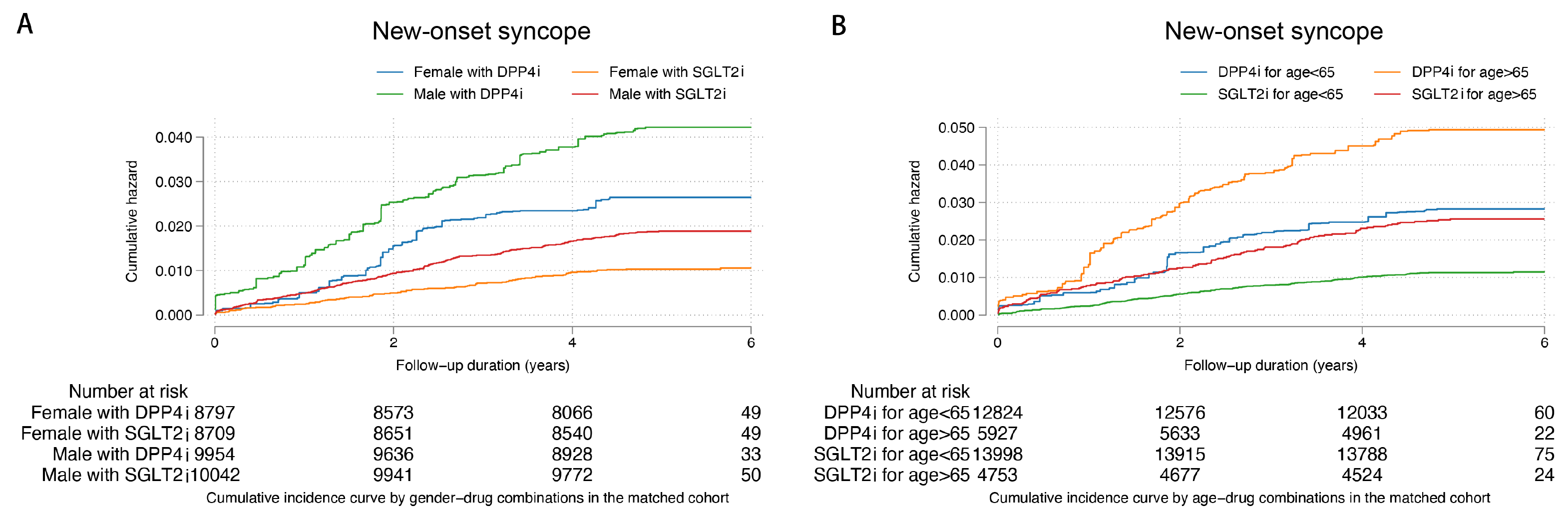

Figure S3. Cumulative incidence curves for new-onset syncope stratified by drug exposure effects of SGLT2i and DPP4i in the subgroup analyses by (A) gender and (B) age.

DPP4i: dipeptidyl peptidase-4 inhibitor; SGLT2i: sodium glucose cotransporter-2 inhibitor.

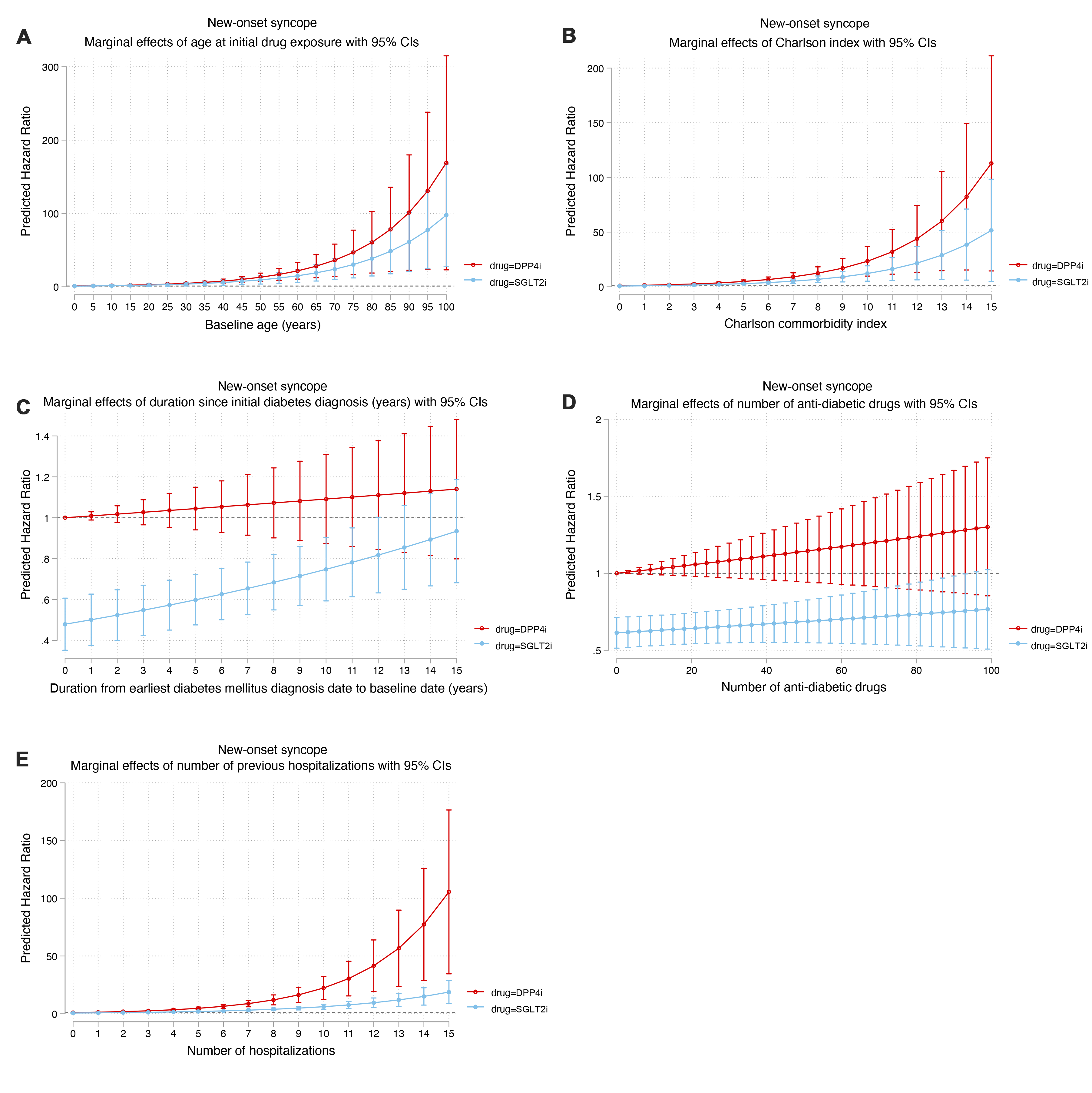

Figure S4. Marginal effects plots for (A) age at initial drug exposure, (B) Charlson index, (C) duration since initial diabetes diagnosis, (D)Number of anti-diabetic drugs, (E) number of hospitalizations on new-onset syncope stratified by drug exposure effects of SGLT2i and DPP4i in the matched cohort (1:1).

CI: confidence interval; DPP4i: dipeptidyl peptidase-4 inhibitor; SGLT2i: sodium glucose cotransporter-2 inhibitor.

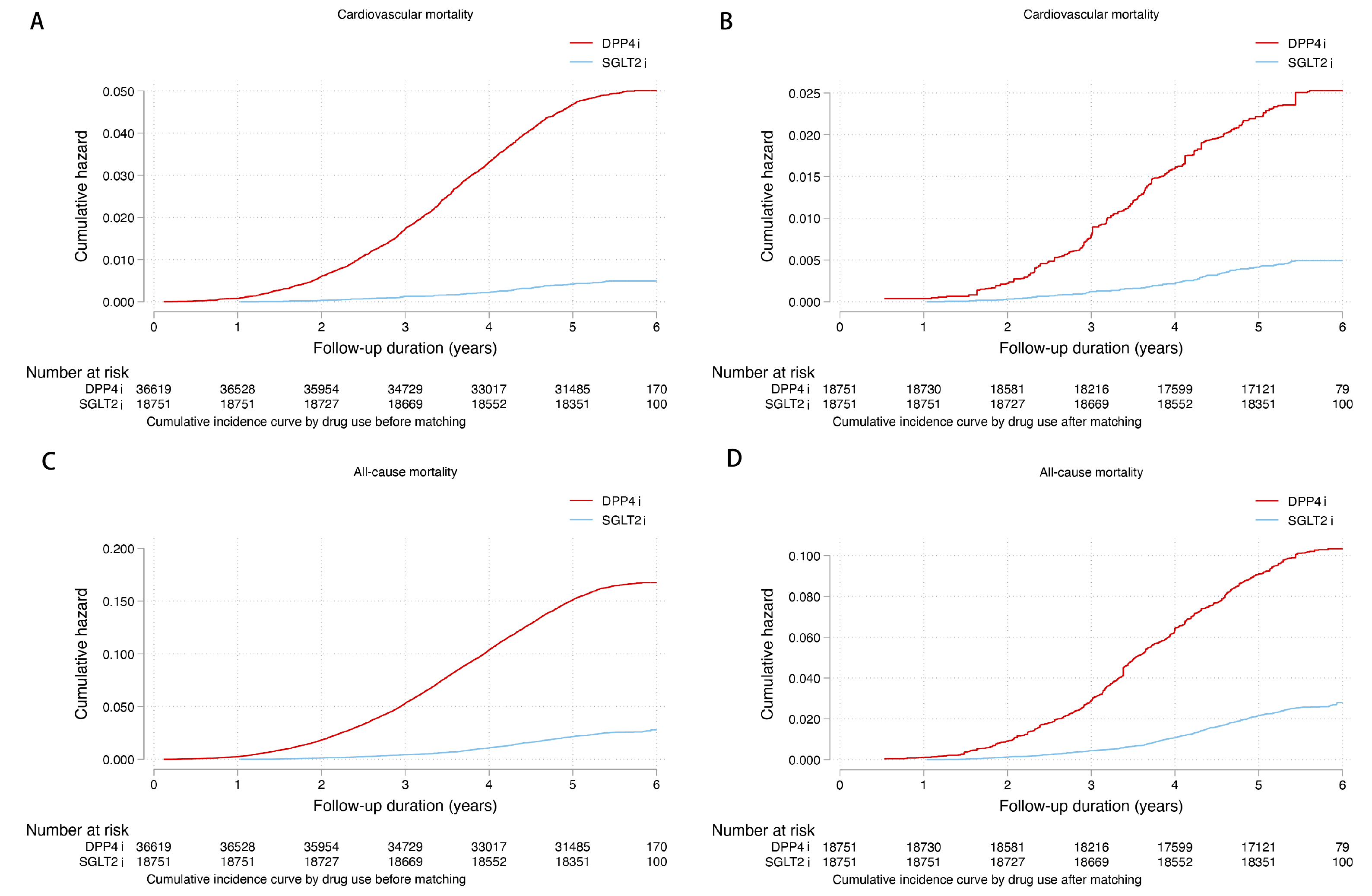

Figure S5. Cumulative incidence curves for (A) cardiovascular mortality before matching; (B) cardiovascular mortality after matching; (C) all-cause mortality before matching; (D) all-cause mortality after matching stratified by drug exposure effects of SGLT2i and DPP4i

DPP4i: dipeptidyl peptidase-4 inhibitor; SGLT2i: sodium glucose cotransporter-2 inhibitor.

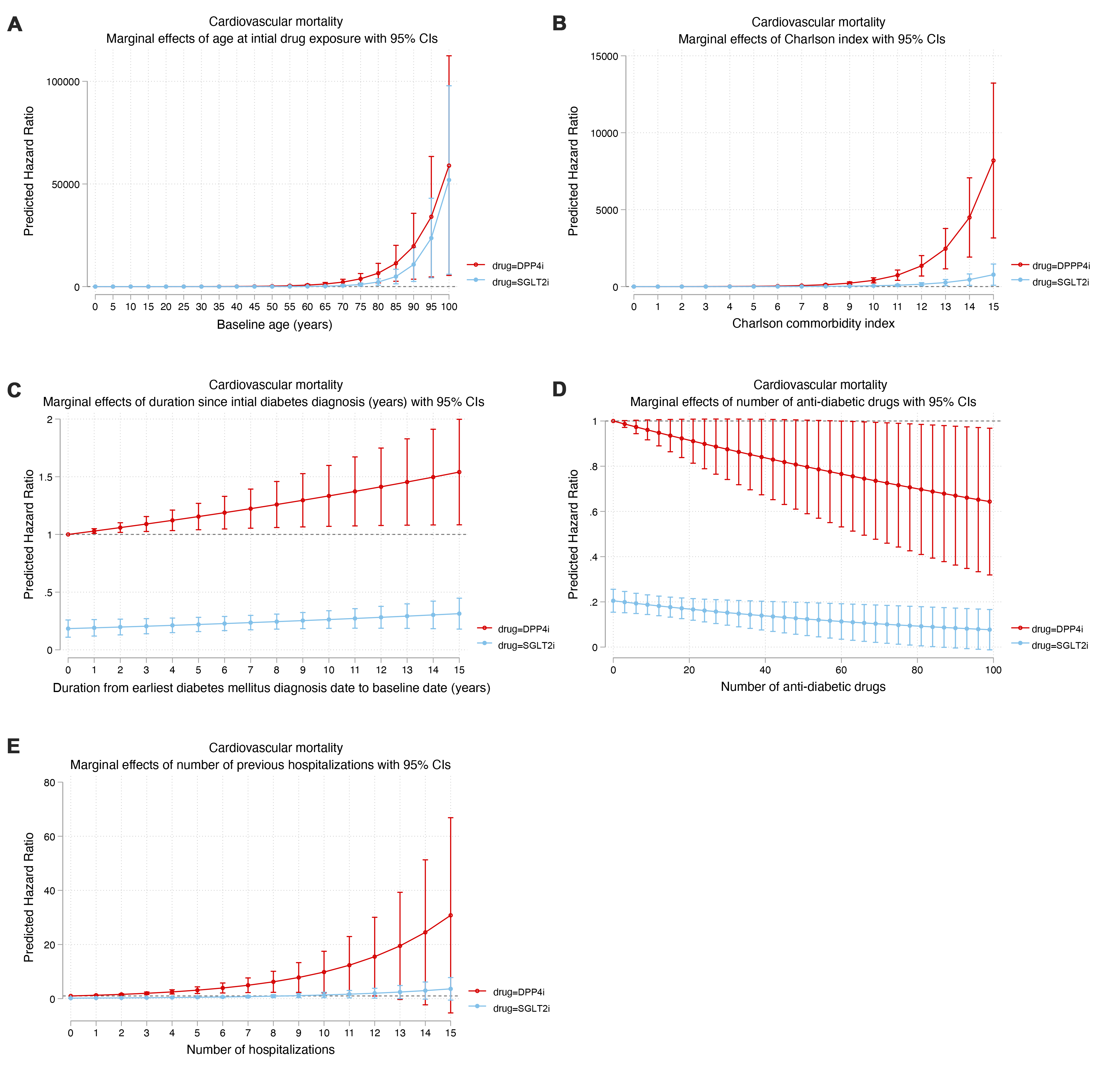

Figure S6. Marginal effects plots for (A) age at initial drug exposure, (B) Charlson index, (C) duration since initial diabetes diagnosis, (D)Number of anti-diabetic drugs, (E) number of hospitalizations on cardiovascular mortality stratified by drug exposure effects of SGLT2i and DPP4i in the matched cohort (1:1).

CI: confidence interval; DPP4i: dipeptidyl peptidase-4 inhibitor; SGLT2i: sodium glucose cotransporter-2 inhibitor.

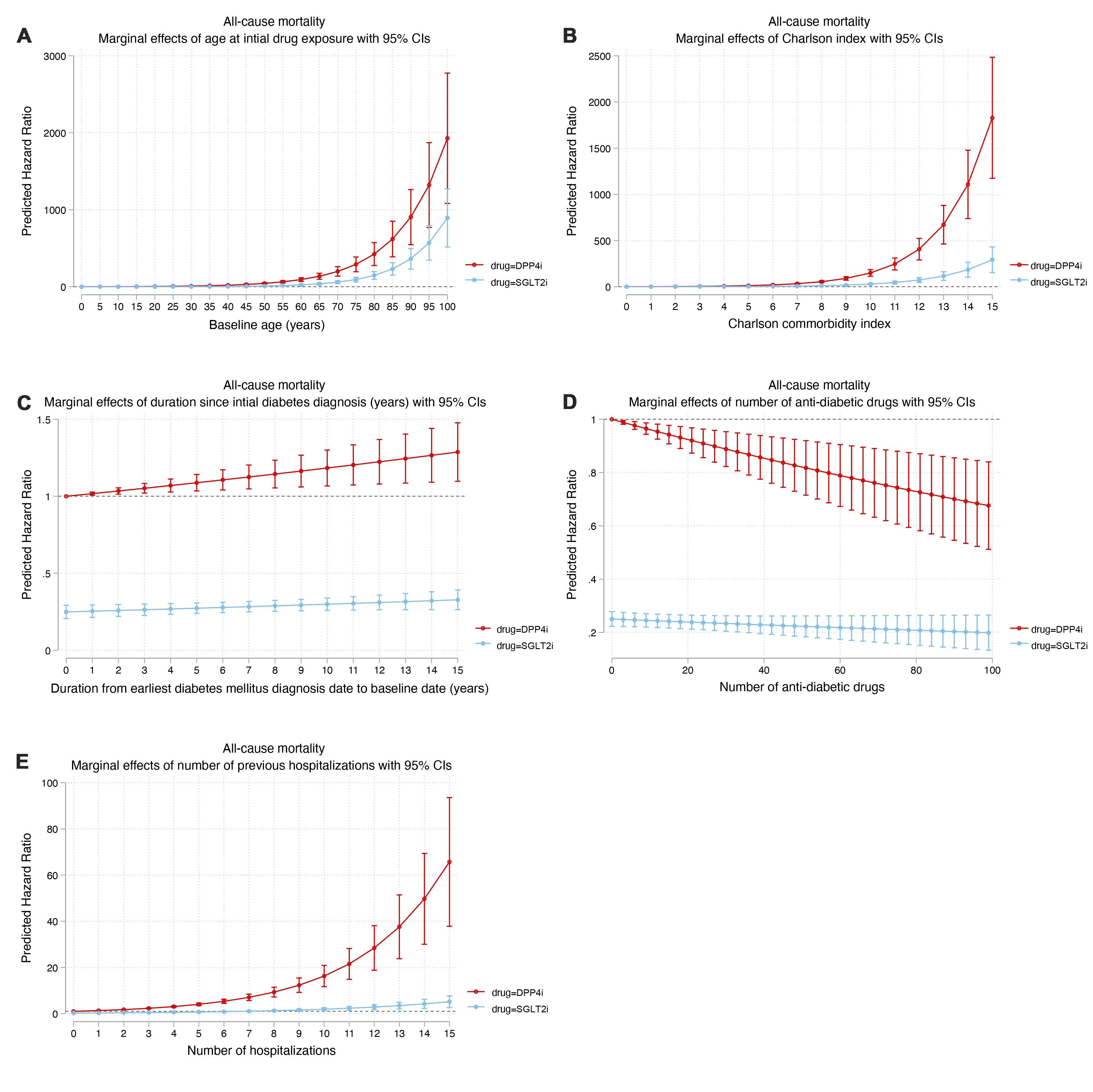

Figure S7. Marginal effects plots for (A) age at initial drug exposure, (B) Charlson index, (C) duration since initial diabetes diagnosis, (D)Number of anti-diabetic drugs, (E) number of hospitalizations on all-cause mortality stratified by drug exposure effects of SGLT2i and DPP4i in the matched cohort (1:1).

CI: confidence interval; DPP4i: dipeptidyl peptidase-4 inhibitor; SGLT2i: sodium glucose cotransporter-2 inhibitor.

Table S1. The *International Classification of Diseases* codes *(Ninth Revision, ICD-9*) for disease diagnosis.

| **Comorbidities** | **ICD-9 codes** |
| --- | --- |
| Syncope | 780.2 |
| Gout | 274 274.01 274.02 274.03 274.1 274.11 274.19 274.8 274.81 274.82 274.89 274.9 |
| Heart failure | 428 428 428.1 428.2 428.2 428.21 428.22 428.23 428.3 428.3 428.31 428.32 428.33 428.4 428.4 428.41 428.42 428.43 428.9 398.91 402.01 402.11 402.91 404.01 404.03 404.11 404.13 404.91 404.93 |
| Hyperlipidaemia | 272 272 272.1 272.2 272.3 |
| Hypertension | 401 401.1 401.9 402 402.01 402.1 402.11 402.9 402.91 403 403.01 403.1 403.11 403.9 403.91 404 404.01 404.02 404.03 404.1 404.11 404.12 404.13 404.9 404.91 404.92 404.93 405 405.01 405.09 405.1 405.11 405.19 405.9 405.91 405.99 437.2 |
| Hypoglycemia | 251.2 |
| IHD with AMI | 410.01 410.02 410.1 410.11 410.12 410.2 410.21 410.22 410.3 410.31 410.32 410.4 410.41 410.42 410.5 410.51 410.52 410.6 410.61 410.62 410.7 410.71 410.72 410.8 410.81 410.82 410.9 410.91 410.92 411 411.1 411.8 411.81 411.89 413 413.1 413.9 414 414.01 414.02 414.03 414.04 414.05 414.06 414.07 414.1 414.11 414.12 414.19 414.2 414.3 414.4 414.8 414.9 410 412 |
| IHD without AMI | 411 411.1 411.8 411.81 411.89 413 413.1 413.9 414 414.01 414.02 414.03 414.04 414.05 414.06 414.07 414.1 414.11 414.12 414.19 414.2 414.3 414.4 414.8 414.9 410 412 |
| Liver diseases | 456 456.1 456.2 572.2 572.3 572.4 572.8 571.4 571.5 571.6 |
| COPD | 490 491 492 493 494 495 496 491.1 491.2 491.21 491.22 491.8 491.9 492.8 493.01 493.02 493.1 493.11 493.12 493.2 493.21 493.22 493.8 493.81 493.82 493.9 493.91 493.92 494.1 495.1 495.2 495.3 495.4 495.5 495.6 495.7 495.8 495.9 |
| Peripheral vascular disease | 250.7 443.9 443 443.1 443.2 443.21 443.22 443.23 443.24 443.29 443.8 443.81 443.82 443.89 441 443.9 785.4 V43.4 |
| Renal diseases | 582 582 582.1 582.2 582.4 582.8 582.81 582.89 582.9 583 583 583.1 583.2 583.4 583.6 583.7 585 585.1 585.2 585.3 585.4 585.5 585.6 585.9 586 588 588 588.1 588.8 588.81 588.89 588.9 |
| Stroke/TIA | 435 435.1 435.2 435.3 435.8 435.9 433.81 433.91 434 436 437 437.1 433.31 433.01 434.01 434.1 434.11 434.9 434.91 437.2 437.3 437.4 437.5 437.6 437.7 437.8 437.9 430 431 432 432.1 432.9 |
| Atrial fibrillation | 427.31 429.4 |
| Anemia | 280 280.1 280.8 280.9 281 281.1 281.2 281.3 281.4 281.8 281.9 282.2 282.3 282.8 282.9 283 283.1 283.11 283.19 283.2 283.9 284 284.01 284.09 284.1 284.11 284.12 284.19 284.81 284.9 285 285.1 285.2 285.21 285.22 285.29 285.3 285.8 285.9 |
| Immune-mediated inflammatory diseases | 714 720 720.8 720.9 696 696.1 696.8 691.8 695.4 373.34 695.4 710.1 710.2 710.3 710.4 555 555.1 555.2 555.9 556.1 556.2 556.3 556.4 556.5 556.6 556.7 556.8 556.9 340 493 493 493.91 493.11 493.2 493.2 493.02 493.1 493.92 493.21 |
| Cancer | 140 140.1 140.3 140.4 140.5 140.6 140.8 140.9 141 141.1 141.2 141.3 141.4 141.5 141.6 141.8 141.9 142 142.1 142.2 142.8 142.9 143 143.1 143.8 143.9 144 144.1 144.8 144.9 145 145.1 145.2 145.3 145.4 145.5 145.6 145.8 145.9 146 146.1 146.2 146.3 146.4 146.5 146.6 146.7 146.8 146.9 147 147.1 147.2 147.3 147.8 147.9 148 148.1 148.2 148.3 148.8 148.9 149 149.1 149.8 149.9 150 150.1 150.2 150.3 150.4 150.5 150.8 150.9 151 151.1 151.2 151.3 151.4 151.5 151.6 151.8 151.9 152 152.1 152.2 152.3 152.8 152.9 153 153.1 153.2 153.3 153.4 153.5 153.6 153.7 153.8 153.9 154 154.1 154.2 154.3 154.8 155 155.1 155.2 156 156.1 156.2 156.8 156.9 157 157.1 157.2 157.3 157.4 157.8 157.9 158 158.8 158.9 159 159.1 159.8 159.9 160 160.1 160.2 160.3 160.4 160.5 160.8 160.9 161 161.1 161.2 161.3 161.8 161.9 162 162.2 162.3 162.4 162.5 162.8 162.9 163 163.1 163.8 163.9 164 164.1 164.2 164.3 164.8 164.9 165 165.8 165.9 170 170.1 170.2 170.3 170.4 170.5 170.6 170.7 170.8 170.9 171 171.2 171.3 171.4 171.5 171.6 171.7 171.8 171.9 172 172.1 172.2 172.3 172.4 172.5 172.6 172.7 172.8 172.9 173 173.01 173.02 173.09 173.1 173.11 173.12 173.19 173.2 173.21 173.22 173.29 173.3 173.31 173.32 173.39 173.4 173.41 173.42 173.49 173.5 173.51 173.52 173.59 173.6 173.61 173.62 173.69 173.7 173.71 173.72 173.79 173.8 173.81 173.82 173.89 173.9 173.91 173.92 173.99 174 174.1 174.2 174.3 174.4 174.5 174.6 174.8 174.9 175 175.9 176 176.1 176.2 176.3 176.4 176.5 176.8 176.9 179 180 180.1 180.8 180.9 181 182 182.1 182.8 183 183.2 183.3 183.4 183.5 183.8 183.9 184 184.1 184.2 184.3 184.4 184.8 184.9 185 186 186.9 187 187.1 187.2 187.3 187.4 187.5 187.6 187.7 187.8 187.9 188 188.1 188.2 188.3 188.4 188.5 188.6 188.7 188.8 188.9 189 189.1 189.2 189.3 189.4 189.8 189.9 190 190.1 190.2 190.3 190.4 190.5 190.6 190.7 190.8 190.9 191 191.1 191.2 191.3 191.4 191.5 191.6 191.7 191.8 191.9 192 192.1 192.2 192.3 192.8 192.9 193 194 194.1 194.3 194.4 194.5 194.6 194.8 194.9 195 195.1 195.2 195.3 195.4 195.5 195.8 200 200.01 200.02 200.03 200.04 200.05 200.06 200.07 200.08 200.1 200.11 200.12 200.13 200.14 200.15 200.16 200.17 200.18 200.2 200.21 200.22 200.23 200.24 200.25 200.26 200.27 200.28 200.3 200.31 200.32 200.33 200.34 200.35 200.36 200.37 200.38 200.4 200.41 200.42 200.43 200.44 200.45 200.46 200.47 200.48 200.5 200.51 200.52 200.53 200.54 200.55 200.56 200.57 200.58 200.6 200.61 200.62 200.63 200.64 200.65 200.66 200.67 200.68 200.7 200.71 200.72 200.73 200.74 200.75 200.76 200.77 200.78 200.8 200.81 200.82 200.83 200.84 200.85 200.86 200.87 200.88 201 201.01 201.02 201.03 201.04 201.05 201.06 201.07 201.08 201.1 201.11 201.12 201.13 201.14 201.15 201.16 201.17 201.18 201.2 201.21 201.22 201.23 201.24 201.25 201.26 201.27 201.28 201.4 201.41 201.42 201.43 201.44 201.45 201.46 201.47 201.48 201.5 201.51 201.52 201.53 201.54 201.55 201.56 201.57 201.58 201.6 201.61 201.62 201.63 201.64 201.65 201.66 201.67 201.68 201.7 201.71 201.72 201.73 201.74 201.75 201.76 201.77 201.78 201.9 201.91 201.92 201.93 201.94 201.95 201.96 201.97 201.98 202 202.01 202.02 202.03 202.04 202.05 202.06 202.07 202.08 202.1 202.11 202.12 202.13 202.14 202.15 202.16 202.17 202.18 202.2 202.21 202.22 202.23 202.24 202.25 202.26 202.27 202.28 202.3 202.31 202.32 202.33 202.34 202.35 202.36 202.37 202.38 202.4 202.41 202.42 202.43 202.44 202.45 202.46 202.47 202.48 202.5 202.51 202.52 202.53 202.54 202.55 202.56 202.57 202.58 202.6 202.61 202.62 202.63 202.64 202.65 202.66 202.67 202.68 202.7 202.71 202.72 202.73 202.74 202.75 202.76 202.77 202.78 202.8 202.81 202.82 202.83 202.84 202.85 202.86 202.87 202.88 202.9 202.91 202.92 202.93 202.94 202.95 202.96 202.97 202.98 203 203.01 203.02 203.1 203.11 203.12 203.8 203.81 203.82 204 204.01 204.02 204.1 204.11 204.12 204.2 204.21 204.22 204.8 204.81 204.82 204.9 204.91 204.92 205 205.01 205.02 205.1 205.11 205.12 205.2 205.21 205.22 205.3 205.31 205.32 205.8 205.81 205.82 205.9 205.91 205.92 206 206.01 206.02 206.1 206.11 206.12 206.2 206.21 206.22 206.8 206.81 206.82 206.9 206.91 206.92 207 207.01 207.02 207.1 207.11 207.12 207.2 207.21 207.22 207.8 207.81 207.82 208 208.01 208.02 208.1 208.11 208.12 208.2 208.21 208.22 208.8 208.81 208.82 208.9 208.91 208.92 196 196.1 196.2 196.3 196.5 196.6 196.8 196.9 197 197.1 197.2 197.3 197.4 197.5 197.6 197.7 197.8 198 198.1 198.2 198.3 198.4 198.5 198.6 198.7 198.8 198.81 198.82 198.89 199 199.1 |

AMI: acute myocardial infarction; COPD: chronic obstructive pulmonary disease; IHD: ischemic heart disease; TIA: transient ischemic attack.

Table S2. Baseline and clinical characteristics of patients with/without new-onset syncope outcome before and after propensity score matching (1:1).

| **Characteristics** | **Before matching** | | | **After matching** | | |
| --- | --- | --- | --- | --- | --- | --- |
|  | **With new-onset syncope (N=1374)** | **Without new-onset syncope (N=53996)** | **SMD** | **new-onset syncope (N=907)** | **No new-onset syncope (N=36595)** | **SMD** |
| ***Demographics*** |  |  |  |  |  |  |
| Male; n (%) | 855 (62.22) | 27098 (50.18) | 0.24* | 591 (65.15) | 19405 (53.02) | 0.25* |
| Baseline age; mean (SD) | 69.7 (11.6) | 63.0 (12.8) | 0.55* | 63.6 (11.2) | 58.8 (11.3) | 0.43* |
| 18-50; n (%) | 60 (4.36) | 7636 (14.14) | 0.34* | 96 (10.58) | 7161 (19.56) | 0.25* |
| 50-60; n (%) | 234 (17.03) | 14958 (27.70) | 0.26* | 282 (31.09) | 12625 (34.49) | 0.07 |
| 60-70; n (%) | 358 (26.05) | 15895 (29.43) | 0.08 | 263 (28.99) | 11424 (31.21) | 0.05 |
| 70-80; n (%) | 418 (30.42) | 9675 (17.91) | 0.30* | 196 (21.60) | 4238 (11.58) | 0.27* |
| >80; n (%) | 304 (22.12) | 5838 (10.81) | 0.31* | 70 (7.71) | 1150 (3.14) | 0.20* |
| ***Prior comorbidities*** |  |  |  |  |  |  |
| Charlson’s standard comorbidity index; mean (SD) | 2.9 (1.6) | 2.1 (1.5) | 0.54* | 2.2 (1.3) | 1.6 (1.2) | 0.48* |
| Duration from earliest diabetes mellitus diagnosis date to baseline date (day); mean (SD) | 7.2 (4.9) | 6.4 (4.9) | 0.16 | 6.8 (4.8) | 6.3 (4.7) | 0.10 |
| Number of hospitalizations; mean (SD) | 1.4 (1.2) | 1.2 (0.7) | 0.27* | 1.7 (1.5) | 1.2 (0.8) | 0.37* |
| Diabetic retinopathy; n (%) | 138 (10.04) | 3777 (6.99) | 0.11 | 87 (12.13) | 2660 (7.23) | 0.17 |
| Diabetic nephropathy; n (%) | 36 (2.62) | 668 (1.23) | 0.10 | 8 (1.11) | 224 (0.60) | 0.05 |
| Diabetic neuropathy; n (%) | 51 (3.71) | 1263 (2.33) | 0.08 | 68 (9.48) | 866 (2.35) | 0.31* |
| Gout; n (%) | 62 (4.51) | 1380 (2.55) | 0.11 | 30 (4.18) | 681 (1.85) | 0.14 |
| Heart failure; n (%) | 117 (8.51) | 2923 (5.41) | 0.12 | 50 (6.97) | 1714 (4.65) | 0.10 |
| Hyperlipidaemia; n (%) | 563 (40.97) | 21398 (39.62) | 0.03 | 376 (52.44) | 17380 (47.24) | 0.10 |
| Hypertension; n (%) | 591 (43.01) | 16568 (30.68) | 0.26* | 401 (55.92) | 12890 (35.04) | 0.43* |
| Hypoglycemia; n (%) | 33 (2.40) | 403 (0.74) | 0.13 | 8 (1.11) | 82 (0.22) | 0.11 |
| IHD with AMI; n (%) | 54 (3.93) | 1258 (2.32) | 0.09 | 24 (3.34) | 1048 (2.84) | 0.03 |
| IHD without AMI; n (%) | 163 (11.86) | 3460 (6.40) | 0.19 | 135 (18.82) | 2967 (8.06) | 0.32* |
| Liver diseases; n (%) | 45 (3.27) | 2005 (3.71) | 0.02 | 21 (2.92) | 1515 (4.11) | 0.06 |
| COPD; n (%) | 14 (1.01) | 832 (1.54) | 0.05 | 4 (0.55) | 188 (0.51) | 0.01 |
| Peripheral vascular disease; n (%) | 17 (1.23) | 397 (0.73) | 0.05 | 7 (0.97) | 187 (0.50) | 0.05 |
| Renal diseases; n (%) | 63 (4.58) | 922 (1.70) | 0.17 | 8 (1.11) | 180 (0.48) | 0.07 |
| Stroke/TIA; n (%) | 73 (5.31) | 1685 (3.12) | 0.11 | 33 (4.60) | 907 (2.46) | 0.12 |
| Atrial fibrillation; n (%) | 64 (4.65) | 1265 (2.34) | 0.13 | 29 (4.04) | 735 (1.99) | 0.12 |
| Anemia; n (%) | 89 (6.47) | 2223 (4.11) | 0.11 | 15 (2.09) | 899 (2.44) | 0.02 |
| Cancer; n (%) | 53 (3.85) | 1441 (2.66) | 0.07 | 19 (2.64) | 751 (2.04) | 0.04 |
| ***Drug exposure*** |  |  |  |  |  |  |
| SGLT2i vs. DPP4i; n (%) | 277 (20.16) | 18474 (34.21) | 0.32* | 277 (30.54) | 18474 (50.48) | 0.41* |
| SGLT2i frequency; mean (SD) | 9.2 (11.4)  n=277 | 7.1 (9.8)  n=18474 | 0.20 | 9.2 (11.4)  n=277 | 7.1 (9.8)  n=18474 | 0.20* |
| DPP4i frequency; mean (SD) | 5.6 (9.9)  n=1097 | 5.2 (7.2)  n=35522 | 0.04 | 5.6 (8.6)  n=630 | 4.5 (6.6)  n=18121 | 0.15 |
| SGLT2i duration (days); mean (SD) | 512.5 (654.3)  n=277 | 527.3 (671.1)  n=18474 | 0.02 | 512.5 (654.3)  n=277 | 527.3 (671.1)  n=18474 | 0.02 |
| DPP4i duration (days); mean (SD) | 483.3 (286.5)  n=1097 | 507.7 (286.0)  n=35522 | 0.09 | 491.3 (275.8)  n=630 | 481.5 (284.1)  n=18121 | 0.03 |
| Number of anti-diabetic drugs; mean (SD) | 483.3 (286.5)  n=1097 | 507.7 (286.0)  n=35522 | 0.09 | 497.4 (278.3)  n=440 | 498.4 (287.7)  n=18311 | <0.01 |
| Metformin; n (%) | 8.1 (18.1) | 8.0 (20.0) | 0.01 | 12.2 (25.6)  n=717 | 10.7 (26.2)  n=36785 | 0.06 |
| Sulphonylurea; n (%) | 1107 (80.56) | 48021 (88.93) | 0.23* | 650 (90.65) | 34186 (92.93) | 0.08 |
| Insulin; n (%) | 1095 (79.69) | 41439 (76.74) | 0.07 | 543 (75.73) | 26423 (71.83) | 0.09 |
| Acarbose; n (%) | 913 (66.44) | 26860 (49.74) | 0.34* | 446 (62.20) | 19174 (52.12) | 0.20* |
| Thiozolidinedone; n (%) | 57 (4.14) | 1978 (3.66) | 0.03 | 28 (3.90) | 1418 (3.85) | <0.01 |
| Glucagon-like peptide-1 receptor agonists; n (%) | 180 (13.10) | 10176 (18.84) | 0.16 | 107 (14.92) | 9186 (24.97) | 0.25* |
| ACEI/ARBs; n (%) | 109 (7.93) | 2815 (5.21) | 0.11 | 22 (3.06) | 2309 (6.27) | 0.15 |
| Antihepatitis; n (%) | 726 (52.83) | 23589 (43.68) | 0.18 | 434 (60.52) | 17017 (46.26) | 0.29* |
| Anticoagulants; n (%) | 31 (2.25) | 881 (1.63) | 0.05 | 10 (1.39) | 694 (1.88) | 0.04 |
| Antiplatelets; n (%) | 564 (41.04) | 15321 (28.37) | 0.27* | 303 (42.25) | 12207 (33.18) | 0.19 |
| Lipid-lowering drugs; n (%) | 565 (41.12) | 15489 (28.68) | 0.26* | 392 (54.67) | 10817 (29.40) | 0.53* |
| Statins and fibrates; n (%) | 637 (46.36) | 24604 (45.56) | 0.02 | 477 (66.52) | 21088 (57.32) | 0.19 |
| Nitrates; n (%) | 798 (58.07) | 30450 (56.39) | 0.03 | 589 (82.14) | 26254 (71.37) | 0.26* |
| Non-steroidal anti-inflammatory drugs; n (%) | 256 (18.63) | 6264 (11.60) | 0.20 | 216 (30.12) | 4569 (12.42) | 0.44* |
| Diuretics; n (%) | 450 (32.75) | 13222 (24.48) | 0.18 | 389 (54.25) | 10522 (28.60) | 0.54* |
| Beta-blockers; n (%) | 445 (32.38) | 13228 (24.49) | 0.18 | 349 (48.67) | 10221 (27.78) | 0.44* |
| Calcium channel blockers; n (%) | 462 (33.62) | 10811 (20.02) | 0.31* | 336 (46.86) | 8527 (23.18) | 0.51* |
| Anti-cancer drugs; n (%) | 36 (2.62) | 1482 (2.74) | 0.01 | 10 (1.39) | 694 (1.88) | 0.04 |
| Steroids/Corticosteroids; n (%) | 124 (9.02) | 4586 (8.49) | 0.02 | 31 (4.32) | 1749 (4.75) | 0.02 |
| ***Abbreviated MDRD*** ***(ml/min/1.73m^2^)*** |  |  |  |  |  |  |
| Abbreviated MDRD | 64.8 (27.4)  n=1220 | 79.7 (28.7)  n=44505 | 0.53* | 4.0 (0.44) | 34.0 (0.09) | 0.07 |
| Most severe renal damage (<15); n (%) | 46.0 (3.34) | 499.0 (0.92) | 0.17 | 4.0 (0.44) | 184.0 (0.50) | 0.01 |
| Severe renal damage [15, 30); n (%) | 62.0 (4.51) | 1344.0 (2.48) | 0.11 | 51.0 (5.62) | 851.0 (2.32) | 0.17 |
| Moderate to severe renal damage [30, 45); n (%) | 204.0 (14.84) | 3639.0 (6.73) | 0.26* | 93.0 (10.25) | 2504.0 (6.84) | 0.12 |
| Mild to moderate renal damage [45, 60); n (%) | 244.0 (17.75) | 5398.0 (9.99) | 0.23* | 423.0 (46.63) | 13594.0 (37.14) | 0.19 |
| Mild renal damage [60, 90]; n (%) | 432.0 (31.44) | 17484.0 (32.38) | 0.02 | 230.0 (25.35) | 13629.0 (37.24) | 0.26* |
| Chronic kidney disease (>90) | 232.0 (16.88) | 16141.0 (29.89) | 0.31* | 87.9 (6.8)  n=568 | 86.6 (7.7)  n=19924 | 0.18 |
| ***Complete blood counts*** |  |  |  |  |  |  |
| Mean corpuscular volume (fL) | 87.5 (7.6)  n=889 | 87.1 (7.6)  n=26918 | 0.05 | 0.17 (0.18)  n=526 | 0.21 (0.2)  n=16105 | 0.21* |
| Eosinophil (x10^9^/L) | 0.23 (0.27)  n=771 | 0.21 (0.24)  n=21528 | 0.05 | 1.8 (0.9)  n=526 | 2.1 (0.9)  n=16111 | 0.31* |
| Lymphocyte (x10^9^/L) | 1.8 (0.8)  n=771 | 2.0 (0.9)  n=21552 | 0.29* | 5.7 (2.7)  n=526 | 5.3 (2.7)  n=16111 | 0.15 |
| Neutrophil (x10^9^/L) | 5.5 (2.9)  n=771 | 5.4 (2.8)  n=21552 | 0.06 | 8.1 (2.8)  n=568 | 8.0 (2.7)  n=19949 | 0.01 |
| White cell count (x10^9^/L) | 7.9 (2.9)  n=889 | 8.0 (3.0)  n=26928 | 0.03 | 29.9 (2.8)  n=568 | 29.2 (3.1)  n=19924 | 0.26* |
| Mean cell haemoglobin (pg) | 29.7 (3.2)  n=889 | 29.4 (3.0)  n=26918 | 0.10 | 227.3 (67.6)  n=568 | 247.7 (71.2)  n=19947 | 0.29* |
| Platelet (x10^9^/L) | 229.1 (73.2)  n=889 | 242.7 (72.7)  n=26925 | 0.19 | 4.5 (0.6)  n=568 | 4.7 (0.6)  n=19924 | 0.26* |
| Red cell count (x10^12^/L) | 4.3 (0.7)  n=889 | 4.5 (0.7)  n=26918 | 0.29* | 4.26 (0.44)  n=804 | 4.29 (0.44)  n=30733 | 0.07 |
| ***Liver and renal functions; mean (SD)*** |  |  |  |  |  |  |
| Potassium (mmol/L) | 4.4 (0.52)  n=1219 | 4.35 (0.48)  n=44359 | 0.11 | 41.0 (4.2)  n=653 | 42.3 (3.6)  n=25245 | 0.33* |
| Albumin (g/L) | 40.1 (4.4)  n=1003 | 41.6 (4.0)  n=33810 | 0.36* | 138.9 (3.5)  n=805 | 139.3 (2.8)  n=30742 | 0.16 |
| Sodium (mmol/L) | 139.1 (3.3)  n=1220 | 139.3 (3.0)  n=44384 | 0.06 | 6.3 (2.7)  n=804 | 5.8 (2.2)  n=30752 | 0.21* |
| Urea (mmol/L) | 8.0 (4.9)  n=1217 | 6.6 (3.6)  n=44380 | 0.33* | 72.9 (6.1)  n=635 | 74.5 (5.1)  n=24145 | 0.28* |
| Protein (g/L) | 72.7 (6.3)  n=940 | 73.9 (5.5)  n=31781 | 0.20 | 92.1 (62.9)  n=805 | 79.4 (30.1)  n=30796 | 0.26* |
| Creatinine (umol/L) | 127.2 (122.8)  n=1220 | 94.2 (76.4)  n=44505 | 0.32* | 82.3 (43.1)  n=653 | 76.4 (33.3)  n=25266 | 0.15 |
| Alkaline phosphatase (U/L) | 80.9 (38.4)  n=1004 | 77.4 (33.3)  n=33921 | 0.10 | 26.0 (23.2)  n=394 | 28.7 (29.8)  n=10370 | 0.10 |
| Aspartate transaminase (U/L) | 26.6 (32.5)  n=461 | 27.9 (50.7)  n=13377 | 0.03 | 29.0 (31.5)  n=497 | 31.2 (30.3)  n=21268 | 0.07 |
| Alanine transaminase (U/L) | 24.6 (24.7)  n=856 | 28.8 (34.0)  n=28760 | 0.14 | 10.4 (8.0)  n=653 | 11.1 (6.7)  n=25200 | 0.09 |
| Bilirubin (umol/L) | 10.7 (6.6)  n=1003 | 11.1 (7.1)  n=33742 | 0.06 | 1.7 (1.6)  n=701 | 1.8 (1.5)  n=29290 | 0.01 |
| ***Lipid profile and variability measures; mean (SD)*** |  |  |  |  |  |  |
| Triglyceride (mmol/L) | 1.6 (1.3)  n=1104 | 1.7 (1.5)  n=41764 | 0.08 | 0.8 (1.5)  n=450 | 0.5 (1.0)  n=17127 | 0.24* |
| SD of triglyceride | 0.45 (0.78)  n=620 | 0.48 (0.97)  n=21312 | 0.02 | 2.39 (0.8)  n=694 | 2.41 (0.83)  n=28876 | 0.03 |
| Low-density lipoprotein (mmol/L) | 2.3 (0.8)  n=1088 | 2.4 (0.8)  n=41084 | 0.09 | 0.4 (0.32)  n=444 | 0.37 (0.34)  n=16717 | 0.09 |
| SD of low-density lipoprotein | 0.38 (0.35)  n=609 | 0.37 (0.35)  n=20724 | 0.04 | 1.18 (0.31)  n=701 | 1.2 (0.33)  n=29249 | 0.08 |
| High-density lipoprotein (mmol/L) | 1.21 (0.36)  n=1102 | 1.21 (0.33)  n=41707 | 0.01 | 0.1 (0.08)  n=445 | 0.1 (0.09)  n=16397 | 0.03 |
| SD of high-density lipoprotein | 0.11 (0.09)  n=609 | 0.1 (0.09)  n=20499 | 0.11 | 4.3 (1.1)  n=701 | 4.4 (1.0)  n=29311 | 0.07 |
| Total cholesterol (mmol/L) | 4.2 (1.0)  n=1104 | 4.4 (1.0)  n=41805 | 0.11 | 0.5 (0.5)  n=452 | 0.4 (0.4)  n=17148 | 0.18 |
| SD of total cholesterol | 0.5 (0.4)  n=622 | 0.4 (0.4)  n=21339 | 0.05 | 8.1 (1.8)  n=746 | 8.2 (1.6)  n=30189 | 0.07 |
| ***Glucose tests and variability measures*** |  |  |  |  |  |  |
| Hemoglobin A1C (%) | 7.9 (1.6)  n=1167 | 8.0 (1.5  )n=43651 | 0.06 | 8.0 (1.5)  n=731 | 8.1 (1.5)  n=29490 | 0.08 |
| Mean hemoglobin A1C (%) | 7.9 (1.4)  n=1121 | 8.0 (1.4)  n=42585 | 0.07 | 1.2 (4.2)  n=560 | 0.9 (9.8)  n=23054 | 0.04 |
| Variance of hemoglobin A1C | 0.9 (3.1)  n=815 | 0.8 (8.3)  n=30658 | 0.02 | 0.7 (0.9)  n=560 | 0.6 (0.8)  n=23054 | 0.13 |
| SD of hemoglobin A1C | 0.63 (0.73)  n=815 | 0.56 (0.7)  n=30658 | 0.10 | 6.5 (6.7)  n=516 | 5.3 (5.0)  n=22522 | 0.20* |
| Fasting glucose (mmol/L) | 8.7 (4.0)  n=1089 | 8.9 (3.8)  n=39462 | 0.04 | 9.0 (3.3)  n=709 | 8.9 (3.0)  n=28043 | 0.03 |
| Mean fasting glucose (mmol/L) | 8.74 (3.18)  n=1117 | 8.74 (2.92)  n=40294 | <0.01 | 8.8 (15.5)  n=517 | 8.0 (26.5)  n=18865 | 0.04 |
| Variance of fasting glucose | 12.2 (30.0)  n=746 | 8.7 (30.4)  n=24388 | 0.12 | 2.2 (1.9)  n=518 | 1.9 (2.1)  n=18890 | 0.15 |
| SD of fasting glucose | 2.5 (2.4)  n=747 | 2.0 (2.2)  n=24413 | 0.24* | 17.7 (12.6)  n=475 | 16.1 (13.1)  n=18416 | 0.12 |

* for SMD$\geq$0.2; ACEI: angiotensin-converting enzyme inhibitor; AMI: acute myocardial infarction; ARB: angiotensin receptor blocker; COPD: chronic obstructive pulmonary disease; DPP4i: dipeptidyl peptidase-4 inhibitor; IHD: ischemic heart disease; MDRD: Modification of Diet in Renal Disease; SD: standard deviation; SGLT2i: sodium glucose cotransporter-2 inhibitor; SMD: standardized mean difference; TIA: transient ischemic attack.

Table S3. Overall annual incidence ratios of primary and secondary outcomes in the matched cohort.

| **Cohort** | **Duration** | **Person-years** | **Failures** | **Rate** | **95% CI** | |
| --- | --- | --- | --- | --- | --- | --- |
|  |  |  |  |  | **Lower** | **Upper** |
| **New-onset syncope** | (0 - 1] | 37359.0 | 225 | 6 | 5.3 | 6.9 |
|  | (1 - 2] | 37066.1 | 296 | 8 | 7.1 | 8.9 |
|  | (2 - 3] | 36509.9 | 171 | 4.7 | 4 | 5.4 |
|  | (3 - 4] | 35711.0 | 123 | 3.4 | 2.9 | 4.1 |
|  | (4 - 5] | 34927.3 | 91 | 2.6 | 2.1 | 3.2 |
|  | (5 - 6] | 20410.6 | 1 | 0 | 0 | 0.3 |
|  | Total | - | 907 | 4.5 | 4.2 | 4.8 |
| **Cardiovascular mortality** | (0 - 1] | 37497.1 | 5 | 0.1 | 0.1 | 0.3 |
|  | (1 - 2] | 37420.6 | 22 | 0.6 | 0.4 | 0.9 |
|  | (2 - 3] | 37087.1 | 124 | 3.3 | 2.8 | 4.0 |
|  | (3 - 4] | 36393.5 | 130 | 3.6 | 3.0 | 4.2 |
|  | (4 - 5] | 35693.0 | 146 | 4.1 | 3.5 | 4.8 |
|  | (5 - 6] | 20884.7 | 44 | 2.1 | 1.6 | 2.8 |
|  | Total | - | 471 | 2.3 | 2.1 | 2.5 |
| **All-cause mortality** | (0 - 1] | 37497.1 | 12 | 0.3 | 0.2 | 0.6 |
|  | (1 - 2] | 37420.6 | 198 | 5.3 | 4.6 | 6.1 |
|  | (2 - 3] | 37087.1 | 463 | 12.5 | 11.4 | 13.7 |
|  | (3 - 4] | 36393.5 | 805 | 22.1 | 20.6 | 23.7 |
|  | (4 - 5] | 35693.0 | 636 | 17.8 | 16.5 | 19.3 |
|  | (5 - 6] | 20884.7 | 232 | 11.1 | 9.8 | 12.6 |
|  | Total | - | 2346 | 11.4 | 11.0 | 11.9 |

CI: confidence interval.

Table S4. Univariate Cox regression models with adjustments to predict new-onset syncope before and after propensity score matching.

| **Characteristics** | **Before matching** | | **After matching** | |
| --- | --- | --- | --- | --- |
|  | **HR [95% CI]** | **P value** | **HR [95% CI]** | **P value** |
| ***Demographics*** |  |  |  |  |
| Male | 1.62 [1.45-1.80] | <0.001 | 1.60 [1.37-1.86] | <0.001 |
| Female | 0.62 [0.55-0.69] | <0.001 | 0.63 [0.54-0.73] | <0.001 |
| Baseline age | 1.05 [1.04-1.05] | <0.001 | 1.05 [1.05-1.06] | <0.001 |
| 18-50 | 0.27 [0.21-0.35] | <0.001 | 0.26 [0.19-0.35] | <0.001 |
| 50-60 | 0.53 [0.46-0.61] | <0.001 | 0.73 [0.62-0.86] | <0.001 |
| 60-70 | 0.83 [0.74-0.94] | 0.003 | 1.12 [0.96-1.31] | 0.150 |
| 70-80 | 2.02 [1.80-2.26] | <0.001 | 2.78 [2.35-3.29] | <0.001 |
| >80 | 2.55 [2.24-2.90] | <0.001 | 2.20 [1.62-2.99] | <0.001 |
| ***Past comorbidities*** |  |  |  |  |
| Charlson’s standard comorbidity index | 1.34 [1.30-1.37] | <0.001 | 1.36 [1.30-1.42] | <0.001 |
| Duration from earliest diabetes mellitus diagnosis date to baseline date (day) | 1.03 [1.02-1.04] | <0.001 | 1.02 [1.01-1.04] | 0.005 |
| Number of hospitalizations | 1.24 [1.21-1.28] | <0.001 | 1.26 [1.23-1.30] | <0.001 |
| Diabetic retinopathy | 1.50 [1.26-1.79] | <0.001 | 1.76 [1.41-2.21] | <0.001 |
| Diabetic nephropathy | 2.24 [1.61-3.12] | <0.001 | 1.85 [0.92-3.71] | 0.084 |
| Diabetic neuropathy | 1.62 [1.23-2.15] | <0.001 | 4.32 [3.37-5.55] | <0.001 |
| Gout | 1.85 [1.43-2.39] | <0.001 | 2.30 [1.59-3.31] | <0.001 |
| Heart failure | 1.68 [1.39-2.03] | <0.001 | 1.55 [1.16-2.07] | 0.003 |
| Hyperlipidaemia | 1.06 [0.95-1.18] | 0.306 | 1.23 [1.06-1.42] | 0.006 |
| Hypertension | 1.73 [1.55-1.92] | <0.001 | 2.36 [2.04-2.73] | <0.001 |
| Hypoglycemia | 3.37 [2.38-4.76] | <0.001 | 4.85 [2.42-9.74] | <0.001 |
| IHD with AMI | 1.76 [1.34-2.31] | <0.001 | 1.21 [0.81-1.82] | 0.359 |
| IHD without AMI | 1.97 [1.67-2.32] | <0.001 | 2.64 [2.19-3.18] | <0.001 |
| Liver diseases | 0.88 [0.66-1.19] | 0.410 | 0.70 [0.45-1.08] | 0.110 |
| COPD | 0.64 [0.38-1.09] | 0.101 | 1.08 [0.40-2.88] | 0.880 |
| Peripheral vascular disease | 1.80 [1.11-2.90] | 0.016 | 1.99 [0.94-4.18] | 0.071 |
| Renal diseases | 2.92 [2.26-3.75] | <0.001 | 2.38 [1.19-4.78] | 0.015 |
| Stroke/TIA | 1.78 [1.41-2.25] | <0.001 | 1.91 [1.35-2.71] | <0.001 |
| Atrial fibrillation | 2.13 [1.66-2.74] | <0.001 | 2.11 [1.46-3.06] | <0.001 |
| Anemia | 1.68 [1.36-2.08] | <0.001 | 0.86 [0.52-1.43] | 0.563 |
| Cancer | 1.51 [1.15-1.99] | 0.003 | 1.34 [0.85-2.11] | 0.211 |
| ***Medications*** |  |  |  |  |
| SGLT2i vs. DPP4i | 0.47 [0.41-0.54] | <0.001 | 0.62 [0.53-0.72] | <0.001 |
| Dapagliflozin vs. DPP4i | 0.56 [0.47-0.66] | <0.001 | 0.75 [0.63-0.89] | 0.001 |
| Empagliflozin vs. DPP4i | 0.58 [0.45-0.76] | <0.001 | 0.77 [0.59-1.01] | 0.058 |
| Canagliflozin vs. DPP4i | 0.48 [0.38-0.62] | <0.001 | 0.64 [0.49-0.82] | <0.001 |
| Ertugliflozin vs. DPP4i | 0.43 [0.29-0.62] | <0.001 | 0.56 [0.38-0.82] | 0.003 |
| DPP4i frequency | 1.00 [1.00-1.01] | 0.106 | 1.01 [1.00-1.02] | 0.046 |
| SGLT2i duration (day) | 1.000 [1.000-1.000] | 0.700 | 1.000 [1.000-1.000] | 0.700 |
| DPP4i duration (day) | 1.000 [0.999-1.000] | 0.001 | 1.000 [1.000-1.000] | 0.796 |
| Number of anti-diabetic drugs | 1.000 [0.998-1.003] | 0.887 | 1.002 [0.999-1.004] | 0.123 |
| Metformin | 0.49 [0.43-0.56] | <0.001 | 0.73 [0.57-0.94] | 0.014 |
| Sulphonylurea | 1.19 [1.04-1.35] | 0.011 | 1.22 [1.03-1.45] | 0.021 |
| Insulin | 2.05 [1.84-2.30] | <0.001 | 1.53 [1.32-1.78] | <0.001 |
| Acarbose | 1.14 [0.88-1.49] | 0.329 | 1.02 [0.70-1.48] | 0.931 |
| Thiozolidinedone | 0.64 [0.55-0.75] | <0.001 | 0.52 [0.43-0.64] | <0.001 |
| Glucagon-like peptide-1 receptor agonists | 1.56 [1.28-1.90] | <0.001 | 0.47 [0.31-0.72] | <0.001 |
| ACEI/ARBs | 1.45 [1.30-1.61] | <0.001 | 1.78 [1.53-2.07] | <0.001 |
| Antihepatitis | 1.39 [0.97-1.98] | 0.072 | 0.73 [0.39-1.37] | 0.332 |
| Anticoagulants | 1.77 [1.59-1.97] | <0.001 | 1.48 [1.28-1.72] | <0.001 |
| Antiplatelets | 1.75 [1.57-1.95] | <0.001 | 2.90 [2.50-3.36] | <0.001 |
| Lipid-lowering drugs | 1.03 [0.93-1.15] | 0.532 | 1.48 [1.27-1.73] | <0.001 |
| Statins and fibrates | 1.06 [0.95-1.18] | 0.306 | 1.84 [1.52-2.23] | <0.001 |
| Nitrates | 1.76 [1.53-2.01] | <0.001 | 3.05 [2.60-3.58] | <0.001 |
| Non-steroidal anti-inflammatory drugs | 1.51 [1.35-1.69] | <0.001 | 2.96 [2.56-3.43] | <0.001 |
| Diuretics | 1.50 [1.34-1.68] | <0.001 | 2.48 [2.14-2.87] | <0.001 |
| Beta-blockers | 2.03 [1.82-2.27] | <0.001 | 2.93 [2.53-3.39] | <0.001 |
| Calcium channel blockers | 1.31 [1.18-1.46] | <0.001 | 2.04 [1.76-2.37] | <0.001 |
| Anti-cancer drugs | 1.08 [0.90-1.30] | 0.404 | 0.91 [0.63-1.30] | 0.607 |
| Steroids/Corticosteroids | 2.28 [1.83-2.84] | <0.001 | 2.01 [1.26-3.21] | 0.004 |
| ***Subclinical biomarker*** |  |  |  |  |
| Abbreviated MDRD (ml/min/1.73m^2^) | 0.981 [0.979-0.983] | <0.001 | 0.98 [0.98-0.99] | <0.001 |
| Most severe renal damage (<15 ml/min/1.73m^2^) | 3.65 [2.72-4.91] | <0.001 | 4.74 [2.46-9.16] | <0.001 |
| Severe renal damage ([15, 30) ml/min/1.73m^2^) | 1.88 [1.46-2.43] | <0.001 | 2.78 [1.76-4.39] | <0.001 |
| Moderate to severe renal damage ([30, 45) ml/min/1.73m^2^) | 2.32 [2.00-2.70] | <0.001 | 1.26 [0.89-1.77] | 0.190 |
| Mild to moderate renal damage ([45, 60) ml/min/1.73m^2^) | 1.82 [1.58-2.09] | <0.001 | 2.42 [2.00-2.94] | <0.001 |
| Mild renal damage ([60, 90] ml/min/1.73m^2^) | 0.84 [0.75-0.94] | 0.004 | 0.97 [0.83-1.14] | 0.734 |
| Chronic kidney disease (>90 ml/min/1.73m^2^) | 0.41 [0.35-0.47] | <0.001 | 0.53 [0.44-0.63] | <0.001 |
| ***Complete blood counts*** |  |  |  |  |
| Mean corpuscular volume (fL) | 1.01 [1.00-1.02] | 0.116 | 1.01 [1.00-1.02] | 0.102 |
| Eosinophil (x10^9^/L) | 1.15 [0.97-1.37] | 0.116 | 0.43 [0.23-0.81] | 0.009 |
| Lymphocyte (x10^9^/L) | 0.65 [0.59-0.72] | <0.001 | 0.60 [0.52-0.69] | <0.001 |
| Neutrophil (x10^9^/L) | 1.02 [1.00-1.05] | 0.042 | 1.07 [1.04-1.09] | <0.001 |
| White cell count (x10^9^/L) | 0.99 [0.97-1.02] | 0.502 | 1.03 [0.99-1.06] | 0.103 |
| Mean cell haemoglobin (pg) | 1.04 [1.01-1.06] | 0.002 | 1.07 [1.04-1.11]0.0001 | <0.001 |
| Platelet (x10^9^/L) | 0.997 [0.996-0.998] | <0.001 | 0.997 [0.995-0.998] | <0.001 |
| Red cell count (x10^12^/L) | 0.63 [0.57-0.69] | <0.001 | 0.68 [0.58-0.79] | <0.001 |
| ***Liver and renal functions*** |  |  |  |  |
| Potassium (mmol/L) | 1.26 [1.12-1.41] | <0.001 | 1.03 [0.87-1.23] | 0.700 |
| Albumin (g/L) | 0.92 [0.91-0.93] | <0.001 | 0.91 [0.89-0.93] | <0.001 |
| Sodium (mmol/L) | 0.98 [0.96-1.00] | 0.019 | 0.97 [0.94-1.00] | 0.022 |
| Urea (mmol/L) | 1.07 [1.06-1.07] | <0.001 | 1.08 [1.06-1.10] | <0.001 |
| Protein (g/L) | 0.96 [0.95-0.97] | <0.001 | 0.94 [0.93-0.96] | <0.001 |
| Creatinine (umol/L) | 1.002 [1.002-1.002] | <0.001 | 1.003 [1.002-1.003] | <0.001 |
| Alkaline phosphatase (U/L) | 1.002 [1.001-1.004] | <0.001 | 1.01 [1.00-1.01] | <0.001 |
| Aspartate transaminase (U/L) | 0.999 [0.996-1.002] | 0.593 | 0.999 [0.995-1.004] | 0.786 |
| Alanine transaminase (U/L) | 0.991 [0.987-0.995] | <0.001 | 1.003 [1.002-1.005] | <0.001 |
| Bilirubin (umol/L) | 0.99 [0.98-1.00] | 0.048 | 1.01 [1.00-1.01] | <0.001 |
| ***Lipid profile and variabilities*** |  |  |  |  |
| Triglyceride (mmol/L) | 0.93 [0.88-0.98] | 0.010 | 1.03 [0.99-1.06] | 0.102 |
| SD of triglyceride | 0.97 [0.89-1.07] | 0.560 | 1.06 [1.00-1.13] | 0.038 |
| Low-density lipoprotein (mmol/L) | 0.89 [0.82-0.96] | 0.003 | 0.94 [0.85-1.05] | 0.266 |
| SD of low-density lipoprotein | 1.13 [0.91-1.41] | 0.269 | 1.19 [0.88-1.62] | 0.258 |
| High-density lipoprotein (mmol/L) | 1.04 [0.87-1.24] | 0.639 | 1.08 [0.86-1.37] | 0.499 |
| SD of high-density lipoprotein | 3.58 [1.61-7.97] | 0.002 | 2.65 [0.85-8.31] | 0.095 |
| Total cholesterol (mmol/L) | 0.89 [0.83-0.94] | <0.001 | 0.98 [0.90-1.06] | 0.570 |
| SD of total cholesterol | 1.11 [0.93-1.32] | 0.251 | 1.10 [0.86-1.39] | 0.456 |
| ***Glucose tests and variabilities*** |  |  |  |  |
| Hemoglobin A1C (%) | 0.96 [0.92-1.00] | 0.032 | 1.00 [0.95-1.05] | 0.868 |
| Mean hemoglobin A1C (%) | 0.95 [0.90-0.99] | 0.016 | 0.95 [0.89-1.01] | 0.094 |
| Variance of hemoglobin A1C | 1.00 [1.00-1.01] | 0.633 | 1.00 [1.00-1.01] | 0.308 |
| SD of hemoglobin A1C | 1.10 [1.04-1.17] | 0.001 | 1.10 [1.03-1.18] | 0.003 |
| CV of hemoglobin A1C | 1.02 [1.01-1.04] | <0.001 | 1.03 [1.01-1.04] | <0.001 |
| Fasting glucose (mmol/L) | 0.99 [0.97-1.01] | 0.252 | 1.02 [1.00-1.03] | 0.115 |
| Mean fasting glucose (mmol/L) | 1.00 [0.98-1.02] | 0.851 | 1.02 [1.00-1.05] | 0.087 |
| Variance of fasting glucose | 1.002 [1.001-1.003] | <0.001 | 1.001 [0.999-1.004] | 0.285 |
| SD of fasting glucose | 1.09 [1.06-1.11] | <0.001 | 1.06 [1.02-1.09] | 0.003 |
| CV of fasting glucose | 1.02 [1.02-1.03] | <0.001 | 1.01 [1.00-1.01] | 0.025 |
| SGLT2i vs. DPP4i | 0.47 [0.41-0.54] | <0.001 | 0.62 [0.53-0.72] | <0.001 |

ACEI: angiotensin-converting enzyme inhibitor; AMI: acute myocardial infarction; ARB: angiotensin receptor blocker; CI: confidence interval; COPD: chronic obstructive pulmonary disease; CV: coefficient of variation; DPP4i: dipeptidyl peptidase-4 inhibitor; HR: hazard ratio; IHD: ischemic heart disease; MDRD: Modification of Diet in Renal Disease; SD: standard deviation; SGLT2i: sodium glucose cotransporter-2 inhibitor; TIA: transient ischemic attack.

Table S5. Subgroup analysis for prior comorbidity measured by Charlson’s index for the exposure effects of SGLT2i vs. DPP4i on new-onset syncope.

| **Charlson’s index** | **SGLT2i vs. DPP4i** | | **Dapagliflozin vs. DPP4i** | | **Empagliflozin vs. DPP4i** | | **Canagliflozin vs. DPP4i** | | **Ertugliflozin vs. DPP4i** | |
| --- | --- | --- | --- | --- | --- | --- | --- | --- | --- | --- |
|  | **HR [95% CI]** | **P value** | **HR [95% CI]** | **P value** | **HR [95% CI]** | **P value** | **HR [95% CI]** | **P value** | **HR [95% CI]** | **P value** |
| 0 | 0.19 [0.11-0.32] | <0.001 | 0.25 [0.13-0.48] | <0.001 | 0.18 [0.04-0.73] | 0.016 | 0.18 [0.04-0.73] | 0.058 | 0.14 [0.02-1.01] | 0.052 |
| 1 | 0.36 [0.28-0.47] | <0.001 | 0.49 [0.36-0.68] | <0.001 | 0.60 [0.37-0.95] | 0.031 | 0.47 [0.29-0.76] | 0.002 | 0.45 [0.22-0.91] | 0.027 |
| 2 | 0.68 [0.53-0.88] | 0.004 | 0.77 [0.57-1.04] | 0.094 | 0.91 [0.59-1.39] | 0.659 | 0.78 [0.52-1.18] | 0.242 | 0.52 [0.26-1.00] | 0.051 |
| 3 | 0.47 [0.34-0.64] | <0.001 | 0.67 [0.46-0.97] | 0.035 | 0.53 [0.27-1.03] | 0.063 | 0.33 [0.17-0.65] | 0.001 | 0.31 [0.12-0.84] | 0.022 |
| 4 | 0.54 [0.34-0.88] | 0.012 | 0.77 [0.44-1.33] | 0.346 | 0.55 [0.20-1.51] | 0.247 | 0.43 [0.16-1.17] | 0.098 | 0.58 [0.18-1.84] | 0.354 |
| 5+ | 0.41 [0.21-0.78] | 0.007 | 0.42 [0.18-0.98] | 0.046 | 0.79 [0.24-2.52] | 0.685 | 0.16 [0.02-1.14] | 0.067 | 0.99 [0.31-3.17] | 0.982 |

CI: confidence interval; DPP4i: dipeptidyl peptidase-4 inhibitor; HR: adjusted hazard ratio; SGLT2i: sodium-glucose cotransporter-2 inhibitor.

Table S6. Subgroup analysis for individual comorbidity at baseline for the exposure effects of SGLT2i vs. DPP4i on new-onset syncope.

| **Subgroup** | **SGLT2i vs. DPP4i** | | | **Dapagliflozin vs. DPP4i** | | | **Empagliflozin vs. DPP4i** | | | **Canagliflozin vs. DPP4i** | | | **Ertugliflozin vs. DPP4i** | | |
| --- | --- | --- | --- | --- | --- | --- | --- | --- | --- | --- | --- | --- | --- | --- | --- |
|  | **HR [95% CI]** | | **P value** | **HR [95% CI]** | | **P value** | **HR [95% CI]** | | **P value** | **HR [95% CI]** | | **P value** | **HR [95% CI]** | | **P value** |
| Anemia (No) | 0.43 [0.37-0.49] | <0.001 | | 0.55 [0.46-0.65] | <0.001 | | 0.60 [0.46-0.79]0.0002 | <0.001 | | 0.50 [0.39-0.65] | <0.001 | | 0.44 [0.30-0.65] | <0.001 | |
| Anemia (Yes) | 0.39 [0.15-1.00] | 0.051 | | 0.79 [0.29-2.14] | 0.638 | | 0.39 [0.05-2.90] | 0.357 | | - | - | | - | - | |
| Atrial fibrillation (No) | 0.41 [0.36-0.48] | <0.001 | | 0.54 [0.45-0.64] | <0.001 | | 0.59 [0.45-0.77]0.0001 | <0.001 | | 0.47 [0.36-0.61] | <0.001 | | 0.44 [0.30-0.65] | <0.001 | |
| Atrial fibrillation (Yes) | 0.86 [0.47-1.57] | 0.621 | | 1.04 [0.52-2.07] | 0.915 | | 0.81 [0.29-2.27] | 0.686 | | 0.99 [0.42-2.35] | 0.982 | | 0.31 [0.04-2.28] | 0.252 | |
| Cancer (No) | 0.44 [0.38-0.50] | <0.001 | | 0.56 [0.47-0.66] | <0.001 | | 0.61 [0.47-0.80]0.0003 | <0.001 | | 0.51 [0.39-0.65] | <0.001 | | 0.45 [0.31-0.65] | <0.001 | |
| Cancer (Yes) | 0.18 [0.06-0.53] | 0.002 | | 0.44 [0.15-1.28] | 0.131 | | - | - | | - | - | | - | - | |
| Heart failure (No) | 0.41 [0.35-0.47] | <0.001 | | 0.54 [0.46-0.65] | <0.001 | | 0.57 [0.43-0.75] | <0.001 | | 0.49 [0.38-0.63]< | <0.001 | | 0.41 [0.27-0.61] | <0.001 | |
| Heart failure (Yes) | 1.12 [0.57-2.17] | 0.746 | | 0.90 [0.42-1.93] | 0.790 | | 1.39 [0.54-3.59] | 0.493 | | 0.57 [0.18-1.88] | 0.359 | | 1.13 [0.35-3.69] | 0.841 | |
| Hyperlipidaemia (No) | 0.42 [0.37-0.49] | <0.001 | | 0.54 [0.46-0.64] | <0.001 | | 0.60 [0.46-0.78] | <0.001 | | 0.50 [0.39-0.64] | <0.001 | | 0.45 [0.31-0.66] | <0.001 | |
| Hyperlipidaemia (Yes) | 0.65 [0.28-1.52] | 0.318 | | 1.17 [0.48-2.86] | 0.738 | | 0.45 [0.06-3.31] | 0.429 | | 0.30 [0.04-2.24] | 0.242 | | - | - | |
| Hypertension (No) | 0.38 [0.32-0.45] | <0.001 | | 0.50 [0.40-0.61] | <0.001 | | 0.52 [0.37-0.72]0.0001 | <0.001 | | 0.50 [0.37-0.67] | <0.001 | | 0.37 [0.23-0.61]0.0001 | <0.001 | |
| Hypertension (Yes) | 0.54 [0.43-0.69] | <0.001 | | 0.69 [0.52-0.92] | 0.012 | | 0.82 [0.54-1.26] | 0.374 | | 0.49 [0.31-0.77] | 0.002 | | 0.61 [0.33-1.11] | 0.104 | |
| IHD with AMI (No) | 0.41 [0.36-0.48] | <0.001 | | 0.53 [0.45-0.63] | <0.001 | | 0.56 [0.43-0.74]<0.001 | <0.001 | | 0.50 [0.39-0.65] | <0.001 | | 0.46 [0.31-0.67] | <0.001 | |
| IHD with AMI (Yes) | 1.32 [0.59-2.98] | 0.500 | | 2.02 [0.88-4.61] | 0.097 | | 2.13 [0.79-5.70] | 0.133 | | 0.22 [0.03-1.66] | 0.143 | | - | - | |
| IHD without AMI (No) | 0.42 [0.36-0.49] | <0.001 | | 0.58 [0.48-0.69] | <0.001 | | 0.51 [0.38-0.70]<0.001 | <0.001 | | 0.48 [0.36-0.63] | <0.001 | | 0.36 [0.23-0.57] | <0.001 | |
| IHD without AMI (Yes) | 0.43 [0.30-0.60] | <0.001 | | 0.46 [0.29-0.72] | <0.001 | | 1.02 [0.61-1.69] | 0.944 | | 0.50 [0.29-0.86] | 0.012 | | 0.73 [0.37-1.44] | 0.367 | |
| Liver diseases (No) | 0.43 [0.37-0.50] | <0.001 | | 0.56 [0.47-0.67] | <0.001 | | 0.60 [0.46-0.78] | <0.001 | | 0.49 [0.38-0.64] | <0.001 | | 0.42 [0.29-0.62] | <0.001 | |
| Liver diseases (Yes) | 0.41 [0.21-0.79] | 0.008 | | 0.40 [0.17-0.95] | 0.038 | | 0.66 [0.20-2.13] | 0.487 | | 0.47 [0.15-1.53] | 0.211 | | 0.85 [0.20-3.51] | 0.819 | |
| Renal diseases (No) | 0.43 [0.37-0.50] | <0.001 | | 0.56 [0.47-0.66] | <0.001 | | 0.60 [0.47-0.79] | <0.001 | | 0.50 [0.39-0.64] | <0.001 | | 0.43 [0.29-0.62] | <0.001 | |
| Renal diseases (Yes) | 0.12 [0.02-0.96] | 0.046 | | - | - | | - | - | | - | - | | 1.84 [0.23-14.73] | 0.565 | |
| Stroke/TIA (No) | 0.42 [0.36-0.49] | <0.001 | | 0.54 [0.45-0.65] | <0.001 | | 0.59 [0.45-0.78] | <0.001 | | 0.51 [0.39-0.66] | <0.001 | | 0.43 [0.29-0.64] | <0.001 | |
| Stroke/TIA (Yes) | 0.55 [0.32-0.96] | 0.036 | | 0.73 [0.39-1.36] | 0.321 | | 0.73 [0.26-2.02] | 0.544 | | 0.27 [0.07-1.11] | 0.070 | | 0.55 [0.13-2.24] | 0.401 | |

AMI: acute myocardial infarction; CI: confidence interval; DPP4i: dipeptidyl peptidase-4 inhibitor; HR: hazard ratio; IHD: ischemic heart disease; SGLT2i: sodium-glucose cotransporter-2 inhibitor; TIA: transient ischemic attack.

Table S7. Subgroup analysis for baseline medication prescriptions for the exposure effects of SGLT2i vs. DPP4i on new-onset syncope.

| **Subgroup** | **SGLT2i vs. DPP4i** | | **Dapagliflozin vs. DPP4i** | | **Empagliflozin vs. DPP4i** | | **Canagliflozin vs. DPP4i** | | **Ertugliflozin vs. DPP4i** | |
| --- | --- | --- | --- | --- | --- | --- | --- | --- | --- | --- |
|  | **HR [95% CI]** | **P value** | **HR [95% CI]** | **P value** | **HR [95% CI]** | **P value** | **HR [95% CI]** | **P value** | **HR [95% CI]** | **P value** |
| ACEI/ARBs (No) | 0.52 [0.39-0.68] | <0.001 | 0.66 [0.48-0.91] | 0.011 | 0.56 [0.33-0.94] | 0.029 | 0.59 [0.37-0.96] | 0.032 | 0.39 [0.17-0.87] | 0.022 |
| ACEI/ARBs (Yes) | 0.37 [0.31-0.44] | <0.001 | 0.50 [0.41-0.61] | <0.001 | 0.62 [0.46-0.83] | 0.002 | 0.44 [0.33-0.59] | <0.001 | 0.44 [0.29-0.67] | <0.001 |
| Anticoagulants (No) | 0.42 [0.36-0.49] | <0.001 | 0.61 [0.51-0.73] | <0.001 | 0.65 [0.48-0.87] | 0.004 | 0.44 [0.33-0.58] | <0.001 | 0.37 [0.25-0.55] | <0.001 |
| Anticoagulants (Yes) | 0.43 [0.30-0.63] | <0.001 | 0.46 [0.30-0.70] | <0.001 | 0.60 [0.34-1.07] | 0.082 | 0.76 [0.40-1.45] | 0.411 | 0.98 [0.24-3.94] | 0.973 |
| Antiplatelets (No) | 0.39 [0.32-0.48] | <0.001 | 0.54 [0.43-0.69] | <0.001 | 0.36 [0.22-0.57] | <0.001 | 0.47 [0.32-0.69] | <0.001 | 0.44 [0.25-0.77] | 0.004 |
| Antiplatelets (Yes) | 0.43 [0.35-0.52] | <0.001 | 0.59 [0.46-0.74] | <0.001 | 0.82 [0.60-1.12] | 0.217 | 0.45 [0.32-0.63] | <0.001 | 0.39 [0.23-0.65] | <0.001 |
| Beta-blockers (No) | 0.44 [0.37-0.52] | <0.001 | 0.55 [0.44-0.67] | <0.001 | 0.54 [0.38-0.76] | <0.001 | 0.56 [0.41-0.76] | <0.001 | 0.43 [0.26-0.69] | <0.001 |
| Beta-blockers (Yes) | 0.39 [0.31-0.50] | <0.001 | 0.58 [0.43-0.77] | <0.001 | 0.69 [0.46-1.04] | 0.077 | 0.36 [0.23-0.56] | <0.001 | 0.42 [0.23-0.77] | 0.005 |
| Calcium channel blockers (No) | 0.39 [0.32-0.49] | <0.001 | 0.54 [0.42-0.70] | <0.001 | 0.52 [0.35-0.78] | 0.001 | 0.46 [0.31-0.68] | <0.001 | 0.27 [0.13-0.54] | <0.001 |
| Calcium channel blockers (Yes) | 0.44 [0.37-0.54] | <0.001 | 0.55 [0.44-0.70] | <0.001 | 0.68 [0.48-0.96] | 0.029 | 0.51 [0.36-0.71] | <0.001 | 0.58 [0.37-0.91] | 0.017 |
| Diuretics (No) | 0.39 [0.32-0.47] | <0.001 | 0.50 [0.40-0.63] | <0.001 | 0.45 [0.31-0.66] | <0.001 | 0.57 [0.42-0.78] | <0.001 | 0.40 [0.24-0.67] | <0.001 |
| Diuretics (Yes) | 0.47 [0.37-0.58] | <0.001 | 0.64 [0.50-0.83] | <0.001 | 0.84 [0.58-1.21] | 0.341 | 0.37 [0.24-0.57] | <0.001 | 0.47 [0.27-0.81] | 0.007 |
| GLP-1RA (No) | 0.41 [0.35-0.47] | <0.001 | 0.53 [0.44-0.63] | <0.001 | 0.60 [0.46-0.78] | <0.001 | 0.48 [0.37-0.62] | <0.001 | 0.41 [0.28-0.61] | <0.001 |
| GLP-1RA (Yes) | - | - | 4.38 [1.54-12.42] | 0.006 | 0.59 [0.08-4.43] | 0.607 | 1.41 [0.40-4.89] | 0.592 | 1.85 [0.42-8.07] | 0.416 |
| Insulin (No) | 0.35 [0.28-0.44] | <0.001 | 0.49 [0.37-0.65] | <0.001 | 0.44 [0.28-0.70] | <0.001 | 0.48 [0.32-0.71] | <0.001 | 0.32 [0.17-0.63] | <0.001 |
| Insulin (Yes) | 0.50 [0.42-0.59] | <0.001 | 0.60 [0.49-0.75] | <0.001 | 0.73 [0.53-1.01] | 0.056 | 0.51 [0.37-0.71] | <0.001 | 0.54 [0.34-0.86] | 0.009 |
| Lipid-lowering drugs (No) | 0.36 [0.27-0.47] | <0.001 | 0.46 [0.32-0.65] | <0.001 | 0.36 [0.20-0.66] | 0.001 | 0.51 [0.31-0.83] | 0.007 | 0.35 [0.16-0.78] | 0.011 |
| Lipid-lowering drugs (Yes) | 0.44 [0.37-0.52] | <0.001 | 0.58 [0.48-0.70] | <0.001 | 0.70 [0.53-0.94] | 0.019 | 0.47 [0.35-0.64] | <0.001 | 0.46 [0.30-0.71] | <0.001 |
| Metformin (No) | 0.34 [0.19-0.61] | <0.001 | 0.39 [0.19-0.80] | 0.010 | 0.44 [0.14-1.39] | 0.162 | 0.23 [0.06-0.95] | 0.043 | 1.14 [0.41-3.16] | 0.796 |
| Metformin (Yes) | 0.43 [0.38-0.50] | <0.001 | 0.57 [0.48-0.68] | <0.001 | 0.61 [0.47-0.80] | <0.001 | 0.51 [0.40-0.66] | <0.001 | 0.40 [0.26-0.59] | <0.001 |
| Nitrates (No) | 0.40 [0.34-0.47] | <0.001 | 0.51 [0.42-0.63] | <0.001 | 0.49 [0.35-0.68] | <0.001 | 0.50 [0.37-0.67] | <0.001 | 0.40 [0.26-0.64] | <0.001 |
| Nitrates (Yes) | 0.52 [0.40-0.69] | <0.001 | 0.77 [0.55-1.06] | 0.111 | 0.93 [0.60-1.43] | 0.744 | 0.41 [0.25-0.68] | <0.001 | 0.49 [0.25-0.95] | 0.034 |
| Non-steroidal anti-inflammatory drugs (No) | 0.41 [0.33-0.50] | <0.001 | 0.56 [0.44-0.70] | <0.001 | 0.39 [0.25-0.61] | <0.001 | 0.49 [0.34-0.71] | <0.001 | 0.46 [0.27-0.79] | 0.005 |
| Non-steroidal anti-inflammatory drugs (Yes) | 0.43 [0.35-0.52] | <0.001 | 0.58 [0.46-0.74] | <0.001 | 0.80 [0.58-1.11] | 0.187 | 0.44 [0.31-0.62] | <0.001 | 0.37 [0.22-0.64] | <0.001 |
| Statins and fibrates (No) | 0.31 [0.22-0.43] | <0.001 | 0.38 [0.25-0.58] | <0.001 | 0.35 [0.18-0.68] | 0.002 | 0.48 [0.28-0.85] | 0.011 | 0.43 [0.19-0.97] | 0.041 |
| Statins and fibrates (Yes) | 0.46 [0.39-0.54] | <0.001 | 0.60 [0.50-0.73] | <0.001 | 0.70 [0.52-0.93] | 0.013 | 0.49 [0.37-0.65] | <0.001 | 0.44 [0.29-0.67] | <0.001 |
| Sulphonylurea (No) | 0.28 [0.21-0.38] | <0.001 | 0.42 [0.30-0.60] | <0.001 | 0.51 [0.29-0.87] | 0.013 | 0.20 [0.09-0.42] | <0.001 | 0.13 [0.03-0.50] | 0.003 |
| Sulphonylurea (Yes) | 0.49 [0.42-0.58] | <0.001 | 0.61 [0.50-0.74] | <0.001 | 0.64 [0.47-0.86] | 0.003 | 0.60 [0.46-0.79] | <0.001 | 0.54 [0.37-0.80] | 0.002 |
| Thiozolidinedone (No) | 0.40 [0.34-0.47] | <0.001 | 0.50 [0.42-0.61] | <0.001 | 0.60 [0.45-0.80] | <0.001 | 0.46 [0.35-0.61] | <0.001 | 0.42 [0.28-0.64] | <0.001 |
| Thiozolidinedone (Yes) | 0.77 [0.54-1.09] | 0.145 | 1.01 [0.70-1.46] | 0.956 | 0.63 [0.32-1.24] | 0.183 | 0.71 [0.40-1.26] | 0.237 | 0.55 [0.22-1.34] | 0.188 |

ACEI: angiotensin-converting enzyme inhibitor; ARB: angiotensin receptor blocker; CI: confidence interval; DPP4i: dipeptidyl peptidase-4 inhibitor; GLP-1RA: glucagon-like peptide-1 receptor agonist; HR: hazard ratio; SGLT2i: sodium-glucose cotransporter-2 inhibitor.

Table S8. Sensitivity analyses for competing risk and alternative propensity score approaches for exposure effects of SGLT2i vs. DPP4i.

| **Model** | **New-onset syncope** | | **Cardiovascular mortality** | | **All-cause mortality** | |
| --- | --- | --- | --- | --- | --- | --- |
|  | **HR [95% CI]** | **P value** | **HR [95% CI]** | **P value** | **HR [95% CI]** | **P value** |
| **Cause-specific hazard models** | 0.29 [0.21-0.35] | <0.001 | 0.31 [0.22-0.44] | <0.001 | 0.53 [0.45-0.63] | <0.001 |
| **Sub-distribution hazard models** | 0.35 [0.28-0.52] | <0.001 | 0.42 [0.31-0.63] | <0.001 | 0.66 [0.45-0.83] | <0.001 |
| **PS stratification** | 0.32 [0.25-0.53] | <0.001 | 0.42 [0.32-0.54] | <0.001 | 0.61 [0.48-0.82] | <0.001 |
| **PS with IPTW** | 0.39 [0.34-0.45] | <0.001 | 0.43 [0.34-0.75] | <0.001 | 0.54 [0.44-0.89] | <0.001 |
| **PS with SIPTW** | 0.49 [0.42-0.63] | <0.001 | 0.55 [0.34-0.82] | <0.001 | 0.43 [0.32-0.78] | <0.001 |

CI: confidence interval; DPP4i: dipeptidyl peptidase-4 inhibitor; HR: hazard ratio; IPTW: inverse probability of treatment weighting; PS: propensity score; SGLT2i: sodium-glucose cotransporter-2 inhibitor; SIPTW: stable inverse probability of treatment weighting.

Table S9. Sensitivity analyses for the exposure effects of SGLT2i vs. DPP4i on new-onset syncope after excluding those with baseline immune-mediated inflammatory diseases and cancer.

| **Model** | **New-onset syncope** | |
| --- | --- | --- |
|  | **HR [95% CI]** | **P value** |
| **Univariate analysis** |  |  |
| Before PS matching | 0.45 [0.28-0.56] | <0.001 |
| After PS matching | 0.34 [0.29-0.62] | <0.001 |
| **Multivariate adjustment analysis** |  |  |
| **Model 1** |  |  |
| SGLT2i vs. DPP4i | 0.35 [0.24-0.65] | <0.001 |
| **Model 2** |  |  |
| SGLT2i vs. DPP4i | 0.39 [0.23-0.61] | <0.001 |
| **Model 3** |  |  |
| SGLT2i vs. DPP4i | 0.41 [0.35-0.59] | <0.001 |
| **Model 4** |  |  |
| SGLT2i vs. DPP4i | 0.43 [0.29-0.55] | <0.001 |
| **Model 5** |  |  |
| SGLT2i vs. DPP4i | 0.42 [0.37-0.62] | <0.001 |

Model 1 adjusted for significant demographics.

Model 2 adjusted for significant demographics, prior immune-mediated inflammatory diseases, and cancer.

Model 3 adjusted for significant demographics, and other past comorbidities.

Model 4 adjusted for significant demographics, past comorbidities, and non-SGLT2i/DPP4i medications.

Model 5 adjusted for significant demographics, past comorbidities, non-SGLT2i/DPP4i medications, abbreviated MDRD, fasting glucose, HbA1c, and duration from earliest diabetes mellitus date to initial drug exposure date.

CI: confidence interval; DPP4i: dipeptidyl peptidase-4 inhibitor; HbA1c: hemoglobin A1c; HR: hazard ratio; MDRD: Modification of Diet in Renal Disease; PS: propensity score; SGLT2i: sodium-glucose cotransporter-2 inhibitor.

Table S10. Sensitivity analysis with consideration of 1-year lag time effects for the exposure effects of SGLT2i vs. DPP4i.

| **Characteristics** | **New-onset syncope** | | **Cardiovascular mortality** | | **All-cause mortality** | |
| --- | --- | --- | --- | --- | --- | --- |
|  | **HR [95% CI]** | **P value** | **HR [95% CI]** | **P value** | **HR [95% CI]** | **P value** |
| SGLT2i vs. DPP4i | 0.43 [0.37-0.49] | <0.001 | 0.23 [0.18-0.28] | <0.001 | 0.24 [0.22-0.27] | <0.001 |
| Dapagliflozin vs. DPP4i | 0.56 [0.47-0.66] | <0.001 | 0.30 [0.23-0.41] | <0.001 | 0.32 [0.28-0.36] | <0.001 |
| Empagliflozin vs. DPP4i | 0.60 [0.46-0.78] | <0.001 | 0.26 [0.15-0.44] | <0.001 | 0.31 [0.25-0.38] | <0.001 |
| Canagliflozin vs. DPP4i | 0.49 [0.38-0.64] | <0.001 | 0.30 [0.19-0.46] | <0.001 | 0.36 [0.30-0.43] | <0.001 |
| Ertugliflozin vs. DPP4i | 0.44 [0.30-0.64] | <0.001 | 0.42 [0.25-0.72] | 0.002 | 0.42 [0.33-0.53] | <0.001 |

CI: confidence interval; DPP4i: dipeptidyl peptidase-4 inhibitor; HR: hazard ratio; SGLT2i: sodium-glucose cotransporter-2 inhibitor.

Table S11. Sensitivity analysis for the exposure effects of SGLT2i vs. DPP4i after excluding patients with CKD stage 4/5, peritoneal dialysis or haemodialysis.

| **Characteristics** | **New-onset syncope** | | **Cardiovascular mortality** | | **All-cause mortality** | |
| --- | --- | --- | --- | --- | --- | --- |
|  | **HR [95% CI]** | **P value** | **HR [95% CI]** | **P value** | **HR [95% CI]** | **P value** |
| SGLT2i vs. DPP4i | 0.47 [0.41-0.55] | <0.001 | 0.23 [0.18-0.29] | <0.001 | 0.25 [0.22-0.28] | <0.001 |
| Dapagliflozin vs. DPP4i | 0.60 [0.51-0.72] | <0.001 | 0.31 [0.22-0.42] | <0.001 | 0.33 [0.29-0.38] | <0.001 |
| Empagliflozin vs. DPP4i | 0.70 [0.53-0.91] | 0.009 | 0.26 [0.14-0.47] | <0.001 | 0.29 [0.22-0.37] | <0.001 |
| Canagliflozin vs. DPP4i | 0.54 [0.42-0.70] | <0.001 | 0.28 [0.17-0.46] | <0.001 | 0.36 [0.29-0.44] | <0.001 |
| Ertugliflozin vs. DPP4i | 0.46 [0.31-0.69] | <0.001 | 0.44 [0.25-0.78] | 0.005 | 0.43 [0.33-0.56] | <0.001 |

CKD stage 4/5 indicates eGFR <30 ml/min/1.73 m^2^.

CI: confidence interval; CKD: Chronic kidney disease; DPP4i: dipeptidyl peptidase-4 inhibitor; eGFR: estimated glomerular filtration rate; HR: hazard ratio; SGLT2i: sodium-glucose cotransporter-2 inhibitor.

Table S12. Univariate Cox regression models with adjustments to predict mortality outcomes before and after propensity score matching.

| **Characteristics** | **Before matching** | |  | | **After matching** | |  | |
| --- | --- | --- | --- | --- | --- | --- | --- | --- |
|  | **All-cause mortality** | | **Cardiovascular mortality** | | **All-cause mortality** | | **Cardiovascular mortality** | |
|  | **HR [95% CI]** | **P value** | **HR [95% CI]** | **P value** | **HR [95% CI]** | **P value** | **HR [95% CI]** | **P value** |
| ***Demographics*** |  |  |  |  |  |  |  |  |
| Male | 1.02 [0.93-1.12] | 0.666 | 0.92 [0.87-0.97] | 0.001 | 1.11 [0.94-1.32] | 0.218 | 1.22 [1.12-1.33] | <0.001 |
| Female | 1.0 [Reference] |  | 1.0 [Reference] |  | 1.0 [Reference] |  | 1.0 [Reference] |  |
| Baseline age (years) | 1.13 [1.13-1.14] | <0.001 | 1.09 [1.09-1.10] | <0.001 | 1.13 [1.12-1.14] | <0.001 | 1.09 [1.08-1.09] | <0.001 |
| 18-50 | 0.06 [0.04-0.10] | <0.001 | 0.16 [0.14-0.19] | <0.001 | 0.06 [0.03-0.12] | <0.001 | 0.26 [0.22-0.31] | <0.001 |
| 50-60 | 0.13 [0.11-0.16] | <0.001 | 0.26 [0.24-0.28] | <0.001 | 0.18 [0.13-0.24] | <0.001 | 0.41 [0.37-0.46] | <0.001 |
| 60-70 | 1.0 [Reference] |  | 1.0 [Reference] |  | 1.0 [Reference] |  | 1.0 [Reference] |  |
| 70-80 | 1.80 [1.62-2.00] | <0.001 | 1.96 [1.86-2.07] | <0.001 | 3.86 [3.22-4.64] | <0.001 | 3.33 [3.04-3.65] | <0.001 |
| >80 | 11.58 [10.55-12.72] | <0.001 | 6.78 [6.44-7.14] | <0.001 | 12.20 [10.01-14.86] | <0.001 | 6.93 [6.19-7.76] | <0.001 |
| ***Prior comorbidities*** |  |  |  |  |  |  |  |  |
| Charlson’s standard comorbidity index | 1.61 [1.58-1.64] | <0.001 | 1.55 [1.53-1.56] | <0.001 | 1.74 [1.68-1.80] | <0.001 | 1.60 [1.57-1.63] | <0.001 |
| Duration from earliest diabetes mellitus diagnosis date to baseline date (day) | 1.03 [1.02-1.04] | <0.001 | 1.02 [1.02-1.03] | <0.001 | 1.03 [1.01-1.05] | 0.003 | 1.01 [1.01-1.02] | 0.001 |
| Number of hospitalizations | 1.20 [1.16-1.24] | <0.001 | 1.20 [1.17-1.22] | <0.001 | 1.14 [1.07-1.22] | <0.001 | 1.20 [1.17-1.23] | <0.001 |
| Diabetic retinopathy | 1.39 [1.19-1.63] | <0.001 | 1.73 [1.60-1.88] | <0.001 | 2.16 [1.69-2.75] | <0.001 | 1.59 [1.39-1.81] | <0.001 |
| Diabetic nephropathy | 4.16 [3.33-5.21] | <0.001 | 3.79 [3.33-4.30] | <0.001 | 3.51 [1.93-6.37] | <0.001 | 2.90 [2.11-3.98] | <0.001 |
| Diabetic neuropathy | 1.50 [1.16-1.94] | 0.002 | 1.86 [1.64-2.11] | <0.001 | 1.42 [0.89-2.28] | 0.142 | 2.18 [1.81-2.63] | <0.001 |
| Gout | 3.16 [2.64-3.78] | <0.001 | 2.65 [2.38-2.94] | <0.001 | 2.20 [1.42-3.40] | <0.001 | 2.07 [1.67-2.58] | <0.001 |
| Heart failure | 2.34 [2.02-2.71] | <0.001 | 2.64 [2.44-2.85] | <0.001 | 2.38 [1.80-3.16] | <0.001 | 2.36 [2.06-2.71] | <0.001 |
| Hyperlipidaemia | 1.11 [1.01-1.22] | 0.035 | 1.07 [1.02-1.13] | 0.009 | 1.02 [0.86-1.21] | 0.785 | 0.96 [0.88-1.04] | 0.282 |
| Hypertension | 2.09 [1.91-2.30] | <0.001 | 2.02 [1.92-2.12] | <0.001 | 2.07 [1.74-2.45] | <0.001 | 2.21 [2.04-2.40] | <0.001 |
| Hypoglycemia | 3.88 [2.90-5.18] | <0.001 | 3.38 [2.85-4.00] | <0.001 | 2.49 [0.80-7.75] | 0.115 | 2.72 [1.61-4.60] | <0.001 |
| IHD with AMI | 2.03 [1.62-2.55] | <0.001 | 2.23 [1.98-2.51] | <0.001 | 3.02 [2.19-4.15] | <0.001 | 2.85 [2.43-3.33] | <0.001 |
| IHD without AMI | 1.32 [1.12-1.56] | 0.001 | 1.39 [1.27-1.52] | <0.001 | 2.08 [1.65-2.64] | <0.001 | 1.67 [1.48-1.89] | <0.001 |
| Liver diseases | 0.97 [0.75-1.25] | 0.808 | 1.18 [1.04-1.34] | 0.008 | 0.54 [0.30-0.95] | 0.033 | 0.90 [0.72-1.12] | 0.336 |
| COPD | 0.03 [0.00-0.25] | <0.001 | 0.07 [0.03-0.15] | <0.001 | 0.36 [0.05-2.55] | 0.306 | 0.17 [0.04-0.67] | 0.012 |
| Peripheral vascular disease | 3.28 [2.37-4.54] | <0.001 | 3.87 [3.29-4.56] | <0.001 | 4.08 [2.18-7.63] | <0.001 | 4.11 [3.04-5.56] | <0.001 |
| Renal diseases | 4.04 [3.31-4.92] | <0.001 | 4.41 [3.98-4.89] | <0.001 | 5.49 [3.16-9.51] | <0.001 | 4.94 [3.73-6.53] | <0.001 |
| Stroke/TIA | 2.95 [2.49-3.50] | <0.001 | 2.30 [2.08-2.55] | <0.001 | 2.90 [2.07-4.07] | <0.001 | 2.13 [1.76-2.57] | <0.001 |
| Atrial fibrillation | 4.01 [3.38-4.76] | <0.001 | 3.20 [2.88-3.54] | <0.001 | 4.19 [3.03-5.78] | <0.001 | 3.53 [2.99-4.18] | <0.001 |
| Anemia | 3.60 [3.12-4.14] | <0.001 | 3.17 [2.93-3.44] | <0.001 | 2.88 [2.05-4.06] | <0.001 | 2.20 [1.83-2.65] | <0.001 |
| Cancer | 2.32 [1.90-2.84] | <0.001 | 2.34 [2.09-2.61] | <0.001 | 3.73 [2.67-5.21] | <0.001 | 2.42 [1.99-2.94] | <0.001 |
| ***Medications*** |  |  |  |  |  |  |  |  |
| SGLT2i vs. DPP4i | 0.10 [0.08-0.12] | <0.001 | 0.15 [0.14-0.17] | <0.001 | 0.19 [0.15-0.24] | <0.001 | 0.25 [0.23-0.28] | <0.001 |
| Dapagliflozin vs. DPP4i | 0.12 [0.09-0.16] | <0.001 | 0.19 [0.17-0.21] | <0.001 | 0.27 [0.20-0.35] | <0.001 | 0.33 [0.29-0.38] | <0.001 |
| Empagliflozin vs. DPP4i | 0.10 [0.06-0.17] | <0.001 | 0.17 [0.14-0.22] | <0.001 | 0.23 [0.13-0.39] | <0.001 | 0.32 [0.26-0.40] | <0.001 |
| Canagliflozin vs. DPP4i | 0.12 [0.08-0.18] | <0.001 | 0.20 [0.17-0.24] | <0.001 | 0.26 [0.17-0.41] | <0.001 | 0.37 [0.31-0.44] | <0.001 |
| Ertugliflozin vs. DPP4i | 0.16 [0.10-0.27] | <0.001 | 0.24 [0.19-0.30] | <0.001 | 0.37 [0.22-0.63] | <0.001 | 0.44 [0.34-0.55] | <0.001 |
| DPP4i frequency | 0.99 [0.99-1.00] | 0.096 | 0.99 [0.99-1.00] | <0.001 | 1.00 [0.99-1.01] | 0.795 | 1.02 [1.01-1.02] | <0.001 |
| SGLT2i duration (days) | 1.000 [0.999-1.000] | 0.161 | 1.000 [1.000-1.000] | 0.008 | 1.000 [0.999-1.000] | 0.161 | 1.000 [1.000-1.000] | 0.008 |
| DPP4i duration (days) | 0.999 [0.999-0.999] | <0.001 | 0.999 [0.999-0.999] | <0.001 | 1.000 [0.999-1.000] | 0.007 | 0.999 [0.999-0.999] | <0.001 |
| Number of anti-diabetic drugs | 0.99 [0.99-1.00] | <0.001 | 1.00 [0.99-1.00] | <0.001 | 0.99 [0.99-1.00] | 0.004 | 1.00 [0.99-1.00] | <0.001 |
| Metformin | 0.26 [0.24-0.29] | <0.001 | 0.27 [0.26-0.29] | <0.001 | 0.34 [0.27-0.42] | <0.001 | 0.44 [0.39-0.50] | <0.001 |
| Sulphonylurea | 1.24 [1.10-1.39] | <0.001 | 1.00 [0.94-1.06] | 0.984 | 1.32 [1.08-1.62] | 0.007 | 1.14 [1.04-1.25] | 0.006 |
| Insulin | 7.90 [6.84-9.12] | <0.001 | 5.26 [4.91-5.62] | <0.001 | 7.79 [5.92-10.25] | <0.001 | 4.21 [3.79-4.69] | <0.001 |
| Acarbose | 1.04 [0.82-1.33] | 0.737 | 1.18 [1.04-1.34] | 0.010 | 0.88 [0.55-1.41] | 0.599 | 1.09 [0.89-1.34] | 0.390 |
| Thiozolidinedone | 0.24 [0.20-0.30] | <0.001 | 0.36 [0.33-0.39] | <0.001 | 0.35 [0.27-0.46] | <0.001 | 0.40 [0.35-0.45] | <0.001 |
| Glucagon-like peptide-1 receptor agonists | 1.14 [0.93-1.39] | 0.200 | 1.03 [0.92-1.16] | 0.561 | 0.14 [0.06-0.34] | <0.001 | 0.31 [0.23-0.41] | <0.001 |
| ACEI/ARBs | 1.40 [1.28-1.54] | <0.001 | 1.40 [1.33-1.47] | <0.001 | 1.40 [1.18-1.67] | <0.001 | 1.38 [1.27-1.50] | <0.001 |
| Antihepatitis | 1.15 [0.81-1.62] | 0.433 | 1.29 [1.08-1.54] | 0.005 | 0.39 [0.15-1.05] | 0.063 | 1.45 [1.13-1.87] | 0.004 |
| Anticoagulants | 1.64 [1.49-1.80] | <0.001 | 1.60 [1.52-1.68] | <0.001 | 2.02 [1.70-2.40] | <0.001 | 1.94 [1.78-2.10] | <0.001 |
| Antiplatelets | 1.56 [1.42-1.72] | <0.001 | 1.60 [1.52-1.69] | <0.001 | 3.04 [2.56-3.61] | <0.001 | 2.22 [2.05-2.41] | <0.001 |
| Lipid-lowering drugs | 1.18 [1.08-1.30] | <0.001 | 1.10 [1.05-1.16] | <0.001 | 1.24 [1.04-1.48] | 0.016 | 1.14 [1.04-1.24] | 0.003 |
| Statins and fibrates | 0.56 [0.51-0.61] | <0.001 | 0.60 [0.57-0.63] | <0.001 | 0.97 [0.80-1.17] | 0.728 | 0.94 [0.86-1.03] | 0.196 |
| Nitrates | 1.35 [1.19-1.54] | <0.001 | 1.51 [1.41-1.62] | <0.001 | 1.65 [1.33-2.05] | <0.001 | 2.31 [2.10-2.54] | <0.001 |
| Non-steroidal anti-inflammatory drugs | 1.58 [1.43-1.75] | <0.001 | 1.46 [1.38-1.54] | <0.001 | 3.54 [2.98-4.21] | <0.001 | 2.20 [2.03-2.39] | <0.001 |
| Diuretics | 1.77 [1.61-1.95] | <0.001 | 1.81 [1.72-1.91] | <0.001 | 2.71 [2.28-3.21] | <0.001 | 2.79 [2.57-3.03] | <0.001 |
| Beta-blockers | 1.52 [1.37-1.69] | <0.001 | 1.58 [1.49-1.67] | <0.001 | 2.28 [1.92-2.71] | <0.001 | 2.40 [2.20-2.61] | <0.001 |
| Calcium channel blockers | 1.36 [1.23-1.49] | <0.001 | 1.23 [1.17-1.30] | <0.001 | 1.83 [1.54-2.17] | <0.001 | 1.45 [1.34-1.58] | <0.001 |
| Anti-cancer drugs | 1.38 [1.19-1.60] | <0.001 | 1.67 [1.55-1.80] | <0.001 | 1.18 [0.81-1.71] | 0.390 | 1.59 [1.35-1.86] | <0.001 |
| Steroids/Corticosteroids | 1.98 [1.61-2.43] | <0.001 | 1.71 [1.52-1.93] | <0.001 | 0.44 [0.14-1.36] | 0.153 | 0.61 [0.39-0.97] | 0.038 |
| ***Subclinical biomarker*** |  |  |  |  |  |  |  |  |
| Abbreviated MDRD (ml/min/1.73m^2^) | 0.970 [0.968-0.972] | <0.001 | 0.970 [0.969-0.971] | <0.001 | 0.97 [0.96-0.97] | <0.001 | 0.973 [0.971-0.975] | <0.001 |
| Most severe renal damage (<15ml/min/1.73m^2^) | 4.02 [3.09-5.22] | <0.001 | 5.36 [4.73-6.08] | <0.001 | 14.84 [9.37-23.49] | <0.001 | 8.30 [6.16-11.20] | <0.001 |
| Severe renal damage ([15, 30) ml/min/1.73m^2^) | 4.75 [4.04-5.58] | <0.001 | 5.11 [4.69-5.57] | <0.001 | 6.32 [4.34-9.20] | <0.001 | 5.44 [4.46-6.64] | <0.001 |
| Moderate to severe renal damage ([30, 45) ml/min/1.73m^2^) | 3.25 [2.87-3.67] | <0.001 | 3.19 [2.99-3.42] | <0.001 | 3.67 [2.83-4.75] | <0.001 | 3.13 [2.74-3.59] | <0.001 |
| Mild to moderate renal damage ([45, 60) ml/min/1.73m^2^) | 2.03 [1.79-2.29] | <0.001 | 1.81 [1.69-1.94] | <0.001 | 1.80 [1.41-2.31] | <0.001 | 2.23 [1.99-2.50] | <0.001 |
| Mild renal damage ([60, 90] ml/min/1.73m^2^) | 0.69 [0.62-0.77] | <0.001 | 0.64 [0.61-0.68] | <0.001 | 1.10 [0.91-1.32] | 0.316 | 0.87 [0.80-0.96] | 0.004 |
| Chronic kidney disease (>90 ml/min/1.73m^2^) | 0.21 [0.18-0.25] | <0.001 | 0.25 [0.23-0.27] | <0.001 | 0.23 [0.18-0.30] | <0.001 | 0.38 [0.34-0.42] | <0.001 |
| ***Complete blood counts; mean (SD)*** |  |  |  |  |  |  |  |  |
| Mean corpuscular volume (fL) | 1.02 [1.01-1.03] | <0.001 | 1.02 [1.02-1.03] | <0.001 | 1.02 [1.00-1.03] | 0.050 | 1.02 [1.01-1.02] | <0.001 |
| Eosinophil (x10^9^/L) | 0.92 [0.70-1.22] | 0.576 | 1.06 [0.94-1.19] | 0.363 | 0.71 [0.38-1.34] | 0.294 | 0.98 [0.74-1.29] | 0.874 |
| Lymphocyte (x10^9^/L) | 0.48 [0.44-0.53] | <0.001 | 0.52 [0.50-0.55] | <0.001 | 0.46 [0.39-0.55] | <0.001 | 0.52 [0.48-0.56] | <0.001 |
| Neutrophil (x10^9^/L) | 1.07 [1.05-1.08] | <0.001 | 1.05 [1.05-1.06] | <0.001 | 1.05 [1.02-1.09] | 0.002 | 1.07 [1.06-1.09] | <0.001 |
| White cell count (x10^9^/L) | 1.02 [1.01-1.03] | <0.001 | 1.02 [1.01-1.02] | <0.001 | 1.06 [1.03-1.09] | <0.001 | 1.06 [1.04-1.07] | <0.001 |
| Mean cell haemoglobin (pg) | 1.05 [1.03-1.07] | <0.001 | 1.06 [1.05-1.07] | <0.001 | 1.03 [0.99-1.06] | 0.183 | 1.04 [1.02-1.06] | <0.001 |
| Platelet (x10^9^/L) | 0.998 [0.997-0.998] | <0.001 | 0.997 [0.997-0.998] | <0.001 | 0.99 [0.99-1.00] | <0.001 | 1.00 [0.99-1.00] | <0.001 |
| Red cell count (x10^12^/L) | 0.37 [0.35-0.40] | <0.001 | 0.39 [0.37-0.40] | <0.001 | 0.27 [0.24-0.31] | <0.001 | 0.43 [0.40-0.46] | <0.001 |
| ***Liver and renal functions; mean (SD)*** |  |  |  |  |  |  |  |  |
| Potassium (mmol/L) | 1.14 [1.03-1.27] | 0.013 | 1.13 [1.07-1.20] | <0.001 | 1.44 [1.18-1.75] | <0.001 | 1.21 [1.10-1.34] | <0.001 |
| Albumin (g/L) | 0.85 [0.84-0.86] | <0.001 | 0.86 [0.86-0.87] | <0.001 | 0.83 [0.82-0.85] | <0.001 | 0.84 [0.84-0.85] | <0.001 |
| Sodium (mmol/L) | 0.95 [0.93-0.96] | <0.001 | 0.95 [0.94-0.96] | <0.001 | 1.03 [0.99-1.06] | 0.141 | 0.94 [0.93-0.96] | <0.001 |
| Urea (mmol/L) | 1.09 [1.08-1.09] | <0.001 | 1.090 [1.086-1.093] | <0.001 | 1.12 [1.11-1.14] | <0.001 | 1.11 [1.10-1.12] | <0.001 |
| Protein (g/L) | 0.94 [0.93-0.95] | <0.001 | 0.95 [0.94-0.95] | <0.001 | 0.96 [0.94-0.97] | <0.001 | 0.96 [0.95-0.97] | <0.001 |
| Creatinine (umol/L) | 1.002 [1.002-1.003] | <0.001 | 1.003 [1.002-1.003] | <0.001 | 1.003 [1.003-1.004] | <0.001 | 1.003 [1.003-1.004] | <0.001 |
| Alkaline phosphatase (U/L) | 1.00 [1.00-1.01] | <0.001 | 1.00 [1.00-1.01] | <0.001 | 1.01 [1.00-1.01] | <0.001 | 1.00 [1.00-1.01] | <0.001 |
| Aspartate transaminase (U/L) | 1.000 [0.999-1.002] | 0.866 | 1.000 [1.000-1.001] | 0.386 | 1.00 [1.00-1.01] | 0.639 | 1.002 [1.000-1.003] | 0.063 |
| Alanine transaminase (U/L) | 0.98 [0.97-0.98] | <0.001 | 0.988 [0.986-0.990] | <0.001 | 0.99 [0.98-0.99] | <0.001 | 0.999 [0.997-1.001] | 0.188 |
| Bilirubin (umol/L) | 0.96 [0.95-0.97] | <0.001 | 0.99 [0.99-1.00] | 0.002 | 0.94 [0.91-0.96] | <0.001 | 1.01 [1.00-1.01] | <0.001 |
| ***Lipid profile and variabilities; mean (SD)*** |  |  |  |  |  |  |  |  |
| Triglyceride (mmol/L) | 0.90 [0.85-0.95] | <0.001 | 0.94 [0.92-0.97] | <0.001 | 0.89 [0.81-0.98] | 0.014 | 0.93 [0.89-0.97] | <0.001 |
| SD of triglyceride | 0.94 [0.84-1.05] | 0.270 | 0.93 [0.88-0.99] | 0.021 | 0.72 [0.51-1.00] | 0.049 | 0.93 [0.85-1.01] | 0.085 |
| Low-density lipoprotein (mmol/L) | 0.89 [0.83-0.96] | 0.002 | 0.95 [0.91-0.98] | 0.006 | 0.81 [0.71-0.93] | 0.002 | 0.94 [0.89-1.00] | 0.061 |
| SD of low-density lipoprotein | 1.20 [0.95-1.51] | 0.124 | 1.21 [1.08-1.36] | 0.001 | 1.16 [0.74-1.82] | 0.506 | 1.33 [1.11-1.60] | 0.002 |
| High-density lipoprotein (mmol/L) | 1.34 [1.14-1.57] | <0.001 | 1.10 [1.00-1.20] | 0.042 | 1.13 [0.86-1.49] | 0.361 | 1.22 [1.08-1.39] | 0.002 |
| SD of high-density lipoprotein | 22.08 [11.82-41.26] | <0.001 | 15.14 [10.75-21.31] | <0.001 | 18.46 [5.85-58.27] | <0.001 | 13.27 [7.79-22.60] | <0.001 |
| Total cholesterol (mmol/L) | 0.90 [0.85-0.96] | 0.001 | 0.94 [0.91-0.97] | <0.001 | 0.83 [0.75-0.93] | <0.001 | 0.93 [0.89-0.98] | 0.008 |
| SD of total cholesterol | 1.29 [1.10-1.52] | 0.002 | 1.24 [1.13-1.35] | <0.001 | 1.19 [0.85-1.65] | 0.314 | 1.27 [1.12-1.45] | <0.001 |
| ***Glucose tests and variabilities; mean (SD)*** |  |  |  |  |  |  |  |  |
| Hemoglobin A1C (%) | 0.91 [0.88-0.95] | <0.001 | 0.95 [0.93-0.97] | <0.001 | 0.86 [0.80-0.92] | <0.001 | 0.99 [0.96-1.02] | 0.633 |
| Mean hemoglobin A1C (%) | 0.90 [0.86-0.94] | <0.001 | 0.95 [0.92-0.97] | <0.001 | 0.85 [0.78-0.92] | <0.001 | 0.97 [0.94-1.00] | 0.084 |
| Variance of hemoglobin A1C | 1.00 [1.00-1.01] | 0.009 | 1.00 [1.00-1.01] | <0.001 | 1.00 [0.97-1.02] | 0.754 | 1.00 [1.00-1.01] | 0.568 |
| SD of hemoglobin A1C | 1.15 [1.11-1.20] | <0.001 | 1.14 [1.12-1.17] | <0.001 | 1.06 [0.94-1.19] | 0.313 | 1.11 [1.07-1.16] | <0.001 |
| CV of hemoglobin A1C | 1.04 [1.03-1.05] | <0.001 | 1.04 [1.03-1.04] | <0.001 | 1.02 [1.00-1.04] | 0.064 | 1.03 [1.02-1.04] | <0.001 |
| Fasting glucose (mmol/L) | 1.03 [1.02-1.04] | <0.001 | 1.02 [1.01-1.03] | <0.001 | 0.97 [0.94-1.00] | 0.062 | 1.02 [1.01-1.03] | <0.001 |
| Mean fasting glucose (mmol/L) | 1.05 [1.03-1.07] | <0.001 | 1.04 [1.03-1.05] | <0.001 | 1.00 [0.96-1.03] | 0.913 | 1.03 [1.01-1.04] | <0.001 |
| Variance of fasting glucose | 1.003 [1.002-1.003] | <0.001 | 1.003 [1.002-1.003] | <0.001 | 1.00 [1.00-1.01] | <0.001 | 1.003 [1.002-1.004] | <0.001 |
| SD of fasting glucose | 1.12 [1.10-1.14] | <0.001 | 1.11 [1.10-1.12] | <0.001 | 1.11 [1.07-1.15] | <0.001 | 1.11 [1.09-1.12] | <0.001 |
| CV of fasting glucose | 1.03 [1.02-1.03] | <0.001 | 1.03 [1.02-1.03] | <0.001 | 1.03 [1.02-1.03] | <0.001 | 1.02 [1.02-1.03] | <0.001 |
| SGLT2i vs. DPP4i | 0.10 [0.08-0.12] | <0.001 | 0.15 [0.14-0.17] | <0.001 | 0.19 [0.15-0.24] | <0.001 | 0.25 [0.23-0.28] | <0.001 |

ACEI: angiotensin-converting enzyme inhibitor; AMI: acute myocardial infarction; ARB: angiotensin receptor blocker; CI: confidence interval; COPD: chronic obstructive pulmonary disease; CV: coefficient of variation; DPP4i: dipeptidyl peptidase-4 inhibitor; HR: hazard ratio; IHD: ischemic heart disease; MDRD: Modification of Diet in Renal Disease; SD: standard deviation; SGLT2i: sodium glucose cotransporter-2 inhibitor; TIA: transient ischemic attack.

Table S13. Sensitivity analysis for the exposure effects of SGLT2i vs. DPP4i on cardiovascular mortality in subgroups of HbA1c and fasting glucose.

| **Subgroup** | **No. patients** | **No. SGLT2i (%)** | **Cardiovascular mortality**  **in SGLT2i (%)** | **No. DPP4i (%)** | **Cardiovascular mortality**  **in DPP4i(%)** | **SGLT2i vs. DPP4i** | | **Dapagliflozin vs. DPP4i** | | **Empagliflozin vs. DPP4i** | | **Canagliflozin vs. DPP4i** | | **Ertugliflozin vs. DPP4i** | |
| --- | --- | --- | --- | --- | --- | --- | --- | --- | --- | --- | --- | --- | --- | --- | --- |
|  |  |  |  |  |  | **HR**  **[95% CI]** | **P value** | **HR**  **[95% CI]** | **P**  **value** | **HR**  **[95% CI]** | **P value** | **HR**  **[95% CI]** | **P value** | **HR**  **[95% CI]** | **P value** |
| ***Baseline HbA1c (%)*** | | | | | | | | | | | | | | | |
| <7.5 | 11425 | 5247 (45.93) | 16  (0.3) | 6178 (54.07) | 132  (2.14) | 0.14 [0.08-0.23] | <0.001 | 0.17 [0.09-0.36] | <0.001 | 0.2 [0.06-0.63] | 0.006 | 0.21 [0.09-0.52] | <0.001 | 0.19 [0.05-0.76] | 0.019 |
| 7.5-9 | 11973 | 6041 (50.46) | 33  (0.55) | 5932 (49.54) | 109  (1.84) | 0.29 [0.19-0.42] | <0.001 | 0.32 [0.20-0.54] | <0.001 | 0.27 [0.1-0.72] | 0.009 | 0.44 [0.23-0.87] | 0.018 | 0.66 [0.29-1.5] | 0.325 |
| >9 | 7537 | 4220 (55.99) | 23  (0.55) | 3317 (44.01) | 76  (2.29) | 0.23 [0.14-0.36] | <0.001 | 0.32 [0.18-0.57] | <0.001 | 0.26 [0.08-0.84] | 0.023 | 0.26 [0.1-0.71] | 0.008 | 0.57 [0.21-1.54] | 0.265 |
| ***Mean HbA1c (%)*** | | | | | | | | | | | | | | | |
| Q1 | 7533 | 3424 (45.45) | 11  (0.32) | 4109 (54.55) | 84  (2.04) | 0.15 [0.08-0.28] | <0.001 | 0.17 [0.07-0.43] | <0.001 | 0.1 [0.01-0.71] | 0.022 | 0.2 [0.06-0.62] | 0.005 | 0.29 [0.07-1.16] | 0.081 |
| Q2 | 7545 | 3465 (45.92) | 14  (0.4) | 4080 (54.08) | 90  (2.21) | 0.18 [0.1-0.31] | <0.001 | 0.24 [0.11-0.48] | <0.001 | 0.32 [0.1-1.0] | 0.049 | 0.21 [0.07-0.66] | 0.008 | 0.3 [0.07-1.22] | 0.092 |
| Q3 | 7585 | 4037 (53.22) | 19  (0.47) | 3548 (46.78) | 32  (0.9) | 0.5 [0.29-0.89] | 0.018 | 0.42 [0.20-0.89] | 0.023 | 0.36 [0.09-1.46] | 0.152 | 0.96 [0.43-2.14] | 0.927 | 1.14 [0.41-3.17] | 0.796 |
| Q4 | 7558 | 4162 (55.07) | 27  (0.65) | 3396 (44.93) | 87  (2.56) | 0.24 [0.16-0.37] | <0.001 | 0.35 [0.21-0.59] | <0.001 | 0.31 [0.11-0.83] | 0.020 | 0.29 [0.12-0.7] | 0.006 | 0.52 [0.19-1.42] | 0.201 |
| ***Variance of HbA1c*** | | | | | | | | | | | | | | | |
| Q1 | 5751 | 2862 (49.77) | 11  (0.38) | 2889 (50.23) | 28  (0.97) | 0.38 [0.19-0.76] | 0.007 | 0.21 [0.07-0.69] | 0.010 | 0.47 [0.11-1.95] | 0.299 | 0.94 [0.37-2.4] | 0.894 | 1.72 [0.61-4.84] | 0.303 |
| Q2 | 5974 | 3258 (54.54) | 16  (0.49) | 2716 (45.46) | 36  (1.33) | 0.36 [0.2-0.65] | <0.001 | 0.4 [0.19-0.85] | 0.017 | - | - | 0.62 [0.25-1.55] | 0.304 | 1.07 [0.39-2.98] | 0.303 |
| Q3 | 5927 | 3194 (53.89) | 13  (0.41) | 2733 (46.11) | 51  (1.87) | 0.21 [0.11-0.38] | <0.001 | 0.31 [0.15-0.65] | 0.002 | 0.29 [0.07-1.2] | 0.088 | 0.2 [0.05-0.81] | 0.024 | 0.21 [0.03-1.55] | 0.127 |
| Q4 | 5962 | 3158 (52.97) | 22  (0.7) | 2804 (47.03) | 81  (2.89) | 0.23 [0.14-0.37] | <0.001 | 0.35 [0.2-0.61] | <0.001 | 0.36 [0.13-0.97] | 0.043 | 0.24 [0.09-0.65] | 0.005 | 0.3 [0.07-1.2] | 0.089 |
| ***CV of HbA1c*** | | | | | | | | | | | | | | | |
| Q1 | 5759 | 2951 (51.24) | 11  (0.37) | 2808 (48.76) | 31  (1.1) | 0.32 [0.16-0.64] | 0.001 | 0.19 [0.06-0.6] | 0.005 | 0.41 [0.1-1.71] | 0.222 | 0.85 [0.33-2.16] | 0.730 | 1.57 [0.56-4.39] | 0.394 |
| Q2 | 5759 | 3139 (54.51) | 16  (0.51) | 2620 (45.49) | 30  (1.15) | 0.43 [0.24-0.8] | 0.007 | 0.46 [0.22-0.99] | 0.047 | 0.21 [0.03-1.53] | 0.124 | 0.7 [0.28-1.77] | 0.453 | 0.89 [0.28-2.88] | 0.850 |
| Q3 | 5751 | 3133 (54.48) | 17  (0.54) | 2618 (45.52) | 59  (2.25) | 0.23 [0.13-0.39] | <0.001 | 0.33 [0.17-0.64] | 0.001 | 0.23 [0.06-0.95] | 0.042 | 0.33 [0.12-0.92] | 0.033 | 0.35 [0.09-1.42] | 0.142 |
| Q4 | 5769 | 3005 (52.09) | 18  (0.6) | 2764 (47.91) | 75  (2.71) | 0.21 [0.13-0.36] | <0.001 | 0.33 [0.18-0.61] | <0.001 | 0.3 [0.1-0.96] | 0.043 | 0.13 [0.03-0.55] | 0.005 | 0.35 [0.09-1.42] | 0.143 |
| ***Baseline fasting glucose (mmol/L)*** | | | | | | | | | | | | | | | |
| <5.6 | 6669 | 2807 (42.09) | 14  (0.5) | 3862 (57.91) | 130  (3.37) | 0.14 [0.08-0.24] | <0.001 | 0.18 [0.09-0.38] | <0.001 | 0.23 [0.07-0.73] | 0.012 | 0.22 [0.08-0.59] | 0.003 | - | - |
| 5.6-6.9 | 5092 | 2382 (46.78) | 11  (0.46) | 2710 (53.22) | 39  (1.44) | 0.3 [0.15-0.59] | <0.001 | 0.31 [0.12-0.78] | 0.013 | 0.19 [0.03-1.4] | 0.104 | 0.27 [0.07-1.11] | 0.070 | 0.91 [0.28-2.94] | 0.880 |
| >6.9 | 19994 | 10679 (53.41) | 50  (0.47) | 9315 (46.59) | 164  (1.76) | 0.26 [0.19-0.35] | <0.001 | 0.35 [0.24-0.52] | <0.001 | 0.25 [0.11-0.56] | <0.001 | 0.35 [0.2-0.63] | <0.001 | 0.59 [0.3-1.15] | 0.124 |
| ***Mean fasting glucose (mmol/L)*** | | | | | | | | | | | | | | | |
| Q1 | 7111 | 3285 (46.2) | 13  (0.4) | 3826 (53.8) | 62  (1.62) | 0.23 [0.12-0.41] | <0.001 | 0.34 [0.16-0.7] | 0.004 | 0.26 [0.06-1.06] | 0.061 | 0.09 [0.01-0.64] | 0.017 | 0.4 [0.1-1.64] | 0.205 |
| Q2 | 7254 | 3645 (50.25) | 13  (0.36) | 3609 (49.75) | 60  (1.66) | 0.21 [0.11-0.38] | <0.001 | 0.26 [0.12-0.58] | <0.001 | - | - | 0.35 [0.13-0.96] | 0.042 | 0.61 [0.19-1.94] | 0.403 |
| Q3 | 7195 | 3979 (55.3) | 21  (0.53) | 3216 (44.7) | 66  (2.05) | 0.25 [0.15-0.4] | <0.001 | 0.28 [0.15-0.54] | <0.001 | 0.39 [0.14-1.08] | 0.069 | 0.57 [0.28-1.18] | 0.130 | 0.47 [0.15-1.47] | 0.193 |
| Q4 | 7192 | 3837 (53.35) | 25  (0.65) | 3355 (46.65) | 81  (2.41) | 0.26 [0.16-0.4] | <0.001 | 0.39 [0.23-0.66] | <0.001 | 0.24 [0.08-0.76] | 0.015 | 0.18 [0.06-0.58] | 0.004 | 0.55 [0.2-1.51] | 0.247 |
| ***Variance of fasting glucose*** | | | | | | | | | | | | | | | |
| Q1 | 4844 | 2616 (54.0) | 11  (0.42) | 2228 (46.0) | 12  (0.54) | 0.77 [0.34-1.74] | 0.526 | 1.02 [0.42-2.47] | 0.973 | 0.39 [0.05-2.92] | 0.362 | 0.56 [0.13-2.39] | 0.435 | 1.25 [0.29-5.32] | 0.766 |
| Q2 | 4847 | 2888 (59.58) | 9  (0.31) | 1959 (40.42) | 60  (3.06) | 0.1 [0.05-0.2] | <0.001 | 0.18 [0.08-0.42] | <0.001 | 0.23 [0.06-0.94] | 0.040 | 0.07 [0.01-0.53] | 0.010 | 0.18 [0.03-1.33] | 0.094 |
| Q3 | 4844 | 2771 (57.2) | 6  (0.22) | 2073 (42.8) | 20  (0.96) | 0.22 [0.09-0.54] | 0.001 | 0.24 [0.07-0.81] | 0.022 | 0.71 [0.17-3.0] | 0.639 | 0.5 [0.12-2.11] | 0.344 | - | - |
| Q4 | 4847 | 2321 (47.89) | 23  (0.99) | 2526 (52.11) | 91  (3.6) | 0.25 [0.16-0.4] | <0.001 | 0.27 [0.14-0.5] | <0.001 | 0.18 [0.04-0.71] | 0.015 | 0.3 [0.12-0.75] | 0.009 | 1.09 [0.51-2.33] | 0.833 |
| ***CV of fasting glucose*** | | | | | | | | | | | | | | | |
| Q1 | 4723 | 2573 (54.48) | 12  (0.47) | 2150 (45.52) | 12  (0.56) | 0.82 [0.37-1.82] | 0.625 | 1.15 [0.49-2.68] | 0.754 | 0.37 [0.05-2.72] | 0.328 | 0.52 [0.12-2.23] | 0.382 | 1.2 [0.28-5.1] | 0.805 |
| Q2 | 4722 | 2852 (60.4) | 7  (0.25) | 1870 (39.6) | 42  (2.25) | 0.11 [0.05-0.24] | <0.001 | 0.12 [0.04-0.4] | <0.001 | 0.48 [0.15-1.53] | 0.213 | 0.1 [0.01-0.76] | 0.025 | 0.26 [0.04-1.9] | 0.185 |
| Q3 | 4716 | 2680 (56.83) | 10  (0.37) | 2036 (43.17) | 23  (1.13) | 0.32 [0.15-0.67] | 0.003 | 0.42 [0.17-1.02] | 0.056 | 0.27 [0.04-1.98] | 0.197 | 0.39 [0.09-1.63] | 0.196 | 0.95 [0.23-3.97] | 0.945 |
| Q4 | 4730 | 2252 (47.61) | 17  (0.75) | 2478 (52.39) | 93  (3.75) | 0.19 [0.11-0.31] | <0.001 | 0.22 [0.11-0.44] | <0.001 | 0.19 [0.05-0.77] | 0.020 | 0.25 [0.09-0.69] | 0.007 | 0.65 [0.24-1.77] | 0.400 |

CI: confidence interval; CV: coefficient of variation; DPP4i: dipeptidyl peptidase-4 inhibitor; HbA1c: hemoglobin A1c; HR: hazard ratio; SGLT2i: sodium-glucose cotransporter-2 inhibitor.

Table S14. Sensitivity analysis for the exposure effects of SGLT2i vs. DPP4i on all-cause mortality in subgroups of HbA1c and fasting glucose.

| **Subgroup** | **No. patients** | **No. SGLT2i (%)** | **All-cause mortality in SGLT2i (%)** | **No. DPP4i (%)** | **All-cause mortality in DPP4i (%)** | **SGLT2i vs. DPP4i** | | **Dapagliflozin vs. DPP4i** | | **Empagliflozin vs. DPP4i** | | **Canagliflozin vs. DPP4i** | | **Ertugliflozin vs. DPP4i** | |
| --- | --- | --- | --- | --- | --- | --- | --- | --- | --- | --- | --- | --- | --- | --- | --- |
|  |  |  |  |  |  | **HR**  **[95% CI]** | **P value** | **HR**  **[95% CI]** | **P value** | **HR**  **[95% CI]** | **P value** | **HR**  **[95% CI]** | **P**  **value** | **HR**  **[95% CI]** | **P value** |
| ***Baseline HbA1c (%)*** | | | | | | | | | | | | | | | |
| <7.5 | 11425 | 5247 (45.93) | 118  (2.25) | 6178 (54.07) | 628 (10.17) | 0.21 [0.17-0.26] | <0.001 | 0.31 [0.24-0.39] | <0.001 | 0.24 [0.15-0.38] | <0.001 | 0.26 [0.18-0.37] | <0.001 | 0.28 [0.17-0.47] | <0.001 |
| 7.5-9 | 11973 | 6041 (50.46) | 149  (2.47) | 5932 (49.54) | 480 (8.09) | 0.29 [0.24-0.35] | <0.001 | 0.37 [0.29-0.46] | <0.001 | 0.26 [0.16-0.42] | <0.001 | 0.49 [0.36-0.67] | <0.001 | 0.67 [0.46-0.99] | 0.045 |
| >9 | 7537 | 4220 (55.99) | 128  (3.03) | 3317 (44.01) | 434 (13.08) | 0.22 [0.18-0.27] | <0.001 | 0.3 [0.23-0.38] | <0.001 | 0.36 [0.24-0.55] | <0.001 | 0.35 [0.24-0.5] | <0.001 | 0.34 [0.2-0.58] | <0.001 |
| ***Mean HbA1c (%)*** | | | | | | | | | | | | | | | |
| Q1 | 7533 | 3424 (45.45) | 83 (2.42) | 4109 (54.55) | 476 (11.58) | 0.2 [0.16-0.25] | <0.001 | 0.28 [0.21-0.38] | <0.001 | 0.2 [0.12-0.36] | <0.001 | 0.23 [0.15-0.36] | <0.001 | 0.29 [0.17-0.52] | <0.001 |
| Q2 | 7545 | 3465 (45.92) | 71 (2.05) | 4080 (54.08) | 300 (7.35) | 0.27 [0.21-0.35] | <0.001 | 0.41 [0.3-0.56] | <0.001 | 0.23 [0.12-0.47] | <0.001 | 0.28 [0.16-0.47] | <0.001 | 0.29 [0.14-0.62] | 0.001 |
| Q3 | 7585 | 4037 (53.22) | 91 (2.25) | 3548 (46.78) | 288 (8.12) | 0.27 [0.21-0.34] | <0.001 | 0.31 [0.23-0.42] | <0.001 | 0.26 [0.14-0.47] | <0.001 | 0.54 [0.37-0.78] | 0.001 | 0.71 [0.45-1.13] | 0.145 |
| Q4 | 7558 | 4162 (55.07) | 137 (3.29) | 3396 (44.93) | 413 (12.16) | 0.26 [0.21-0.31] | <0.001 | 0.35 [0.28-0.44] | <0.001 | 0.38 [0.25-0.58] | <0.001 | 0.42 [0.3-0.6] | <0.001 | 0.4 [0.24-0.67] | <0.001 |
| ***Variance of HbA1c*** | | | | | | | | | | | | | | | |
| Q1 | 5751 | 2862 (49.77) | 58 (2.03) | 2889 (50.23) | 252 (8.72) | 0.22 [0.17-0.3] | <0.001 | 0.25 [0.17-0.37] | <0.001 | 0.29 [0.15-0.54] | <0.001 | 0.35 [0.21-0.57] | <0.001 | 0.6 [0.34-1.08] | 0.087 |
| Q2 | 5974 | 3258 (54.54) | 70 (2.15) | 2716 (45.46) | 211 (7.77) | 0.27 [0.2-0.35] | <0.001 | 0.35 [0.25-0.5] | <0.001 | 0.23 [0.11-0.49] | <0.001 | 0.52 [0.34-0.79] | 0.0024 | 0.47 [0.25-0.89] | 0.021 |
| Q3 | 5927 | 3194 (53.89) | 78 (2.44) | 2733 (46.11) | 218 (7.98) | 0.3 [0.23-0.38] | <0.001 | 0.42 [0.31-0.57] | <0.001 | 0.28 [0.15-0.55] | <0.001 | 0.45 [0.28-0.7] | <0.001 | 0.42 [0.22-0.82] | 0.011 |
| Q4 | 5962 | 3158 (52.97) | 110 (3.48) | 2804 (47.03) | 367 (13.09) | 0.25 [0.2-0.31] | <0.001 | 0.36 [0.28-0.47] | <0.001 | 0.32 [0.2-0.53] | <0.001 | 0.4 [0.27-0.57] | <0.001 | 0.35 [0.19-0.64] | <0.001 |
| ***CV of HbA1c*** | | | | | | | | | | | | | | | |
| Q1 | 5759 | 2951 (51.24) | 56 (1.9) | 2808 (48.76) | 272 (9.69) | 0.19 [0.14-0.25] | <0.001 | 0.18 [0.12-0.28] | <0.001 | 0.29 [0.16-0.52] | <0.001 | 0.36 [0.23-0.59] | <0.001 | 0.56 [0.32-1.0] | 0.051 |
| Q2 | 5759 | 3139 (54.51) | 72 (2.29) | 2620 (45.49) | 168 (6.41) | 0.35 [0.26-0.46] | <0.001 | 0.49 [0.36-0.68] | <0.001 | 0.2 [0.08-0.49] | <0.001 | 0.58 [0.37-0.9] | 0.015 | 0.56 [0.3-1.05] | 0.070 |
| Q3 | 5751 | 3133 (54.48) | 79 (2.52) | 2618 (45.52) | 264 (10.08) | 0.24 [0.19-0.31] | <0.001 | 0.34 [0.25-0.47] | <0.001 | 0.31 [0.18-0.56] | 0.001 | 0.43 [0.28-0.66] | <0.001 | 0.27 [0.13-0.57] | <0.001 |
| Q4 | 5769 | 3005 (52.09) | 104 (3.46) | 2764 (47.91) | 327 (11.83) | 0.28 [0.23-0.35] | <0.001 | 0.39 [0.3-0.51] | <0.001 | 0.33 [0.2-0.55] | <0.001 | 0.39 [0.26-0.58] | <0.001 | 0.46 [0.26-0.81] | 0.007 |
| ***Baseline fasting glucose (mmol/L)*** | | | | | | | | | | | | | | | |
| <5.6 | 6669 | 2807 (42.09) | 79 (2.81) | 3862 (57.91) | 445 (11.52) | 0.23 [0.18-0.29] | <0.001 | 0.29 [0.21-0.39] | <0.001 | 0.28 [0.16-0.48] | <0.001 | 0.33 [0.22-0.51] | <0.001 | 0.38 [0.21-0.67] | <0.001 |
| 5.6-6.9 | 5092 | 2382 (46.78) | 44 (1.85) | 2710 (53.22) | 297 (10.96) | 0.16 [0.12-0.22] | <0.001 | 0.28 [0.2-0.41] | <0.001 | 0.11 [0.04-0.3] | <0.001 | 0.14 [0.06-0.29] | <0.001 | 0.21 [0.09-0.52] | <0.001 |
| >6.9 | 19994 | 10679 (53.41) | 280 (2.62) | 9315 (46.59) | 846 (9.08) | 0.28 [0.24-0.32] | <0.001 | 0.36 [0.31-0.43] | <0.001 | 0.32 [0.24-0.44] | <0.001 | 0.44 [0.35-0.56] | <0.001 | 0.48 [0.35-0.67] | <0.001 |
| ***Mean fasting glucose (mmol/L)*** | | | | | | | | | | | | | | | |
| Q1 | 7111 | 3285 (46.2) | 74 (2.25) | 3826 (53.8) | 430 (11.24) | 0.19 [0.15-0.24] | <0.001 | 0.28 [0.2-0.38] | <0.001 | 0.15 [0.08-0.31] | <0.001 | 0.28 [0.18-0.43] | <0.001 | 0.24 [0.12-0.48] | <0.001 |
| Q2 | 7254 | 3645 (50.25) | 73 (2.0) | 3609 (49.75) | 230 (6.37) | 0.31 [0.23-0.4] | <0.001 | 0.36 [0.25-0.5] | <0.001 | 0.3 [0.15-0.58] | <0.001 | 0.5 [0.33-0.76] | 0.001 | 0.49 [0.26-0.91] | 0.025 |
| Q3 | 7195 | 3979 (55.3) | 96 (2.41) | 3216 (44.7) | 323 (10.04) | 0.23 [0.18-0.29] | <0.001 | 0.33 [0.25-0.44] | <0.001 | 0.3 [0.18-0.51] | <0.001 | 0.36 [0.24-0.54] | <0.001 | 0.55 [0.34-0.9] | 0.016 |
| Q4 | 7192 | 3837 (53.35) | 140 (3.65) | 3355 (46.65) | 392 (11.68) | 0.3 [0.24-0.36] | <0.001 | 0.38 [0.3-0.49] | <0.001 | 0.37 [0.25-0.57] | <0.001 | 0.46 [0.32-0.64] | <0.001 | 0.47 [0.29-0.76] | 0.002 |
| ***Variance of fasting glucose*** | | | | | | | | | | | | | | | |
| Q1 | 4844 | 2616 (54.0) | 56 (2.14) | 2228 (46.0) | 123 (5.52) | 0.38 [0.28-0.52] | <0.001 | 0.58 [0.41-0.84] | 0.004 | 0.4 [0.2-0.82] | 0.012 | 0.35 [0.18-0.66] | 0.001 | 0.53 [0.25-1.14] | 0.104 |
| Q2 | 4847 | 2888 (59.58) | 59 (2.04) | 1959 (40.42) | 202 (10.31) | 0.19 [0.14-0.25] | <0.001 | 0.27 [0.18-0.38] | <0.001 | 0.27 [0.14-0.53] | <0.001 | 0.46 [0.3-0.72] | <0.001 | 0.3 [0.13-0.66] | 0.003 |
| Q3 | 4844 | 2771 (57.2) | 70 (2.53) | 2073 (42.8) | 156 (7.53) | 0.33 [0.25-0.43] | <0.001 | 0.38 [0.27-0.54] | <0.001 | 0.52 [0.3-0.91] | 0.022 | 0.58 [0.37-0.92] | 0.020 | 0.71 [0.38-1.35] | 0.297 |
| Q4 | 4847 | 2321 (47.89) | 100 (4.31) | 2526 (52.11) | 440 (17.42) | 0.23 [0.18-0.28] | <0.001 | 0.29 [0.22-0.38] | <0.001 | 0.3 [0.18-0.49] | <0.001 | 0.32 [0.22-0.48] | <0.001 | 0.41 [0.24-0.71] | 0.001 |
| ***CV of fasting glucose*** | | | | | | | | | | | | | | | |
| Q1 | 4723 | 2573 (54.48) | 57 (2.22) | 2150 (45.52) | 126 (5.86) | 0.37 [0.27-0.51] | <0.001 | 0.54 [0.37-0.78] | 0.001 | 0.44 [0.22-0.86] | 0.016 | 0.4 [0.23-0.73] | 0.002 | 0.53 [0.25-1.12] | 0.095 |
| Q2 | 4722 | 2852 (60.4) | 52 (1.82) | 1870 (39.6) | 118 (6.31) | 0.28 [0.2-0.39] | <0.001 | 0.33 [0.21-0.5] | <0.001 | 0.46 [0.24-0.87] | 0.016 | 0.63 [0.39-1.02] | 0.059 | 0.46 [0.2-1.04] | 0.062 |
| Q3 | 4716 | 2680 (56.83) | 75 (2.8) | 2036 (43.17) | 222 (10.9) | 0.24 [0.19-0.32] | <0.001 | 0.34 [0.25-0.46] | <0.001 | 0.27 [0.14-0.52] | <0.001 | 0.39 [0.24-0.62] | <0.001 | 0.62 [0.35-1.1] | 0.104 |
| Q4 | 4730 | 2252 (47.61) | 84 (3.73) | 2478 (52.39) | 400 (16.14) | 0.21 [0.17-0.27] | <0.001 | 0.28 [0.21-0.38] | <0.001 | 0.31 [0.18-0.52] | <0.001 | 0.32 [0.21-0.49] | <0.001 | 0.29 [0.14-0.58] | <0.001 |

CI: confidence interval; CV: coefficient of variation; DPP4i: dipeptidyl peptidase-4 inhibitor; HbA1c: hemoglobin A1c; HR: hazard ratio; SGLT2i: sodium-glucose cotransporter-2 inhibitor.

Table S15. The associations between SGLT2i vs. DPP4i vs. GLP-1RA and outcomes using stabilized IPTW.

| **Model** | **New-onset syncope** | | **Cardiovascular mortality** | | **All-cause mortality** | |
| --- | --- | --- | --- | --- | --- | --- |
|  | **HR [95% CI]** | **P value** | **HR [95% CI]** | **P value** | **HR [95% CI]** | **P value** |
| SGLT2i vs. DPP4i | 0.32 [0.25-0.42] | <0.001 | 0.45 [0.32-0.66] | <0.001 | 0.26[0.18-0.39] | <0.001 |
| SGLT2i vs. GLP-1RA | 0.89 [0.66-1.19] | 0.382 | 0.93 [0.77-1.45] | 0.682 | 0.90[0.66-1.26] | 0.509 |
| GLP-1RA vs. DPP4i | 0.46 [0.35-0.92] | <0.001 | 0.65 [0.35-0.75] | <0.001 | 0.39[0.42-0.55] | <0.001 |

CI: confidence interval; DPP4i: dipeptidyl peptidase-4 inhibitor; GLP-1RA: glucagon-like peptide-1 receptor agonist; HR: hazard ratio; IPTW: inverse probability of treatment weighting; SGLT2i: sodium glucose cotransporter-2 inhibitor.

The results from three-arm (SGLT2i vs. DPP4i vs. GLP-1RA) as-treat analysis showed that both SGLT2i (HR, 0.32; 95%CI [0.25-0.42],	P<0.001) and GLP-1RA (HR, 0.46; 95%CI [0.35-0.92],	P<0.001) had protective effects on new-onset syncope than DPP4i. In addition, we have observed a favorable signal for SGLT2i in preventing syncope than GLP-1RA, whereas the difference did not reach the traditional significance (HR, 0.89; 95%CI [0.66-1.19], P=0.3815). It should be noted that the sample size of the GLP-1RA cohort in our database is relatively small, which may introduce selection bias.
